# A deep learning transformer model predicts high rates of undiagnosed rare disease in large electronic health systems

**DOI:** 10.1101/2023.12.21.23300393

**Authors:** Daniel M. Jordan, Ha My T. Vy, Ron Do

## Abstract

It is estimated that as many as 1 in 16 people worldwide suffer from rare diseases. Rare disease patients face difficulty finding diagnosis and treatment for their conditions, including long diagnostic odysseys, multiple incorrect diagnoses, and unavailable or prohibitively expensive treatments. As a result, it is likely that large electronic health record (EHR) systems include high numbers of participants suffering from undiagnosed rare disease. While this has been shown in detail for specific diseases, these studies are expensive and time consuming and have only been feasible to perform for a handful of the thousands of known rare diseases. The bulk of these undiagnosed cases are effectively hidden, with no straightforward way to differentiate them from healthy controls. The ability to access them at scale would enormously expand our capacity to study and develop drugs for rare diseases, adding to tools aimed at increasing availability of study cohorts for rare disease. In this study, we train a deep learning transformer algorithm, RarePT (Rare-Phenotype Prediction Transformer), to impute undiagnosed rare disease from EHR diagnosis codes in 436,407 participants in the UK Biobank and validated on an independent cohort from 3,333,560 individuals from the Mount Sinai Health System. We applied our model to 155 rare diagnosis codes with fewer than 250 cases each in the UK Biobank and predicted participants with elevated risk for each diagnosis, with the number of participants predicted to be at risk ranging from 85 to 22,000 for different diagnoses. These risk predictions are significantly associated with increased mortality for 65% of diagnoses, with disease burden expressed as disability-adjusted life years (DALY) for 73% of diagnoses, and with 72% of available disease-specific diagnostic tests. They are also highly enriched for known rare diagnoses in patients not included in the training set, with an odds ratio (OR) of 48.0 in cross-validation cohorts of the UK Biobank and an OR of 30.6 in the independent Mount Sinai Health System cohort. Most importantly, RarePT successfully screens for undiagnosed patients in 32 rare diseases with available diagnostic tests in the UK Biobank. Using the trained model to estimate the prevalence of undiagnosed disease in the UK Biobank for these 32 rare phenotypes, we find that at least 50% of patients remain undiagnosed for 20 of 32 diseases. These estimates provide empirical evidence of a high prevalence of undiagnosed rare disease, as well as demonstrating the enormous potential benefit of using RarePT to screen for undiagnosed rare disease patients in large electronic health systems.

## Introduction

Rare diseases, also known as orphan diseases, are defined by the European Union as those affecting fewer than one in 2,000 people, and in the United States as those affecting fewer than 200,000 people nationwide^1,2^. Rare diseases are collectively very common, and it is estimated that as many as 1 in 16 people (6.2%) suffer from one or more rare diseases^3^. This makes them a serious public health concern, as rare disease patients are far less likely to receive accurate diagnoses or, once diagnosed, to have access to effective treatments for their conditions^4–6^. This is due to the difficulty of studying rare diseases, a scarcity of clinical expertise and diagnostic methods, as well as the unprofitability of developing drugs targeting them. In fact, many of these diseases are so understudied and underdiagnosed that we do not know with any certainty what their true prevalence is, and how many undiagnosed patients there may be^3^. One of the primary reasons for all these problems is the difficulty of finding large enough populations of patients to conduct well-powered studies on these diseases, either in the context of basic or translational research or in the context of drug trials. This is a pressing problem, and tools such as MatchMaker Exchange, which help researchers match similar cases to increase sample size for studies of rare diseases, are widely used^7–9^. Further development of these tools is also an active area of research, including expanding them to include comparisons of phenotypic features mined from electronic health records (EHR) or imaging data^9–12^. These tools are vital for rare disease research because undiagnosed rare disease patients masquerade as healthy controls, making them invisible and inaccessible to researchers unless they can be revealed. There is an urgent need for new approaches to reveal people suffering from hidden rare diseases in research and drug trial cohorts, and in clinical practice.

In this study, we present such an approach, using a deep learning transformer model trained on EHR data. Artificial intelligence (AI) language models based on the transformer architecture, such as BERT (“Bidirectional Encoder Representations from Transformers” and GPT (“Generative Pretrained Transformers”), have proved very successful at learning the relationships between concepts in natural languages^13,14^. Transformer models have also been successfully applied to problems in biology that are not directly related to language processing in methods such as AlphaFold-2, AlphaMissense, DeepMAPS, and Enformer^15–18^. One of the strength of the transformer architecture is that, with appropriate tokenization and training schemes, transformers can correctly model concepts that are extremely rare, and some transformer-based models have been shown to define a new word after seeing it only a small number of times^19–22^. We have designed a modified transformer architecture to model phenotypic concepts based on phenotypes derived from structured diagnosis codes in electronic health records (EHR), along with a modified training procedure designed to maximize power to screen for missing rare diagnoses. The resulting model, RarePT (Rare-Phenotype Prediction Transformer), was trained on EHR data from 436,407 individuals from the UK Biobank and validated on an independent cohort from 3,333,560 individuals from the Mount Sinai Health System in New York City, USA (**Table 1**). RarePT shows remarkable power to recapitulate rare diagnoses, which is robust across different racial and ethnic groups, different hospitals with different coding practices, and even different countries with different health care standards and coding vocabularies. It also detects UK Biobank participants with undiagnosed rare disease, enabling empirical measurement of the true prevalence of undiagnosed cases for rare diseases.

**Table 1.**
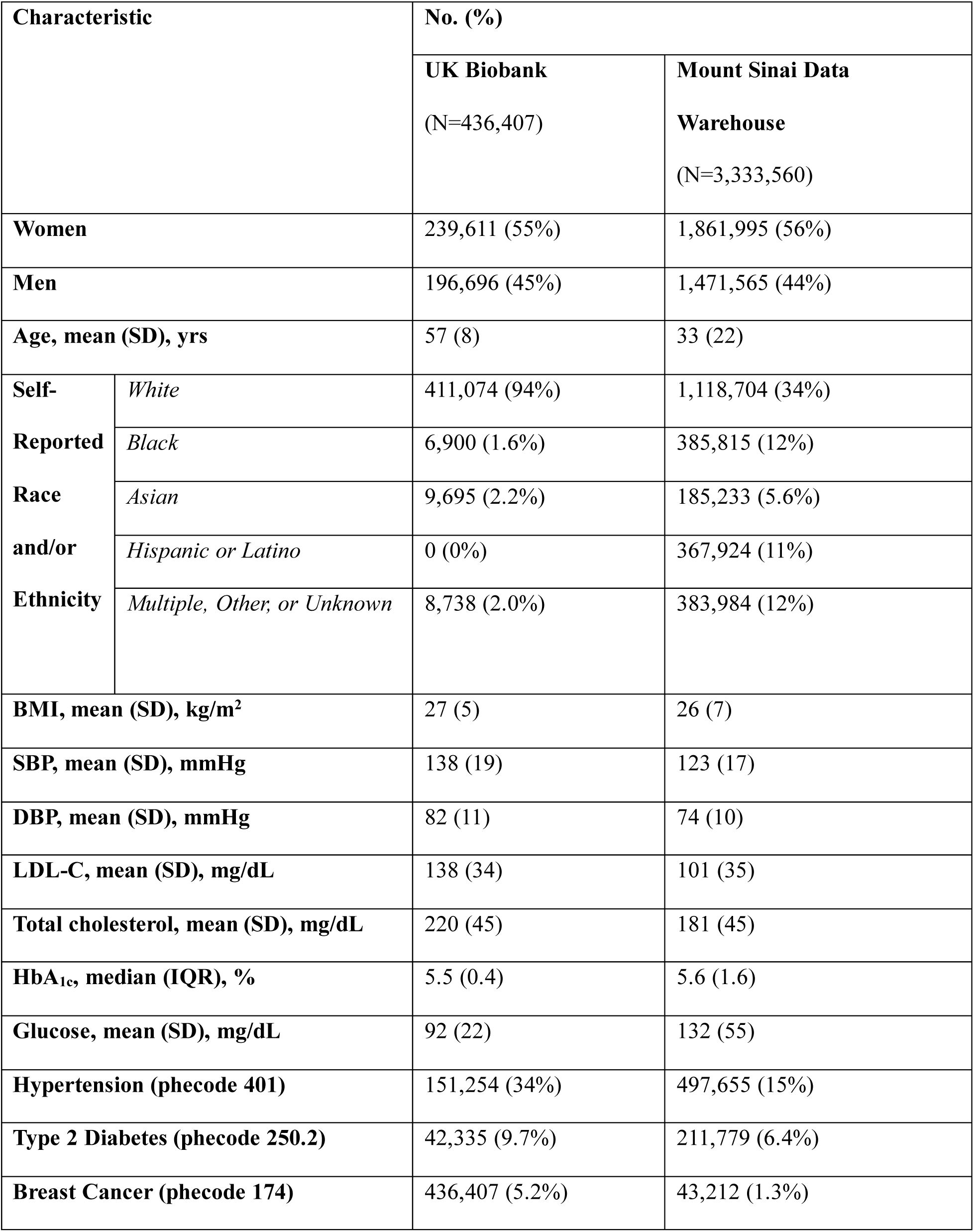
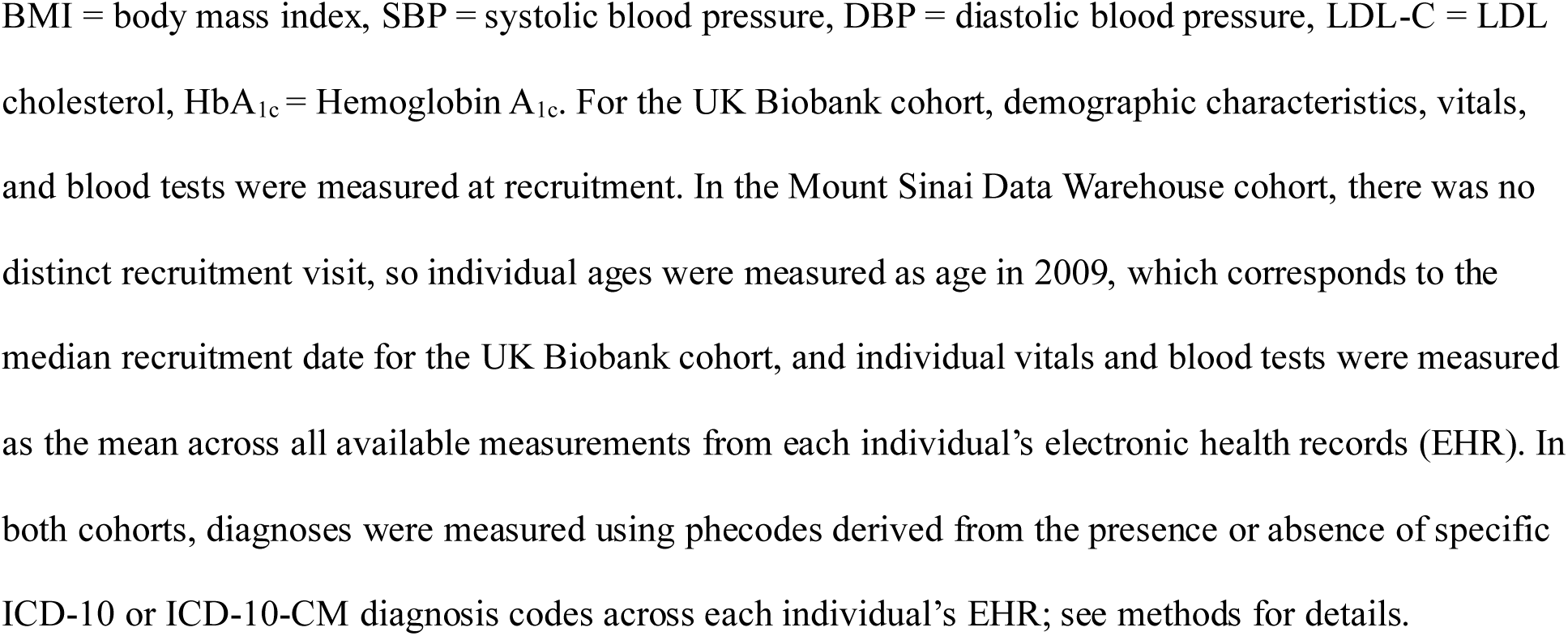
Baseline demographic and clinical characteristics of cohorts.

| Characteristic |  | No. (%) |  |
| --- | --- | --- | --- |
|  |  | UK Biobank<br>(N=436,407) | Mount Sinai Data<br>Warehouse<br>(N=3,333,560) |
| **Women** |  | 239,611 (55%) | 1,861,995 (56%) |
| **Men** |  | 196,696 (45%) | 1,471,565 (44%) |
| **Age, mean (SD), yrs** |  | 57 (8) | 33 (22) |
| **Self-**<br>**Reported**<br>**Race**<br>**and/or**<br>**Ethnicity** | *White* | 411,074 (94%) | 1,118,704 (34%) |
|  | *Black* | 6,900 (1.6%) | 385,815 (12%) |
|  | *Asian* | 9,695 (2.2%) | 185,233 (5.6%) |
|  | *Hispanic or Latino* | 0 (0%) | 367,924 (11%) |
|  | *Multiple, Other, or Unknown* | 8,738 (2.0%) | 383,984 (12%) |
| **BMI, mean (SD), kg/m<sup>2</sup>** |  | 27 (5) | 26 (7) |
| **SBP, mean (SD), mmHg** |  | 138 (19) | 123 (17) |
| **DBP, mean (SD), mmHg** |  | 82 (11) | 74 (10) |
| **LDL-C, mean (SD), mg/dL** |  | 138 (34) | 101 (35) |
| **Total cholesterol, mean (SD), mg/dL** |  | 220 (45) | 181 (45) |
| **HbA<sub>1c</sub>, median (IQR), %** |  | 5.5 (0.4) | 5.6 (1.6) |
| **Glucose, mean (SD), mg/dL** |  | 92 (22) | 132 (55) |
| **Hypertension (phecode 401)** |  | 151,254 (34%) | 497,655 (15%) |
| **Type 2 Diabetes (phecode 250.2)** |  | 42,335 (9.7%) | 211,779 (6.4%) |
| **Breast Cancer (phecode 174)** |  | 436,407 (5.2%) | 43,212 (1.3%) |

BMI = body mass index, SBP = systolic blood pressure, DBP = diastolic blood pressure, LDL-C = LDL cholesterol, HbA<sub>1c</sub> = Hemoglobin A<sub>1c</sub>. For the UK Biobank cohort, demographic characteristics, vitals, and blood tests were measured at recruitment. In the Mount Sinai Data Warehouse cohort, there was no distinct recruitment visit, so individual ages were measured as age in 2009, which corresponds to the median recruitment date for the UK Biobank cohort, and individual vitals and blood tests were measured as the mean across all available measurements from each individual's electronic health records (EHR). In both cohorts, diagnoses were measured using phcodes derived from the presence or absence of specific ICD-10 or ICD-10-CM diagnosis codes across each individual's EHR; see methods for details.

## Results

### Model training and cross-validation

We implemented a transformer model with a self-attention mechanism similar to AI language models such as BERT and GPT, along with a “masked diagnosis modeling” training objective by analogy to the “masked language modeling” objective used by some of these language models^19^. In this approach, training examples consist of complete sequences with a single token removed, and the model is trained to reconstruct the missing token. In the natural language processing case, this is a sequence of words with a single word removed; in our case, it is a participant record with a single diagnosis removed (**Figure 1a**). The model learns the meanings of tokens based on the context they appear in, resulting in embeddings that cluster tokens that commonly appear together and tokens that appear in similar context. Models trained with this objective are known to learn informative embeddings even for very rare tokens in many cases^19–22^. We made use of this feature to train a model to predict rare diagnoses, a critical need due to underdiagnosis and understudying of rare diseases.

**Figure 1.**
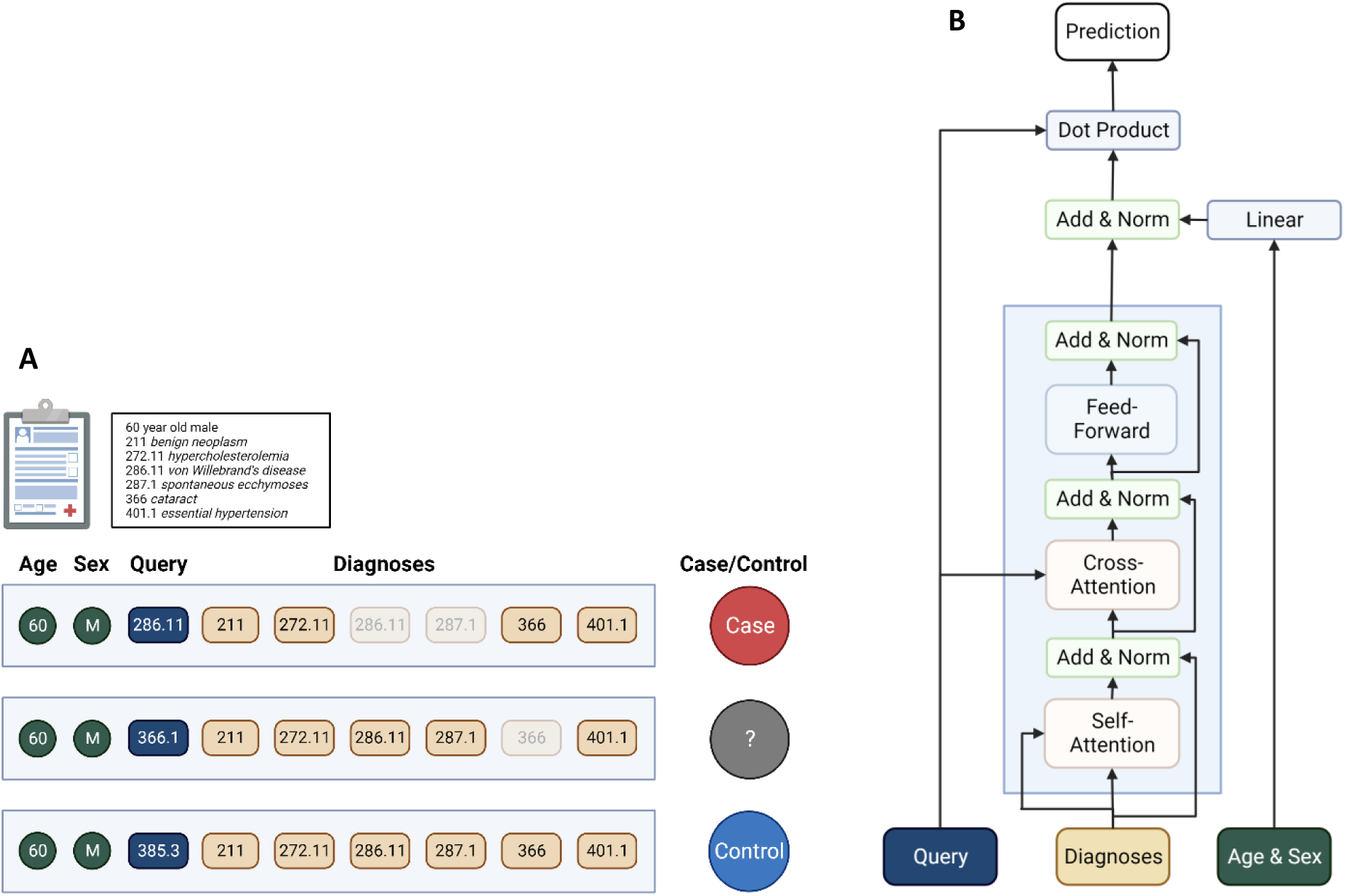
Schematics of masked phenotype modeling training procedure and RarePT model architecture. *(a) Masked phenotype modeling training procedure.* Each individual is represented by demographic characteristics (age and sex) and the set of all diagnoses present in their EHR across all encounters, represented as phecodes. A single training example consists of all demographic characteristics, a “query” phecode indicating the phecode to be trained on, and the target case/control designation. The example is considered a “Case” if the individual’s EHR contains the query phecode, “Unknown” if the individual’s EHR does not contain the query phecode but does contain a phecode defined as an exclusion for the query phecode, and “Control” if the individual’s EHR contains neither the query phecode nor any phecode defined as an exclusion for the query phecode. In all instances, the query phecode and all phecodes defined as exclusions for the query phecode are hidden before training or prediction. *(b) Rare-Phenotype Prediction Transformer (RarePT) architecture.* We apply a simplified transformer architecture to the diagnoses. Diagnoses are fed into a transformer decoder in a many-hot encoding, in a single step with no sequence. The transformer decoder consists of a self-attention layer, a cross-attention attending to the query phecode, and a feed-forward layer, all connected by residual skip connections, as in the standard transformer architecture. Demographic characteristics are interpreted by a dense linear layer and then applied as an adjustment to the transformer output. The final prediction is the dot product of the query phecode input with the demographic-adjusted transformer output.

An additional advantage of the masked diagnosis modeling training objective for rare tokens is that it allows us to weight the importance of tokens to the training objective independent of their prevalence in the training corpus. This is because each training example specifies which token the model must predict correctly to be scored as successful, and the model is not necessarily required to predict every token in each example. The importance of each token to the training objective is determined by how many examples have it as the masked token. In order to prevent very common diagnoses from dominating the learned embeddings, we limited training examples to a fixed number of cases and controls for each diagnosis. While we used 100 cases and controls for each diagnosis, in principle this is a tunable parameter of the training process. Lower values allow rarer diagnoses to be included, while higher values increase the amount of training data available.

In this study, we express diagnoses as phecodes^23^. We determined phecodes from ICD-10 codes for 436,407 participants in the UK Biobank based on a standard mapping^24^. We then filtered out all phecodes with fewer than 100 cases and controls and constructed a training dataset consisting of 100 randomly selected cases and 100 randomly selected controls for each phecode. The resulting training set consisted of 259,400 training examples representing 1,297 query phecodes and 111,331 unique participants. For each training example, input data included the following features:

1. The identity of the query phecode
2. All other phecodes for which the participant is considered a case
3. Age at recruitment
4. Sex reported from recruitment

These training examples were split into 5 subsamples for cross-validation, stratified so that each participant appeared in only one split and so that each split contained a similar number of cases and controls. Neural network architecture and other training hyperparameters were tuned on the training data for each split using the Hyperband algorithm^25^, and then the tuned model was trained on the same data and tested on the held-out test data; see Methods for details of model tuning and training parameters. The final tuned architecture is shown in in **Figure 1b**; details of training performance can be found in **Supplementary Figure S1** and **Supplementary Table S1**. In general, the models performed well on the test data and showed only minor loss of performance between training and test data.

### RarePT predicts rare diagnoses in the UK Biobank

To test the ability of the trained model to predict rare diagnoses, we first selected all phecodes appearing in fewer than 1 in 2,000 UK Biobank participants, corresponding to the definition of rare diseases used by the European Union^2^. There were 155 rare phecodes meeting this criterion, shown in **Supplementary Table S2**. Not all phecodes that are rare in the UK Biobank represent phenotypes that meet the definition of rare diseases in the general population. One reason for this is the known bias of the UK Biobank population towards healthier and older participants^26–28^, which reduces the apparent prevalence of many diseases, especially severe diseases with early onset. For example, phecode 315.3 “mental retardation” appears in fewer than 200 participants in the UK Biobank even though the disorders it represents are much more common in the general population, which is likely because severe childhood disorders are underrepresented in this cohort of healthy adults. It is also likely that many of these rare phecodes correspond to diagnosis codes that rarely appear in electronic health records (EHR) despite the conditions they refer to being common. Likely examples of this include 367.4 “presbyopia” and 523.1 “gingivitis.” Nevertheless, even if not all of these phecodes represent phenotypes that are rare in the general population, they do represent phenotypes that are rare in the data used to train our model, and the model’s performance on these phecodes is informative about how our methodology handles rare phenotypes. In total, 21,636 of the 436,407 participants tested have one or more of these rare diagnoses, giving them a cumulative prevalence of 5.0%. This matches the estimated cumulative prevalence of 1.5-6.2% for rare diseases in the general population^3^, suggesting that our selection of rare phecodes does accurately capture the population distribution of rare diseases.

After training on a 111,311-participant subset of UK Biobank data constructed to force each phecode to have prevalence of 50%, we measured RarePT’s performance in the full UK Biobank dataset of 436,407 participants. We arbitrarily chose a threshold of 0.95 in the model’s probability score output, so that participants with a score of 0.95 or higher in a given phecode were treated as positive predictions for that phecode. With this definition, across all five cross-validated models, we generate specific positive predictions for each of our 155 rare phecodes. The number of positive predictions varied by phecode, ranging between 85 and 22,000 with a median of 2,135 positive predictions per phecode (**Supplementary Table 3**). These positive predictions are broadly distributed across participants rather than being concentrated in a small group of unhealthy participants, with no participant receiving more than 29 positive predictions and 41% of participants (177,484) receiving a positive prediction for at least one of the 155 phecodes. **Figure 2** shows the performance of the 5 cross-validated models at predicting rare phecodes in the full dataset, excluding each model’s training data. We measured prediction performance using diagnostic odds ratio (OR), defined as the ratio between the odds of a participant having a diagnosis given a positive prediction from the model and the odds of a participant having a diagnosis given a negative prediction from the model. The median OR for a positive prediction across all 155 rare phecodes and across the five models trained in cross-validation was 48.0. Some specific phecodes reached a median OR over 20,000, and the lowest median OR for any rare phecode was 5.13 (**Figure 2a, Supplementary Table S3**). These values compare favorably to many commonly used diagnostic tests, where diagnostic odds ratios in the range of 20-50 are considered very good^29,30^. Similarly, the positive predictive value (PPV) for cases is nearly 40% for some phecodes, which is well within the range of a useful screening test (**Supplementary Figure S2, Supplementary Table S3**). Because PPV depends on the prevalence of the condition within the test population, we expect this number to increase further when applying this method in situations where the prior expectation of encountering a given diagnosis is increased, such as in patients with undiagnosed rare conditions or patients who carry rare genetic variants. Importantly, the predictions are able to distinguish not only between cases and controls but also between cases for one phecode and cases for another, indicating that RarePT is making specific predictions for each phecode rather than measuring general health (**Figure 2b**).

**Figure 2.**
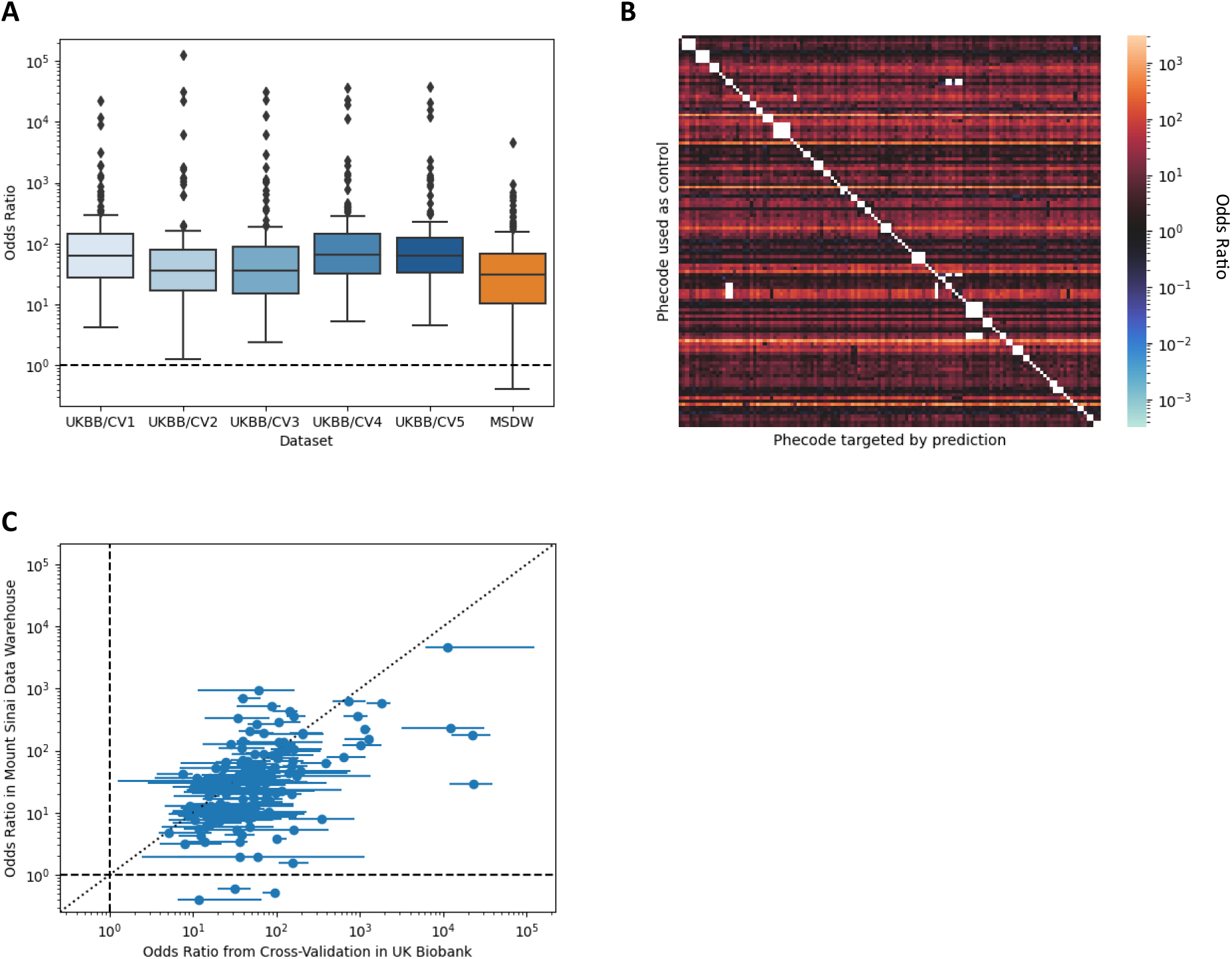
Performance of RarePT to predict diagnosed cases of 155 rare phecodes. (a) Box and whisker plots showing distribution of diagnostic odds ratio (ratio of odds in predicted cases to odds in predicted controls) across rare phecodes. The five blue boxes labelled “UKBB/CV1-5” show the performance of the five models trained using five-fold cross-validation within the UK Biobank, excluding each model’s training set; median OR across all five cross-validation sets was 48 (full range 1.27-39,000). The orange box labeled “MSDW” shows the performance of the model trained using the full UK Biobank dataset on the Mount Sinai Data Warehouse dataset, an independent cohort from the Mount Sinai Health System in New York; median OR for this cohort was 31 (full range 0.41-4,600). Performance on the independent dataset is only slightly reduced, despite extensive differences between these datasets and health systems, demonstrating the robustness of the approach. Boxes show 1^st^ quartile, median, and 3^rd^ quartile; whiskers extend to 80% of interquartile range; dashed line shows random chance. *(b) Heatmap showing odds ratios (median across five-fold cross-validation) for distinguishing between diagnosed cases of one phecode and diagnosed cases of a second phecode.* Columns represent different case phecodes, rows represent different control phecodes, both ordered so that phecodes in the same biological category are grouped together. For each cell, the RarePT models trained in cross-validation were presented with a dataset consisting of all diagnosed cases of both phecodes in UK Biobank, excluding the model’s own training set. The model was asked to predict case status for the case phecode, treating cases of the control phecode as controls. Red indicates that the model’s case predictions are enriched for cases of the correct phecode, while blue indicates that the model’s case predictions are depleted for cases of the correct phecode. Blank (white) cells indicate phecodes that are labelled as exclusions in the definition of the target phecode. While some comparisons are more successful than others, mostly depending on the identity of the phecode used as controls, RarePT successfully predicts cases across a wide range of rare phecodes, even when the background is cases for other rare phecodes. Blue This demonstrates that RarePT’s predictions are specific to each specific phecode, and are not primarily predicting categories of diagnoses or general health. *(c) Scatterplot comparing RarePT’s performance for rare phecodes on UK Biobank and Mount Sinai Data Warehouse cohorts.* Each data point represents a single phecode; horizontal error bars represent the results of five-fold cross validation within the UK Biobank cohort. Performance for specific phecodes is strongly correlated between cohorts (Pearson r = 0.456, p = 5.03 × 10^-40^, t-test), demonstrating that the specific features used to differentiate different phecodes are robust to differences between cohorts, including differences in diagnosis coding, recruitment, and ethnic and racial composition. Dashed lines show random chance within each cohort; dotted line shows equal performance between cohorts.

### Model trained on UK Biobank is predictive in an independent EHR cohort

We applied the trained RarePT model to an independent dataset derived from the Mount Sinai Data Warehouse (MSDW), consisting of anonymized EHR for a cohort of 3,333,560 patients seen in the Mount Sinai Health System in New York City. We determined phecodes for these participants in the same way as for the UK Biobank participants, but using a mapping designed for the US clinical modification to ICD-10 (ICD-10CM) rather the international standard ICD-10 system used by UK hospitals^24^. 151 of the 155 phecodes determined to be rare in the UK Biobank cohort were present in the MSDW cohort. As a health system based cohort, this cohort is expected to be significantly less healthy than the UK Biobank^31^, and therefore we expect most phecodes to have higher prevalence than in the UK Biobank cohort. Nevertheless, a majority of the rare phecodes we tested (86/151; 57%) still had prevalence less than 1 in 2,000 in the MSDW cohort (**Supplementary Table S2**). Likewise, we expect more positive predictions for each phecode, both due to the dataset being over 7-fold larger and due to participants being less healthy in general.

For these phecodes in the MSDW cohort, RarePT produced between 100 and 721,000 positive predictions per phecode, with a median of 11,500, and produced at least one positive prediction in 47% of participants (1,518,757). These predictions performed similarly to the predictions for the UK Biobank cohort, with a median OR of 30.6 across all 151 phecodes **(Figure 2a, Supplementary Table S4)**. Performance for individual phecodes was also strongly correlated across the two datasets (Pearson r = 0.456, p = 5.03 × 10^-40^, t-test; **Figure 2c**). The fact that performance is similar across the two datasets indicates that RarePT’s predictions are based on features that are robust to different methodologies for sample ascertainment and data collection, rather than features that are only informative in the specialized context of the UK Biobank. This replication is especially remarkable given the extensive differences between the two cohorts: in addition to one being a population-based cohort of healthy volunteers and the other being a health system cohort, these cohorts are also from different countries with different standard medical practices, different billing structures and coding systems, and different distributions of race, ethnicity, and genetic ancestry. This indicates the wide applicability of the RarePT method and suggests that its performance does not depend on specific features of diagnosis coding in a particular health system.

### Rare disease predictions are associated with mortality, disease burden, and known diagnostic biomarkers

To further demonstrate that RarePT is capturing clinically relevant signals of disease rather than bioinformatic artifacts related to diagnosis coding, we performed regression analyses to test the association of positive predictions with mortality, disability, and, where available, known diagnostic biomarkers. We retrieved the latest mortality data for UK Biobank participants as of October 2023, and performed Cox proportional hazard regression to test whether a positive prediction is associated with mortality, controlling for age, sex, and self-reported ethnicity. 101 phecodes (65% of phecodes tested) had a significant association (p < 0.05) with increased mortality, of which 93 (60%) remained significant after Bonferroni correction for 155 phecodes (p < 0.00032). The median phecode had a regression coefficient of 0.70, corresponding to a hazard ratio of 2.01, or a twofold increase in mortality rate (**Supplementary Table S5**).

Next, we estimated Disability Adjusted Life Years (DALY) and its two components, Years of Life Lost (YLL) and Years Living with Disability (YLD), for 80 conditions for all UK Biobank participants^32^. These measurements represent the number of years lost to both mortality and disability as a result of illness and are used as a measure of disease burden, particularly in the Global Burden of Disease study^33^. We performed linear regressions with DALY, YLD, and YLL as the dependent variables to test whether a positive prediction is associated with greater disease burden. In all of these regressions, we controlled for age, sex, and self-reported ethnicity. 113 phecodes (73% of phecodes tested) had a significant association (p < 0.05) with increased estimated DALY, and 106 (68%) remained significant after Bonferroni correction for 155 phecodes (p < 0.00032). 134 phecodes (87%) had a significant association with increased estimated YLD individually, 133 (86%) after Bonferroni correction; 110 phecodes (71%) had a significant association with increased estimated YLL individually, 106 (68%) after Bonferroni correction. For the median phecode, a positive prediction was associated with an increase in estimated DALY of 1.1 years (**Supplementary Table S5**).

To identify diagnostic biomarkers, we used the SNOMED-CT vocabulary of clinical terms^34,35^ to identify phenotypes whose clinical definition includes laboratory tests that are available for large numbers of participants in the UK Biobank. We identified 75 defined relationships between 32 rare phecodes and 23 laboratory tests (**Supplementary Table S6**). These tests were performed as part of the UK Biobank recruitment process and were generally not returned to participants or their physicians, so the availability of a test result does not indicate that it was ordered by a physician and the result of a test was not visible to the physicians responsible for entering diagnoses into the participants’ EHR. Since RarePT makes its predictions using only diagnosis codes and has no access to physician-ordered laboratory tests except through diagnosis codes, this means our model’s predictions are independent of these test results. This is in contrast to health system based cohorts, including our MSDW cohort, where diagnostic tests are ordered and administered in the context of treating the patient, so that the presence and timing of a test are informative about the judgment of the health care providers and the test result forms part of the diagnostic criteria^36^.

For each of these 75 relationships, we performed a logistic regression to test whether a confident case prediction is associated with abnormal test results, again controlling for age, sex, and ethnicity. 54 of these regressions, representing 72% of these relationships, had a result that was in the expected direction and statistically significant (p < 0.05), and 45 (60%) remained significant after Bonferroni correction for 75 regressions (p < 0.00067). The median regression coefficient was 0.57, corresponding to an OR of 1.77. In other words, for the median diagnostic test, a participant with a positive prediction from RarePT had 77% higher odds of having an abnormal test result. In 100 random permutations of phecode-laboratory test relationships, no permutation showed as many Bonferroni-significant associations (p < 0.01) (**Figure 3a, Supplementary Table S7-S8**). We additionally performed linear regression for each of these relationships, testing for a relationship between the model prediction and the quantitative test result. 43 of these regressions, representing 57% of these relationships, had a result that was in the expected direction and statistically significant, and 38 (51%) remained significant after Bonferroni correction. Again, 0 out of 100 random permutations showed as many Bonferroni-significant associations (p < 0.01) (**Figure 3a, Supplementary Table S7-8**).

**Figure 3.**
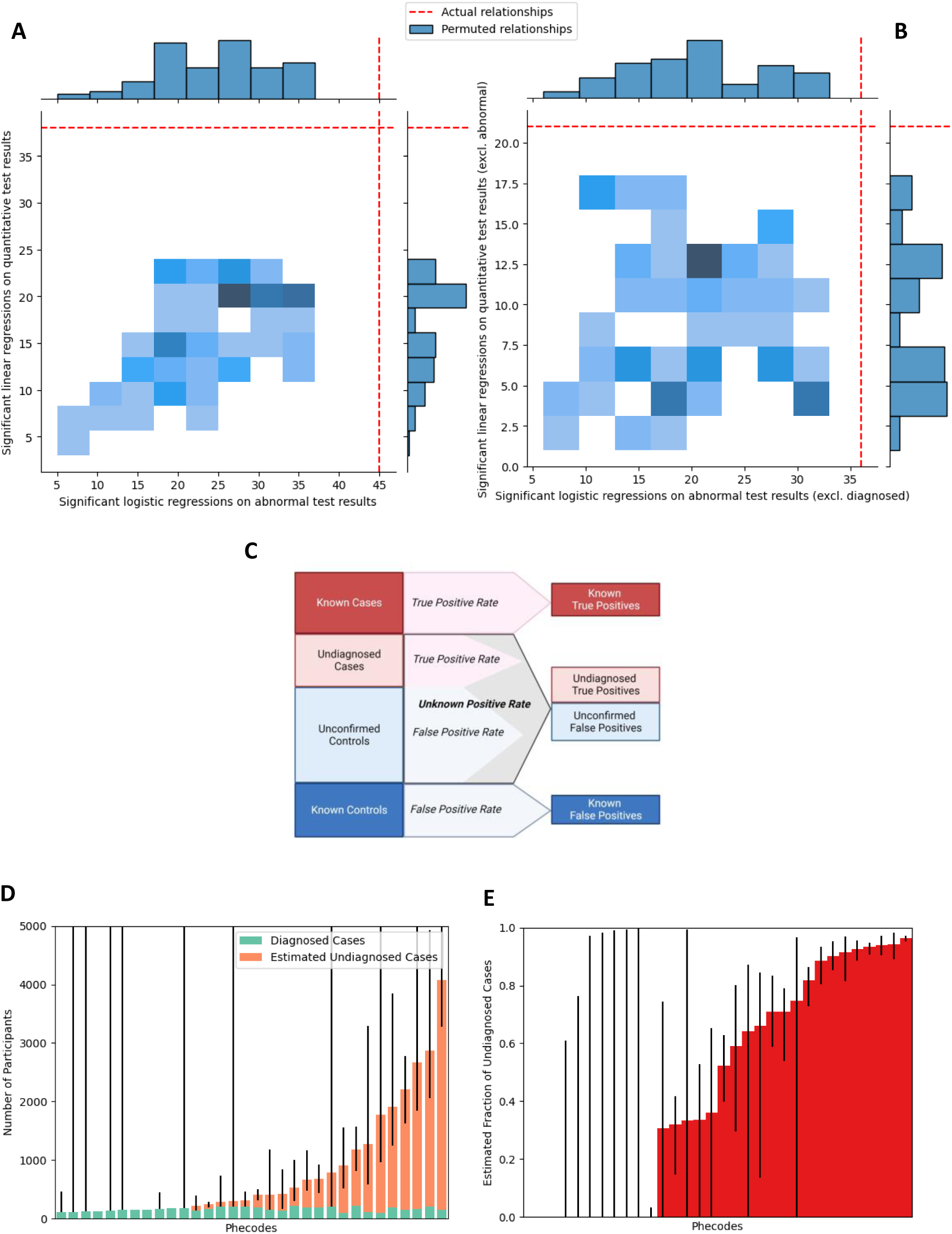
Estimated performance of RarePT to predict undiagnosed cases of 32 rare phecodes with available relevant laboratory tests. *(a-b) Regression analysis showing RarePT predicts diagnostic test results in UK Biobank participants.* Plots show number of significant regressions after Bonferroni correction for 75 tests in known relevant diagnostic tests (red dashed lines) and 100 random permutations of test-disease relationships (blue boxes). X axis shows logistic regression of abnormal test results vs. RarePT prediction, while Y axis shows linear regression of quantitative test results vs. RarePT prediction, both controlling for age at recruitment, sex, and self-reported ethnicity. Panel a shows results among all UK Biobank participants; Panel b excludes known cases, and the Y axis of panel b additionally excludes participants with abnormal test results. For both analyses, the number of significant regressions were lower than the actual observed results for all 100 permutations in both analyses. This demonstrates that RarePT predicts relevant clinical features of disease, not only the presence of a diagnosis. Individuals with a positive prediction from RarePT but no diagnosis likely have phenotypic profiles that are similar to the predicted rare diagnosis, and may include undiagnosed cases. *(c) Illustration of estimation of undiagnosed cases.* All individuals can be classified as known cases with EHR diagnosis, known controls with confirmed normal test results (defined as participants within 1 standard deviation of the population mean for all tests associated with a given phecode), or unknown. Unknown individuals are a mixture of undiagnosed cases and unconfirmed controls in an unknown proportion. We assume that the behavior of the model on unknown individuals (“unknown positive rate”) is a mixture of its behavior on cases (“true positive rate”) and controls (“false positive rate”). With this assumption, we estimate the proportion of cases and controls in the unknown group based on the relationship between these three positive prediction rates. See Supplementary Note 1 for discussion and derivation of this relationship. *(d-e) Number of undiagnosed cases and fraction of cases undiagnosed for 32 rare phecodes, estimated from RarePT’s performance on known cases and controls.* Error bars represent bootstrap 95% confidence intervals. While the presence of undiagnosed cases detected by RarePT varies across different rare phecodes, a majority of phecodes tested have an estimate above zero, and half are estimated to have a higher number of undiagnosed cases than diagnosed cases. This highlights the importance of developing methods to screen for undiagnosed cases of rare disease.

Taken together, these analyses demonstrate that positive predictions from RarePT do not merely predict diagnosis codes for rare diseases, but also capture clinically and biologically relevant features relevant to the diagnoses and to health outcomes more generally.

### Disease predictions suggest high rates of underdiagnosis for rare diseases

It has been demonstrated for many diseases, both rare and common, that only a fraction of affected individuals actually have a diagnosis annotated in their EHR^37–43^. As a result, it is likely that many of the participants annotated as controls in our dataset are actually undiagnosed cases. In order to evaluate RarePT’s performance in these undiagnosed cases, we repeated the regression analyses of mortality and estimated DALY restricting to participants labelled as controls, so that participants who had the corresponding diagnosis in their EHR were excluded. The mortality analysis produced similar results after excluding known diagnosed cases: 101 phecodes (67% of phecodes tested) had a significant association (p < 0.05) with increased mortality, of which 93 (60%) remained significant after Bonferroni correction for 155 phecodes (p < 0.00032). The median regression coefficient for the proportional hazard regression on mortality was 0.86, corresponding to a hazard ratio of 2.4. The DALY analysis also produced similar results: 114 phecodes (74% of phecodes tested) had a significant association (p < 0.05) with increased estimated DALY, and 106 (68%) remained significant after Bonferroni correction for 155 phecodes (p < 0.00032). 131 phecodes (85%) had a significant association with increased estimated YLD individually, 126 (81%) after Bonferroni correction; 110 phecodes (71%) had a significant association with increased estimated YLL individually, 104 (67%) after Bonferroni correction. For the median phecode, a positive prediction was associated with an increase in DALY of 1.5 years in controls. These results demonstrate that RarePT predictions are associated with health outcomes even when a diagnosis is not present in the EHR, suggesting that RarePT identifies clinically relevant features even in undiagnosed individuals and may be identifying undiagnosed cases.

We next repeated the logistic regression analysis testing RarePT predictions against abnormal test results. As expected, excluding known cases reduced the significance of many, but not all, of these regressions. Nevertheless, 47 of these regressions, representing 63% of these relationships, had a result that was in the expected direction and statistically significant (p < 0.05), and 36 (48%) remained significant after Bonferroni correction for 75 regressions (p < 0.00067). The median regression coefficient was 0.45, corresponding to an OR of 1.57. As with the regressions that included cases, in 100 random permutations of phecode-laboratory test relationships, no permutation showed as many Bonferroni-significant associations (p < 0.01) (**Figure 3b, Supplementary Tables S10-S11)**. We also repeated the linear regression analysis testing RarePT predictions against quantitative test results, excluding both known cases and participants with abnormal test results. 29 of these regressions, representing 39% of these relationships, had a result that was in the expected direction and statistically significant, with 21 (28%) remaining significant after Bonferroni correction. Again, 0 out of 100 random permutations showed as many Bonferroni-significant associations (**Figure 3b, Supplementary Tables S10-S11**). This analysis supports the conclusion that RarePT’s predictions are predictive not only of existing rare diagnoses, but also of undiagnosed cases.

In order to estimate the number of these undiagnosed cases that exist in the UK Biobank dataset, we first identified participants whose test results show that they are unlikely to be undiagnosed cases for a particular phecode. We defined this category of “confirmed controls” as participants whose test results fell within 1 standard deviation of the population mean for a particular test. This is possible because these tests were administered in an unbiased way to a large cross-section of participants, and the presence of a negative test result does not indicate that a physician ordered the test to rule out a suspected diagnosis. We then measured RarePT’s performance based on these confirmed controls and the observed diagnosed cases. Assuming that RarePT performs similarly for unconfirmed controls and undiagnosed cases as for confirmed controls and diagnosed cases, the prevalence of undiagnosed cases can be estimated by comparing the expected number of false positives among unconfirmed controls to the actual number of unconfirmed controls predicted as cases (**Figure 3c, Supplementary Note 1**).

We estimated the number of undiagnosed cases and the fraction of actual cases that are undiagnosed for each rare phecode with an associated diagnostic test, using a bootstrap sampling procedure to obtain 95% confidence intervals (**Figure 3d-e, Supplementary Table S12**). The estimated proportion of undiagnosed cases varied widely by phecode, but nearly three-quarters of phecodes tested (23/32 = 72%) had an estimate greater than 20%. Even more remarkably, nearly two-thirds of phecodes tested (20/32 = 63%) had more undiagnosed cases than diagnosed cases, and over a third (12/32 = 38%) had a bootstrap confidence interval entirely above the number of diagnosed cases. The median estimated rate of underdiagnosis across all phecodes tested was 83%, meaning that we estimate 83% of cases are undiagnosed for the median rare phecode. This analysis suggests that there are a very large number of undiagnosed cases of rare diseases in large population biobanks like the UK Biobank. Furthermore, it suggests that RarePT is able to predict some of these hidden undiagnosed cases, allowing them to be identified for the first time.

## Discussion

Here we present RarePT, a transformer-based phenotype prediction method designed to predict rare disease diagnoses based on diagnosis codes present in a patient’s electronic health records (EHR). We apply this method to predicting rare disease in the UK Biobank, and find that a very large fraction of rare disease cases are undiagnosed. Our method adds to a growing collection of phenotype prediction methods that use machine learning to clean and extend EHR data for downstream analysis ^44–48^. Our method is distinct from other approaches in that it focuses specifically on rare disease. It is typically difficult to train machine learning approaches for rare disease because the low prevalence of these diseases limits the availability of training data. We overcame this difficulty using a “masked diagnosis modeling” approach inspired by the approaches used to train AI language models such as BERT^19^. This approach learns about diagnoses by identifying which other diagnoses are most likely to appear in similar contexts, allowing it to learn informative features even for rare diagnoses. In addition to this training strategy, we reweighted our training data to give equal importance to rare and common diagnoses, boosting our power to predict rare diseases.

The trained RarePT model is highly predictive of a wide range of rare disease diagnoses, showing the promise of our deep learning approach as a screening test for specific rare diagnoses that could be applied in a clinical setting in the future. Across all rare phecodes, RarePT’s predictions are associated with a median diagnostic odds ratio (OR) of 48.0 in cross-validation – that is, participants predicted to have a rare diagnosis by our model are 48.0 times more likely to have that diagnosis in their EHR compared to participants without such a prediction. For some specific rare diseases, this performance is even better, with the top 10% of diagnoses achieving a diagnostic OR over 350 in cross-validation and the top 5% achieving a diagnostic OR over 1,500 in cross-validation. These values compare favorably to many diagnostic tests currently in standard clinical use. Remarkably, this performance is replicated in a completely independent cohort of patients at the Mount Sinai Health System in New York, which represents not only a different health system but an entirely different country with different medical practices and different standards for diagnostic coding and billing. The ability to predict rare disease diagnoses in this independent cohort shows the power and transferability of this approach.

In addition to successfully predicting rare diagnoses in participants’ EHR, RarePT also provides new evidence that a substantial number of participants may suffer from rare diseases without a diagnosis appearing in the EHR. This reinforces the known fact that many diagnoses are missing from EHR, due to biases in diagnosis, inconsistent use of billing codes, incomplete or fragmented patient records, and other issues^49–53^. This effect has previously been quantified for a variety of diseases, both common and rare, and often found to be substantial. For example, biobank studies have estimated that up to 75% of patients with erythropoetic protoporphyria (EPP)^39^, approximately 85% of patients with familial hypercholesterolemia^54^, and approximately 90% of patients with glycated hemoglobin (HbA_1c_) levels indicating diabetes^42^ remain undiagnosed. This is especially problematic for rare diseases, due to the known difficulty of correctly diagnosing rare diseases and the long diagnostic odyssey experienced by many rare disease patients^3–6,55^. We hypothesized that RarePT would correctly predict many of these undiagnosed cases, causing them to appear as false positives despite actually being correct predictions. For rare diseases where relevant biomarkers were available, these biomarkers consistently showed a significant excess of abnormal values and more extreme values within the normal range in individuals predicted positive by RarePT, supporting the hypothesis that many of RarePT’s predictions are actually undiagnosed cases. Consistent with literature estimates for common diseases, we estimate the prevalence of undiagnosed cases in rare diseases to be remarkably high: 72% of phecodes we tested appeared to have an underdiagnosis rate above 20%, and 63% of phecodes we tested were consistent with a majority of cases being undiagnosed. While these numbers may be higher than the true rate in the general population due to the UK Biobank being biased towards healthier participants, who are less likely to seek out and receive diagnoses than the general population^26,27,31^, both the existence and magnitude of this phenomenon are consistent with previous results on underdiagnosis of diseases in EHR. The RarePT model allows us to measure this underdiagnosis systematically across a range of rare diseases, which has not previously been possible, as well as to identify specific individuals who may be suffering from rare disease and have a missing or incorrect diagnosis.

There are many potential practical applications for RarePT. One of these is as a phenotype imputation step in a data preprocessing pipeline for downstream bioinformatic analysis, which has previously been unavailable for rare diagnoses^31,56,57^. Another application is in collecting rare disease cohorts for research studies or drug trials. Due to the rarity of rare diseases, identifying multiple patients with the same disease is a pressing problem in rare disease research, and the international research community has developed several tools to address it^7–9^. RarePT can augment or support these tools by allowing researchers to rapidly search EHR for patients who are likely to have a particular disease or patients who are phenotypically similar to another specific patient. It also has the potential to be developed into a clinical screening test for rare diseases, particularly in patients with a specific risk factor such as family history of disease or a genetic risk allele.

There are several limitations and areas of further development for this approach. First, phecodes are designed for phenome-wide association (PheWAS) studies primarily targeting common phenotypes and are not specifically designed to target rare diseases. While some specific rare phenotypes may be inaccessible to RarePT for this reason, recent studies have shown that vocabularies for common disease, including phecodes, do capture information about rare phenotypes^58–60^, and we identified phecodes that were rare in both the UK Biobank and MSDW cohorts. Future versions of this approach could increase the resolution for rare phenotypes by using phenotype ontologies designed to represent rare diseases, such as the Human Phenotype Ontology (HPO)^61^ or the OrphaNet disease ontology^62^. However, there are tradeoffs involved in this choice. Using a more fine-grained vocabulary for rare disease would dramatically increase the complexity of the model and its computational requirements. The analyses we present here require generation of millions of phenotype predictions, which took several hours of GPU time under the current RarePT architecture and would quickly become infeasible if the phenotype encoding used a complex hierarchical structure with a vocabulary many times larger. Choosing a phenotype encoding scheme that can more precisely capture rare phenotypes could also harm the model’s ability to capture information about rare and common disease in the same vocabulary, which may reduce the power of the model for rare disease and its transferability to other cohorts beyond its training set.

Second, the ICD-10 diagnosis codes we use to derive phecodes are known to be noisy and unreliable^52,63,64^. We have relied on established methods for automated phenome-wide phenotyping, many of which use diagnosis codes in spite of their limitations because more reliable sources of data are either difficult to access in an automated way or are not available phenome-wide^31,53^. This is particularly true for rare diseases, due to the difficulty of finding specialized experts who are capable of reviewing individual patient charts in detail to arrive at a confident diagnosis, particularly at scale^65–67^. In spite of this, the use of these automated phenotyping approaches could be a concern in our analysis of undiagnosed cases, since both our identification of participants with a disease diagnosis and our identification of patients with abnormal test results are based on these potentially unreliable automated procedures. It is possible that some of the supposedly undiagnosed cases we identified were actually diagnosed cases where the diagnosis escaped detection by our automated phenotyping process. It is also possible that the availability of certain test results and not others biased our analysis towards specific categories of phenotypes that are not representative of rare diseases in general. For example, blood disorders and metabolic disorders appear to be overrepresented among phenotypes with available diagnostic tests, while neoplasms and neurological disorders are entirely absent (**Supplementary Table S6)**. However, while diagnosis rates may differ by category, there is no reason to suppose that categories with greater availability of diagnostic tests are diagnosed at a lower rate than those with less availability of diagnostic tests. Indeed, if anything, the availability of simple diagnostic tests should increase the rate of diagnosis, making our estimates conservative. It is likely that we could make more reliable determinations of both diagnosis status and true phenotype by making use of other features available in the EHR, such as lab results, vitals, medications, or unstructured physician’s notes. We chose not to include these features in this analysis to avoid circularity in training and analysis, as the diagnoses contained in the EHR are informed by the lab results, vitals, and physician’s notes from the same EHR. Without careful insulation of these different modalities of data, any trained model or statistical analysis is likely to simply recapitulate the physician’s diagnostic criteria without gaining any predictive power for undiagnosed patients, a problem which RarePT avoids by excluding these redundant data sources. Previous studies have also identified undiagnosed cases using a longitudinal study design with direct physician involvement^40,41^. RarePT could facilitate this kind of analysis for specific diagnoses in future studies, following up on this broad automated analysis with in-depth analysis of individual diseases incorporating specific clinical expertise.

Finally, there are many opportunities to improve on our model architecture. Deep learning and AI is a rapidly evolving field, and the transformer-based architecture we used for this analysis may not be the optimal way to learn the semantic structure of EHR diagnoses. Recent studies have proposed new ways of derived phenotype embeddings, including extracting them from curated knowledge graphs or from general-purpose large language models pretrained on non-EHR data^68–70^. There are also a variety of approaches that have been used to process time series data from EHR in machine learning applications, including neural network models designed for time series data such as recurrent neural networks and using pretrained large language models to process EHR^71,72^. While incorporating newer and more sophisticated approaches may improve the model, they may also promote overfitting and reduce transferability of the model across health systems and datasets, as well as slowing down training and prediction. RarePT is both transferable and tractable, essential properties for a method designed to process large health system datasets.

In this paper we have shown that our deep learning phenotype prediction approach, RarePT, is capable of modeling and predicting rare disease diagnoses on a phenome-wide basis, with performance that compares favorably to diagnostic screening tests used in clinical settings. Remarkably, RarePT achieves this performance not only in held-out segments of the UK Biobank cohort it was trained on, but also on an entirely separate cohort of patients in the Mount Sinai Health System in New York City. This demonstrates that the predictive features RarePT uses are not specific to the UK Biobank, but are robust to differences in recruitment strategy, differences in race and ethnicity, and even differences in medical practices and billing procedures between countries. In addition to capturing specific diagnoses, RarePT predictions also predict clinical outcomes, including mortality, quality of life, and specific biomarkers associated with rare disease. Finally, we used predicted phenotypes from the model to estimate the prevalence of undiagnosed rare disease in the UK Biobank, showing that it is likely extremely high. This kind of systematic phenome-wide analysis has not previously been possible for rare diseases, highlighting the utility of RarePT to conduct large-scale studies on rare disease. The high rate of undiagnosed rare disease in large population datasets like the UK Biobank also highlights the need for new methods like RarePT to address the problem of undiagnosed rare disease and suggests a wide range of valuable clinical and research applications.

## Author Contributions

Dr. Jordan and Dr. Do had full access to all of the data in the study and take responsibility for the integrity of the data and accuracy of the data analysis.

*Concept and design:* Jordan, Do.

*Acquisition, analysis, or interpretation of the data:* Jordan, Vy, Do.

*Drafting of the manuscript:* Jordan, Do.

*Critical revision of the manuscript for important intellectual concept:* Jordan, Do.

*Statistical analysis:* Jordan.

*Administrative, technical, or material support:* Do

*Supervision:* Do.

## Materials and Correspondence

Requests for materials and correspondence should be addressed to Dr. Do.

## Data availability statement

Summary data required to generate figures will be deposited in a public repository prior to publication, and are available on request from the authors otherwise. Individual-level data from the UK Biobank and the Mount Sinai Data Warehouse are governed by third-party data use agreements and cannot be made available with this study. Researchers who qualify for access to deidentified data under the policies of the UK Biobank and/or the Mount Sinai Health System can access these data upon application to the respective institutions.

## Code availability statement

Code required to train all models, run all analyses, and generate all figures will be published in a public repository under an open-source license prior to publication, and is available upon request from the authors otherwise.

## Disclaimer

The content is solely the responsibility of the authors and does not necessarily represent the official views of the National Institutes of Health.

## Methods

### Data collection and preprocessing

The primary training data were derived from the UK Biobank^73^. For each participant, we retrieved age at recruitment (field 21022), sex (field 31), and a list of all ICD-10 diagnosis codes recorded across all inpatient hospital records (field 41270). We also retrieved self-reported ethnicity (field 21000), body mass index (BMI, field 21001), blood pressure (fields 4079-4080), LDL cholesterol (field 30780), total cholesterol (field 30690), blood glucose (field 30740), and glycated hemoglobin (HbA_1C_, field 30750) as baseline cohort characteristics (shown in **Table 1**), and 23 diagnostic tests and biomarkers assayed as part of the recruitment process (**Supplementary Table S6**), though these were not included in the data used to train the model. For this study, we represented each ICD-10 code as a binary indicator that could either be present or absent, ignoring the dates associated with each code. We mapped ICD-10 codes to version 1.2 of the Phecode classification, using the published mapping^24^. This resulted in a dataset of 436,407 participants, 239,711 female and 196,696 male, including 1,558 of the 1,570 phecodes with defined ICD-10 code mappings. Demographic and clinical characteristics of this cohort are shown in **Table 1**.

We constructed a balanced dataset of training examples consisting of 100 cases and 100 controls of each phecode, excluding phecodes with fewer than 100 cases or fewer than 100 controls. This excluded 273 phecodes, leaving 1,297 unique query phecodes. These 273 phecodes remained in the training data as diagnoses but were never used as the query in any training examples. Cases were defined as participants whose phecode diagnoses contained the query phecode. Controls were defined as participants whose phecode diagnoses did not include the query phecode or any phecodes listed as exclusions for the query phecode. For sex-specific phecodes, controls were also required to be the correct sex for the query phecode. For cases, the query phecode and all exclusion phecodes were removed from the diagnosis list as part of preprocessing (**Figure 1a**). Phecode diagnoses were encoded using a many-hot encoding, with phecodes ordered linearly by their numerical code; query phecodes were encoded using a one-hot encoding with the same ordering of phecodes.

Training examples were randomly split into five equal subsets for five-fold cross validation. Since the same individual can appear multiple times with different query phecodes, we required each cross-validation subset to contain a unique set of individuals, so that each individual can only appear in one subset. This prevents the model from improving performance by recognizing specific individuals from the training set and recapitulating the known diagnoses of those individuals. Cross-validation subsets were also required to contain similar numbers of cases and controls.

Our independent validation data were derived from the Mount Sinai Data Warehouse (MSDW), a database of clinical and operational data derived from the electronic health records (EHR) systems of the Mount Sinai Health System in New York City. These data are anonymized, standardized, and preprocessed for use in clinical and translational research. For each patient in this database, we retrieved age as of 2009 (the median date associated with the “age of recruitment” field in the UK Biobank), physician-reported sex, and a list of all ICD-10-CM diagnosis codes recorded in the EHR. We mapped these to phecodes using the published mapping for ICD-10-CM diagnosis codes, which is slightly different from the mapping for the ICD-10 codes used in the UK Biobank^24^. Patient records were processed into examples suitable for the model in the same way as described above. The final size of this cohort was 3,333,560. Demographic and clinical characteristics of this cohort are shown in **Table 1**.

Study protocols were approved by the Institutional Review Board at the Icahn School of Medicine at Mount Sinai (New York City, NY, USA; GCO#07–0529; STUDY-11–01139) and all participants provided informed consent. Use of data from the UK Biobank was approved with the UK Biobank Resource under application number 16218.

### Model architecture, tuning, and training

The model was implemented in Python using the Keras package^74^. **Figure 1a** shows a schematic of the model architecture. The input diagnosed phecodes feed into a stack of modified transformer decoder modules, based on the TransformerDecoder layer implemented in the KerasNLP package^75^. The standard TransformerDecoder layer was modified to remove the causal attention mask that prevents the self-attention layer from paying attention to positions that are later in the sequence than the token currently being considered. Since the phecode encoding is ordered by phenotype category and we are ignoring temporal sequencing, this causal mask would be inappropriate. Each decoder layer also contains a cross-attention layer which takes input from the encoded query phecode, allowing the model to learn attention relationships between the diagnosis phecodes and the query phecode. To adjust for demographic variables, the demographic variables are passed through a single densely connected layer to transform them into the same dimension as the phecodes, and then added to the output of the transformer layers and normalized. Finally, the prediction is given by a dot product between the input query phecodes and the demographics-adjusted output of the transformer layer, and transformed into a probability score using a softmax function. The Python code implementing this architecture will be made publicly available along with this publication.

Hyperparameter tuning was performed using the Hyperband algorithm, as implemented in the KerasTune package^25,76^. The list of hyperparameters and their final tuned values are found in **Supplementary Table S13**. We randomly sampled 80% of the training data to use for training and used the remaining 20% as the validation set for the hyperband algorithm, choosing the hyperparameters that minimized the training loss function on the validation set. For five-fold cross-validation runs, this 20% held out validation sample was contained within the training set and did not overlap the cross-validation test set. The hyperband algorithm was run for up to 18 epochs per model, stopping if validation loss failed to improve in 5 consecutive epochs. The final selected model was then trained for up to 54 epochs, and the best epoch was selected based on validation loss. Finally, after tuning was complete, the held-out validation set was added back into the training set and selected model was retrained for the selected number of epochs on the complete training set. Again, the Python code implementing this training procedure will be made publicly available along with this publication. We repeated this hyperparameter tuning for each of the five training subsets produced by cross-validation as well as on the full training set, and all six runs selected identical values for all hyperparameters. Models were trained using NVIDIA A100 GPUs on the Mount Sinai local high-performance computing cluster, Minerva. Each cross-validation run took approximately 5 hours of GPU time to tune and train, and the full model took approximately 6 hours, for a total of approximately 31 hours.

### Prediction of rare phenotypes

We calculated the prevalence of each phecode in the UK Biobank by dividing the number of cases by the total number of participants. We identified 155 rare phecodes with prevalence less than 1 in 2,000, or 0.05%, corresponding to the European Union definition of a rare disease. We calculated model predictions from each of the six trained models (five cross-validation models and one full-dataset model) using each of these 155 phecodes as a query for 436,407 participants in the UK Biobank, excluding from each model the participants contained in its own training set. We additionally produced predictions from the full trained model for 3,333,560 patients in the MSDW cohort for each of the 151 rare phecodes that were also present in that cohort. Generating model predictions for the UK Biobank cohort took approximately 3 hours of GPU time for each of the six models, for a total of approximately 18 hours; generating model predictions for the MSDW cohort took approximately 45 hours of GPU time. The MSDW cohort took substantially longer because the dataset was too large for the model to fit into memory and had to be broken up into batches.

We quantified the performance of our models by diagnostic odds ratios and positive predictive values. We arbitrarily chose a threshold probability score of 0.95 to represent a relatively high-confidence case prediction, and treated predictions with probability score > 0.95 as predicted cases and ≤ 0.95 as predicted controls. We quantified performance using odds ratio (OR) and positive predictive value (PPV), as these are measures relevant to diagnostic screening tests^29^. We calculated OR as

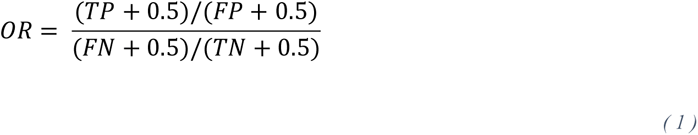

where *TP* is the number of true positive predictions (cases correctly predicted as cases), *FP* is the number of false positive predictions (controls incorrectly predicted as cases), *TN* is the number of true negative predictions (controls correctly predicted as controls), and *FN* is the number of false negative cases. In other words, this is the ratio between the odds of a positive prediction being a case and the odds of a negative prediction being a case. We added a correction of 0.5 to each count to correct for zeros^77^. We calculated PPV as

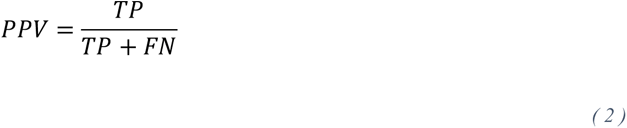

In other words, this is the probability that a positive prediction is a case. In all instances, we excluded controls with an exclusion phecode, controls whose sex did not match the phecode, and all individuals who were included in the training set of the cross-validated models.

### Mortality and DALY analyses

We estimated disability-adjusted life years (DALY) and its components years lost to disability (YLD) and years of life lost (YLL) for UKBB individuals using per-disease estimates from the 2019 Global Burden of Disease (GBD) study. We used the 80 non-overlapping non-communicable diseases that account for the majority of a population’s DALY as described by Jukarainen et al.^32,33^ GBD definitions of specific diseases and conditions were used to label individuals affected by these diseases in the UK Biobank. Estimates of disease burden in the UK from GBD were then applied to individuals with each disease to produce estimated values of DALY, YLD, and YLL.^32^ These estimated values were tested against RarePT predictions by linear regression. We retrieved a single prediction score by using the model trained on the full dataset for individuals who were not included in the training set, and the appropriate cross-validation model for individuals who were included in the training set (that is, the cross-validation model whose training set did not include that individual). We turned this score into a binary prediction using an arbitrary threshold of 0.95. We then performed linear regression testing the ability of this score (independent variable) to predict DALY, YLD, or YLL (dependent variable), controlling for age, sex, and self-reported ethnicity. We repeated this analysis both including all UK Biobank participants and excluding known diagnosed cases and exclusions for each phecode.

We additionally retrieved date of death and date of recruitment from the UK Biobank (fields 40000 and 53) and performed an analysis of mortality using Cox proportional hazard regression. We treated time from recruitment to death as a right-censored dependent variable, again using the binary RarePT prediction as an independent variable along with age at recruitment, sex, and self-reported ethnicity. As with DALY, we repeated this analysis both including all UK Biobank participants and excluding known diagnosed cases and exclusions for each phecode. All regressions were performed in Python using the statsmodels package^78^.

### Biomarker and diagnostic test analysis

We collected biomarkers and diagnostic tests associated with phecodes using the SNOMED-CT database of clinical terms^34,35^. We identified all ICD-10 codes that mapped to any of our 155 rare phecodes and also mapped to a SNOMED-CT term with an “interprets” relationship (concept 363714003). The “interprets” relationship indicates that the concept represented by the diagnosis code has an underlying evaluation that is “intrinsic to the meaning of” that concept^79^. Examples of this kind of relationship include the relationship between obesity and measured body weight, hypercholesterolemia and total serum cholesterol, or thrombocytopenia and platelet count. In most cases, SNOMED-CT also identifies the direction of the relationship using the “has interpretation” relationship (concept 363713009). For example, hypercholesterolemia is interpreted as total serum cholesterol above reference range, while thrombocytopenia is interpreted as platelet count below reference range. For each concept that was the target of an “interprets” relationship, we manually searched for a corresponding measurement available in the UK Biobank and a corresponding reference range. The result was a list of 75 relationships between rare phecodes and UK Biobank data fields, encompassing 32 rare phecodes and 23 data fields, each with an expected direction of relationship (above, below, or outside) and sex-specific reference ranges **(Supplementary Table S6)**.

As with the previously described regression analyses, we retrieved a single prediction score for each of these 32 rare phecodes by using the model trained on the full dataset for individuals who were not included in the training set, and the appropriate cross-validation model for individuals who were included in the training set (that is, the cross-validation model whose training set did not include that individual). We turned this score into a binary prediction using the same 0.95 threshold. We then performed two regression analyses testing the ability of this binary prediction (independent variable) to predict the corresponding data field (dependent variable), controlling for age, sex, and self-reported ethnicity. In the first analysis, we used the expected direction of relationship and the reference range to construct a binary variable indicating whether each individual had an abnormal result in the direction expected. We performed logistic regression using this binary variable as the dependent variable. We repeated this analysis both including all participants and excluding individuals labelled as cases or exclusions for each phecode. This regression tests whether the model can predict individuals with abnormal test results consistent with a diagnosis even in individuals labelled as controls. In the second analysis, we normalized the values of each biomarker within the reference range so that the sample population for each sex had mean 0 and variance 1 after excluding all individuals with values outside the reference range. Finally, we aligned the values so that the expected direction of association was always positive, by multiplying them by -1 for “below reference range” relationships and taking their absolute value for “outside reference range” relationships. We performed standard linear regression using this normalized and aligned value as the dependent variable. We repeated this analysis both including all participants and excluding both individuals labelled as cases or exclusions and individuals with abnormal test results. This regression tests whether the model can predict individuals with elevated or reduced results even if they are still within the normal range. Regressions were performed in Python using the statsmodels package^78^.

Finally, we identified a set of “confirmed controls” for each phecode, defined as individuals labelled as controls who also had all associated results within the reference range and within one standard deviation of the population mean for their sex. We consider these individuals very unlikely to be undiagnosed cases incorrectly labelled as controls. We used the performance of our model on these confirmed controls to estimate the false positive rate of our model for each of the 32 phecodes with available relationships to test results. We then used this false positive rate to estimate the number of undiagnosed cases using the following relationship:

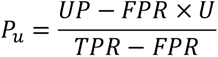

Where *P_u_* represents the number of undiagnosed cases; *U* represents the total number of individuals with unknown case-control status, excluding controls confirmed by laboratory tests but including undiagnosed cases; *UP* represents the total number of unknowns predicted as cases by the model, again excluding controls confirmed by laboratory tests but including undiagnosed cases; *FPR* represents the false positive rate of the model as estimated from confirmed controls; and *TPR* represents the true positive rate of the model as estimated from diagnosed cases. See **Supplementary Note 1** for derivation and discussion of this relationship.

The Python code implementing all these analyses will be made publicly available along with this publication.

### Software Package and Workflow

For portability and reproducibility, model training and analysis code is formatted as a Snakemake workflow.^80^ This allows easy retraining of the RarePT model and reproduction of the analyses reported here on any appropriately-formatted individual-level dataset. After creating an appropriately named and formatted input data file and setting up Snakemake for their execution environment, users can train a new model with a single command:

snakemake results/models/my_dataset.100_case_control_sample.full_from_5_fold_cv.seed_18

The number of cases and controls sampled, the number of cross-validation folds used for testing, and the random seed can be changed by changing the appropriate values in the targeted filename. Likewise,

snakemake results/data/my_dataset.100_case_control_sample.5_fold_cv.seed_18.all_rare_predictions_with_cv.parquet

trains a model and uses cross-validation to generate model predictions for all rare phecodes, and

snakemake results/data/uk_biobank.100_case_control_sample.full_from_5_fold_cv.seed_18.vs.my_dataset.all_rare_predictions.parquet

uses the UK Biobank trained model to generate model predictions for all rare phecodes in a user dataset. Snakemake can be configured for many different high-performance computing and cloud computing environments and, when properly configured, automatically manages resource requirements and package dependencies.

The Snakemake workflow will be published with acceptance of this manuscript in a peer-reviewed journal. Prior to formal publication, it is available on request from the authors.

## List of Supplementary Items

**Supplementary Note S1.** Estimation of undiagnosed cases.

**Supplementary Figure S1.** Receiver operating characteristic (ROC) curves for training and testing cross-validation datasets in UK Biobank.

**Supplementary Figure S2.** Positive predictive value of RarePT predictions for 155 rare phecodes in UK Biobank and MSDW cohorts.

**Supplementary Table S1.** Cross-validation performance metrics for training and testing.

**Supplementary Table S2.** List of 155 rare phecodes in UK Biobank and Mount Sinai Data Warehouse.

**Supplementary Table S3.** Model performance by phecode for 155 rare phecodes in UK Biobank cohort.

**Supplementary Table S4.** Model performance by phecode for 151 rare phecodes in MSDW cohort.

**Supplementary Table S5.** Regression results for mortality, DALY, and related measures.

**Supplementary Table S6.** List of diagnostic tests relevant to rare phecodes.

**Supplementary Table S7.** Regression results for 75 diagnostic tests known to be relevant to rare phecodes.

**Supplementary Table S8.** Regression results for 100 permutations of phecode-test relationships.

**Supplementary Table S9.** Regression results for mortality, DALY, and related measures in controls only.

**Supplementary Table S10.** Regression results for 75 diagnostic tests known to be relevant to rare phecodes in controls only.

**Supplementary Table S11.** Regression results for 100 permutations of phecode-test relationships in controls only.

**Supplementary Table S12.** Estimated numbers of undiagnosed cases for 32 rare phecodes.

**Supplementary Table S13.** Description of model hyperparameters and final tuned values.

