## Supplementary Materials for "A deep learning transformer model predicts high rates of undiagnosed rare disease in large electronic health systems"

**Supplementary Note: Estimation of undiagnosed cases**

For a given diagnosis code, we can classify all participants into four categories:

1. Confirmed controls, meaning people who should not have the diagnosis and whose EHR correctly does not include the diagnosis, and who additionally appear normal in appropriate laboratory tests (defined as within 1 standard deviation of the population mean for their sex);
2. Unconfirmed controls, meaning people who should not have the diagnosis and whose EHR correctly does not include the diagnosis, but who cannot be labelled as “confirmed controls” based on available laboratory tests;
3. Undiagnosed cases, meaning people who should have the diagnosis but whose EHR incorrectly fails to include the diagnosis;
4. Diagnosed cases, meaning people who should have the diagnosis and whose EHR correctly includes the diagnosis.

(In principle, there are also two other possible categories:

5. Misdiagnosed cases, meaning people who should not have the diagnosis but whose EHR incorrectly includes the diagnosis;
6. Incorrectly confirmed controls, meaning people who should have the diagnosis but whose EHR incorrectly does not include the diagnosis, but additionally incorrectly appear normal according to laboratory tests.

We are ignoring these two categories for this analysis, because we assume they are small and we have no reliable way to estimate their actual size.)

We can represent the model’s performance on these categories using a 2 × 4 contingency table, rather than a normal 2 × 2 contingency table:

|  | Predicted Controls | Predicted Cases |
| --- | --- | --- |
| Confirmed Controls  $N$ | Confirmed True Negatives  $TN$ | Confirmed False Positives  $FP$ |
| Unconfirmed Controls  $N_{u}$ | Unconfirmed True Negatives  $TN_{u}$ | Unconfirmed False Positives  $FP_{u}$ |
| Undiagnosed Cases  $P_{u}$ | Undiagnosed False Negatives  $FN_{u}$ | Undiagnosed True Positives  $TP_{u}$ |
| Diagnosed Cases  $P$ | Diagnosed False Negatives  $FN$ | Diagnosed True Positives  $TP$ |

In practice, we cannot tell the difference between unconfirmed controls and undiagnosed cases, with both being collapsed into a single “unknown” category, causing this to collapse into a 2 × 3 table rather than a 2 × 4 table:

|  | Predicted Controls | Predicted Cases |
| --- | --- | --- |
| Confirmed Controls  $N$ | Confirmed True Negatives  $TN$ | Confirmed False Positives  $FP$ |
| Unknown  $U= N_{u}+ P_{u}$ | Unknown Negatives  $UN= TN_{u}+ FN_{u}$ | Unknown Positives  $UP= FP_{u}+ TP_{u}$ |
| Diagnosed Cases  $P$ | Diagnosed False Negatives  $FN$ | Diagnosed True Positives  $TP$ |

However, despite the fact that we cannot tell these two categories apart, we do expect them to behave differently. Since the model does not have access to the laboratory results, we can assume that it cannot tell the difference between confirmed and unconfirmed controls. In particular, we assume that the false positive rate (FPR) of the model should be the same for confirmed and unconfirmed controls:

$$FPR=\frac{FP}{N}=\frac{FP_{u}}{N_{u}}$$

*(1)*

Likewise, since we mask diagnosis status for the query diagnosis code from the model, we can also assume that the model cannot tell the difference between diagnosed and undiagnosed cases, and in particular that the true positive rate (TPR) of the model should be the same for diagnosed and undiagnosed cases:

$$TPR=\frac{TP}{P}=\frac{TP_{u}}{P_{u}}$$

*(2)*

Intuitively, since unknown individuals are a mixture of undiagnosed cases and unconfirmed controls, we expect the number of unknown positives to be produced by a weighted average of the FPR and TPR, weighted by the relative proportion of undiagnosed cases and unconfirmed controls. Since we can calculate the FPR of the model from confirmed controls and the TPR of the model from diagnosed cases, and we can observe directly the number of unknown positives, we can use these quantities to calculate this mixture proportion.

To derive this relationship explicitly, we can express the number of unknown positives as:

$$UP=TP_{u}+FP_{u}=TPR\times P_{u}+FPR\times N_{u}$$

*(3)*

The quantity of interest is the number of undiagnosed cases $P_{u}$. We can express $N_{u}$ in terms of $P_{u}$ by observing that:

$$N_{u}=U-P_{u}$$

*(4)*

Substituting this into equation 3 yields:

$$UP=TPR\times P_{u}+FPR\times(U-P_{u})$$

*(5)*

And solving for $P_{u}$ gives:

$$P_{u}=\frac{UP-FPR\times U}{TPR-FPR}$$

*(6)*

$UP$ and $U$are directly observable, $TPR$ can be calculated from $TP$ and $P$ using equation 1, and $FPR$ can be calculated from $FP$ and $N$ using equation 2. Equation 6 therefore allows us to estimate the number of undiagnosed cases for any diagnosis code given a working predictive model and a diagnostic laboratory test.

**Supplementary Figure S1. Receiver operating characteristic (ROC) curves for training and testing cross-validation datasets in UK Biobank.** Thin lines represent 5-fold cross-validation runs, thick lines represent the median of cross-validation.

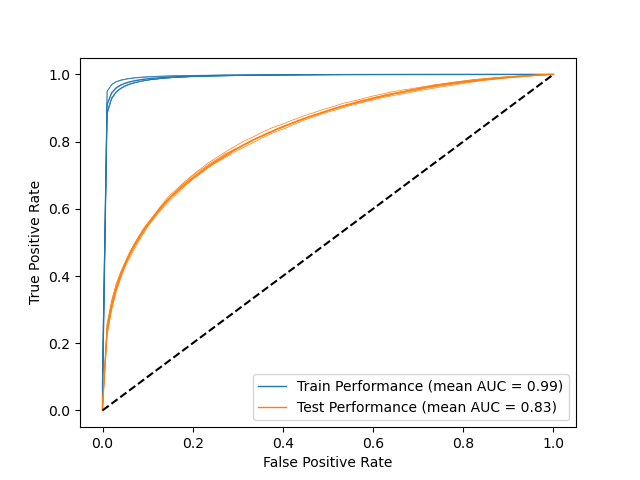

**Supplementary Figure S2. Positive predictive value of RarePT predictions for 155 rare phecodes in UK Biobank and MSDW cohorts.** The five blue boxes labelled “UKBB/CV1-5” show the performance of the five models trained using five-fold cross-validation within the UK Biobank, excluding each model’s training set. The orange box labeled “MSDW” shows the performance of the model trained using the full UK Biobank dataset on the Mount Sinai Data Warehouse dataset, an independent cohort from the Mount Sinai Health System in New York. Boxes show 1^st^ quartile, median, and 3^rd^ quartile; whiskers extend to 80% of interquartile range.

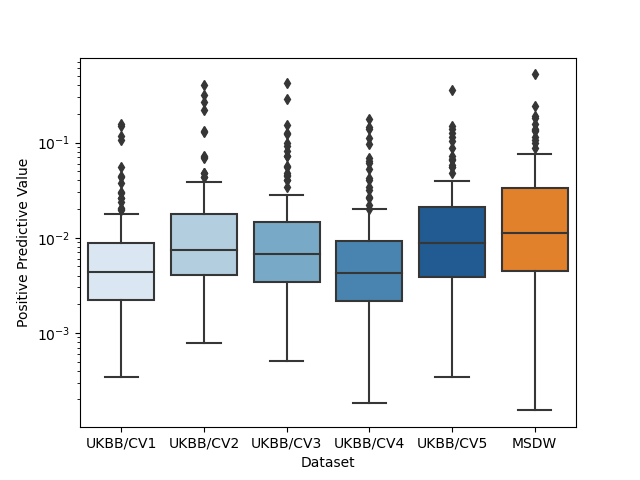

**Supplementary Table S1. Cross-validation performance metrics for training and testing.**

| **Cross-Validation split** | **Cross-entropy training loss** | | **Area Under Receiver Operating Characteristic Curve** | | **Accuracy on balanced dataset (at 0.5 threshold)** | | **Precision on balanced dataset (at 0.5 threshold)** | | **Recall on balanced dataset (at 0.5 threshold)** | |
| --- | --- | --- | --- | --- | --- | --- | --- | --- | --- | --- |
|  | **Train** | **Test** | **Train** | **Test** | **Train** | **Test** | **Train** | **Test** | **Train** | **Test** |
| 1 | 0.29 | 0.53 | 0.97 | 0.82 | 0.90 | 0.74 | 0.91 | 0.71 | 0.89 | 0.80 |
| 2 | 0.36 | 0.51 | 0.94 | 0.82 | 0.86 | 0.74 | 0.87 | 0.73 | 0.85 | 0.76 |
| 3 | 0.36 | 0.52 | 0.94 | 0.83 | 0.87 | 0.74 | 0.88 | 0.71 | 0.86 | 0.79 |
| 4 | 0.29 | 0.51 | 0.97 | 0.83 | 0.90 | 0.75 | 0.91 | 0.73 | 0.89 | 0.78 |
| 5 | 0.36 | 0.51 | 0.94 | 0.83 | 0.87 | 0.75 | 0.87 | 0.74 | 0.85 | 0.76 |
| Full | 0.36 | N/A | 0.94 | N/A | 0.86 | N/A | 0.87 | N/A | 0.84 | N/A |

**Supplementary Table S2. List of 155 rare phecodes in UK Biobank and Mount Sinai Data Warehouse.**

| **Phecode** | **Phenotype** | **UK Biobank** | | | **Mount Sinai Data Warehouse** | | |
| --- | --- | --- | --- | --- | --- | --- | --- |
|  |  | **Cases** | **Controls** | **Excluded** | **Cases** | **Controls** | **Excluded** |
| 008.7 | Intestinal infection due to protozoa | 102 | 410572 | 25733 | 634 | 3306223 | 29009 |
| 070.1 | Viral hepatitis A | 129 | 424544 | 11734 | 381 | 3093314 | 242171 |
| 071 | Human immunodeficiency virus [HIV] disease | 118 | 424544 | 11745 | 0 | 3093314 | 242552 |
| 071.1 | HIV infection, symptomatic | 210 | 424544 | 11653 | 0 | 3093314 | 242552 |
| 079.1 | Varicella infection | 122 | 424544 | 11741 | 1062 | 3093314 | 241490 |
| 079.2 | Infectious mononucleosis | 101 | 424544 | 11762 | 2708 | 3093314 | 239844 |
| 110.11 | Dermatophytosis of nail | 210 | 428476 | 7721 | 30624 | 3230997 | 74245 |
| 110.13 | Dermatophytosis of the body | 116 | 428476 | 7815 | 13897 | 3230997 | 90972 |
| 110.2 | Dermatomycoses | 152 | 428476 | 7779 | 13480 | 3230997 | 91389 |
| 112.3 | Candidiasis of skin and nails | 127 | 428476 | 7804 | 4865 | 3230997 | 100004 |
| 130 | Spirochetal infection | 161 | 434691 | 1555 | 6664 | 3315974 | 13228 |
| 145.1 | Cancer of lip | 102 | 432533 | 3772 | 91 | 3325779 | 9996 |
| 149 | Cancer of larynx, pharynx, nasal cavities | 128 | 432533 | 3746 | 503 | 3325779 | 9584 |
| 149.3 | Cancer of hypopharynx | 120 | 432533 | 3754 | 230 | 3325779 | 9857 |
| 149.9 | Cancer of of nasal cavities | 111 | 432533 | 3763 | 615 | 3325779 | 9472 |
| 164 | Cancer of intrathoracic organs | 121 | 429290 | 6996 | 364 | 3323454 | 12048 |
| 174.3 | Neoplasm of uncertain behavior of breast | 160 | 403379 | 32868 | 2557 | 3242769 | 90540 |
| 187.1 | Malignant neoplasm of unspecified male genital organ | 145 | 181744 | 254518 | 189 | 1438462 | 1897215 |
| 187.8 | Neoplasm of uncertain behavior of male genital organs | 167 | 181744 | 254496 | 233 | 1438462 | 1897171 |
| 189.12 | Malignant neoplasm of renal pelvis | 165 | 427877 | 8365 | 304 | 3321058 | 14504 |
| 191.1 | Cancer of brain and nervous system | 102 | 432667 | 3638 | 396 | 3320107 | 15363 |
| 194 | Cancer of other endocrine glands | 177 | 433017 | 3213 | 361 | 3313460 | 22045 |
| 199.4 | Neurofibromatosis | 162 | 347725 | 88520 | 855 | 3229541 | 105470 |
| 204.11 | Lymphoid leukemia, acute | 128 | 426531 | 9748 | 580 | 3308690 | 26596 |
| 204.3 | Monocytic leukemia | 116 | 426531 | 9760 | 184 | 3308690 | 26992 |
| 224 | Benign neoplasm of eye | 119 | 432667 | 3621 | 583 | 3320107 | 15176 |
| 224.1 | Benign neoplasm of eye, uveal | 215 | 432667 | 3525 | 3066 | 3320107 | 12693 |

**Supplementary Table S2 (cont).**

| **Phecode** | **Phenotype** | **UK Biobank** | | | **Mount Sinai Data Warehouse** | | |
| --- | --- | --- | --- | --- | --- | --- | --- |
|  |  | **Cases** | **Controls** | **Excluded** | **Cases** | **Controls** | **Excluded** |
| 225.2 | Benign neoplasm of spinal cord, meninges | 108 | 432667 | 3632 | 348 | 3320107 | 15411 |
| 242.2 | Toxic multinodular goiter | 218 | 400906 | 35283 | 587 | 3120722 | 214557 |
| 242.3 | Exophthalmos | 210 | 400906 | 35291 | 0 | 3120722 | 215144 |
| 246.7 | Abnormal results of function study of thyroid | 153 | 400906 | 35348 | 8767 | 3120722 | 206377 |
| 250.12 | Type 1 diabetes with renal manifestations | 167 | 391009 | 45231 | 1297 | 2993665 | 340904 |
| 250.3 | Insulin pump user | 133 | 391009 | 45265 | 1410 | 2993665 | 340791 |
| 250.5 | Glycosuria or Acetonuria | 181 | 391009 | 45217 | 1236 | 2993665 | 340965 |
| 253 | Disorders of the pituitary gland and its hypothalamic control | 214 | 426601 | 9592 | 2559 | 3243013 | 90294 |
| 253.11 | Acromegaly and gigantism | 152 | 426601 | 9654 | 520 | 3243013 | 92333 |
| 253.3 | Diabetes insipidus | 170 | 426601 | 9636 | 756 | 3243013 | 92097 |
| 255.11 | Cushing's syndrome | 204 | 426601 | 9602 | 1757 | 3243013 | 91096 |
| 255.12 | Hyperaldosteronism | 154 | 426601 | 9652 | 873 | 3243013 | 91980 |
| 258 | Iatrogenic endocrine disorders | 204 | 426601 | 9602 | 217 | 3243013 | 92636 |
| 261.41 | Rickets or osteomalacia | 137 | 424163 | 12107 | 1163 | 3075743 | 258960 |
| 270.35 | Macroglobulinemia | 176 | 434025 | 2206 | - | - | - |
| 272.12 | Hyperglyceridemia | 172 | 359522 | 76713 | 17131 | 2869662 | 449073 |
| 277.51 | Lipoprotein disorders | 121 | 430990 | 5296 | 4365 | 3298365 | 33136 |
| 278.3 | Localized adiposity | 111 | 400597 | 35699 | 2267 | 3042681 | 290918 |
| 278.4 | Abnormal weight gain | 142 | 400597 | 35668 | 27444 | 3042681 | 265741 |
| 281.9 | Deficiency anemias | 148 | 389688 | 46571 |  |  |  |
| 286.11 | Von willebrand's disease | 180 | 429459 | 6768 | 1321 | 3281246 | 53299 |
| 286.13 | Congenital factor VIII disorder | 117 | 429459 | 6831 | 619 | 3281246 | 54001 |
| 286.5 | Hemorrhagic disorder due to intrinsic circulating anticoagulants | 142 | 429459 | 6806 | 247 | 3281246 | 54373 |
| 288.3 | Eosinophilia | 211 | 418735 | 17461 | 2869 | 3239873 | 93124 |
| 289.9 | Abnormality of red blood cells | 188 | 418735 | 17484 | - | - | - |
| 290.3 | Other persistent mental disorders due to conditions classified elsewhere | 185 | 410889 | 25333 | 607 | 3256381 | 78878 |
| 297 | Suicidal ideation or attempt | 131 | 340879 | 95397 | 121 | 2935066 | 400679 |
| 300.4 | Dysthymic disorder | 105 | 340879 | 95423 | 7066 | 2935066 | 393734 |
| 305.2 | Eating disorder | 200 | 340879 | 95328 | 6514 | 2935066 | 394286 |
| 315.3 | Mental retardation | 196 | 434323 | 1888 | 3409 | 3263727 | 68730 |

**Supplementary Table S2 (cont).**

| **Phecode** | **Phenotype** | **UK Biobank** | | | **Mount Sinai Data Warehouse** | | |
| --- | --- | --- | --- | --- | --- | --- | --- |
|  |  | **Cases** | **Controls** | **Excluded** | **Cases** | **Controls** | **Excluded** |
| 323.2 | Acute (transverse) myelitis | 132 | 434356 | 1919 | 426 | 3330605 | 4835 |
| 325 | Phlebitis and thrombophlebitis of intracranial venous sinuses | 103 | 434356 | 1948 | 520 | 3330605 | 4741 |
| 337.1 | Peripheral autonomic neuropathy | 157 | 404867 | 31383 | 1185 | 3190040 | 144641 |
| 353.1 | Nerve plexus lesions | 214 | 404412 | 31781 | 3523 | 3184717 | 147626 |
| 353.2 | Nerve root lesions | 124 | 404412 | 31871 | 1920 | 3184717 | 149229 |
| 361.2 | Retinoschisis and retinal cysts | 200 | 402140 | 34067 | 320 | 3242049 | 93497 |
| 362.3 | Other nondiabetic retinopathy | 143 | 402107 | 34157 | 89 | 3236131 | 99646 |
| 364.41 | Keratoconus | 171 | 402140 | 34096 | 1482 | 3242049 | 92335 |
| 366.1 | Nonsenile Cataract | 118 | 375850 | 60439 | 398 | 3265196 | 70272 |
| 366.3 | Traumatic cataract | 160 | 375850 | 60397 | 182 | 3265196 | 70488 |
| 367.4 | Presbyopia | 123 | 423780 | 12504 | 11247 | 3275594 | 49025 |
| 377.1 | Optic atrophy | 163 | 414691 | 21553 | 2949 | 3228213 | 104704 |
| 378 | Strabismus and other disorders of binocular eye movements | 139 | 414691 | 21577 | 3838 | 3228213 | 103815 |
| 388 | Other disorders of ear | 179 | 421161 | 15067 | 9399 | 3228376 | 98091 |
| 394.4 | Acute rheumatic heart disease | 108 | 396181 | 40118 | 1231 | 3168532 | 166103 |
| 401.2 | Hypertensive heart and/or renal disease | 138 | 283976 | 152293 | 3052 | 2800522 | 532292 |
| 420.22 | Chronic pericarditis | 204 | 428210 | 7993 | 628 | 3290682 | 44556 |
| 425.2 | Secondary/extrinsic cardiomyopathies | 131 | 428210 | 8066 | 2455 | 3290682 | 42729 |
| 425.8 | Other cardiomyopathy | 108 | 428210 | 8089 | 98 | 3290682 | 45086 |
| 426.8 | Other cardiac conduction disorders | 155 | 368986 | 67266 | 1779 | 3043802 | 290285 |
| 441.2 | Chronic vascular insufficiency of intestine | 160 | 413569 | 22678 | 857 | 3193743 | 141266 |
| 446 | Polyarteritis nodosa and allied conditions | 174 | 413569 | 22664 | 727 | 3193743 | 141396 |
| 459.1 | Hemorrhage NOS | 200 | 381464 | 54743 | 5885 | 3261168 | 68813 |
| 473.1 | Chronic laryngitis | 132 | 403599 | 32676 | 6487 | 3013170 | 316209 |
| 500.1 | Extrinsic allergic alveolitis | 199 | 401112 | 35096 | 444 | 3263229 | 72193 |
| 519 | Other diseases of respiratory system, not elsewhere classified | 195 | 413328 | 22884 | 717 | 3302476 | 32673 |
| 523.1 | Gingivitis | 202 | 418498 | 17707 | 1319 | 3286000 | 48547 |

**Supplementary Table S2 (cont).**

| **Phecode** | **Phenotype** | **UK Biobank** | | | **Mount Sinai Data Warehouse** | | |
| --- | --- | --- | --- | --- | --- | --- | --- |
|  |  | **Cases** | **Controls** | **Excluded** | **Cases** | **Controls** | **Excluded** |
| 527.1 | Hypertrophy of salivary gland | 108 | 426096 | 10203 | 331 | 3294712 | 40823 |
| 527.7 | Disturbance of salivary secretion | 200 | 426096 | 10111 | 7616 | 3294712 | 33538 |
| 528.12 | Oral aphthae | 135 | 426096 | 10176 | 3777 | 3294712 | 37377 |
| 528.41 | Cyst of the salivary gland | 156 | 426096 | 10155 | 473 | 3294712 | 40681 |
| 573.2 | Liver replaced by transplant | 176 | 410586 | 25645 | 2534 | 3206773 | 126559 |
| 580.11 | Proliferative glomerulonephritis | 132 | 391999 | 44276 | 334 | 3173998 | 161534 |
| 588 | Disorders resulting from impaired renal function | 194 | 391999 | 44214 | - | - | - |
| 597.2 | Urinary complications NEC | 182 | 409473 | 26752 | 14 | 3291686 | 44166 |
| 609.2 | Abnormal spermatozoa | 207 | 157577 | 278623 | 2282 | 1308117 | 2025467 |
| 611.1 | Abnormal mammogram | 101 | 426266 | 10040 | 33610 | 3208817 | 93439 |
| 612.3 | Congenital anomalies of breast | 115 | 426266 | 10026 | 874 | 3208817 | 126175 |
| 613.5 | Mastodynia | 152 | 433705 | 2550 | 19904 | 3299405 | 16557 |
| 620 | Dysplasia of female genital organs | 130 | 221895 | 214382 | 641 | 1839784 | 1495441 |
| 626.11 | Absent or infrequent menstruation | 158 | 199354 | 236895 | 54429 | 1643765 | 1637672 |
| 626.4 | Premenstrual tension syndromes | 179 | 199354 | 236874 | 1924 | 1643765 | 1690177 |
| 643.1 | Hyperemesis gravidarum | 200 | 239398 | 196809 | 1732 | 1858070 | 1476064 |
| 647 | Infectious and parasitic complications affecting pregnancy | 126 | 239162 | 197119 | 1962 | 1858841 | 1475063 |
| 649 | Other conditions or status of the mother complicating pregnancy, childbirth, or the puerperium | 153 | 239255 | 196999 | 9388 | 1844165 | 1482313 |
| 656.2 | Respiratory conditions of fetus and newborn | 193 | 435484 | 730 | 4163 | 3264834 | 66869 |
| 686.5 | Pyoderma | 103 | 413801 | 22503 | 602 | 3259004 | 76260 |
| 690.1 | Seborrheic dermatitis | 200 | 421861 | 14346 | 86655 | 3140208 | 109003 |
| 691 | Congenital anomalies of skin | 150 | 421861 | 14396 | 2443 | 3140208 | 193215 |
| 694.1 | Vitiligo | 203 | 421861 | 14343 | 4577 | 3140208 | 191081 |
| 695.21 | Dermatitis herpetiformis | 110 | 421861 | 14436 | 157 | 3140208 | 195501 |
| 695.81 | Erythema nodosum | 129 | 421861 | 14417 | 583 | 3140208 | 195075 |
| 696.3 | Pityriasis | 108 | 421861 | 14438 | 4175 | 3140208 | 191483 |
| 701.1 | Keratoderma, acquired | 113 | 426667 | 9627 | 2427 | 3236199 | 97240 |
| 701.3 | Circumscribed scleroderma | 106 | 426667 | 9634 | 2721 | 3236199 | 96946 |

**Supplementary Table S2 (cont).**

| **Phecode** | **Phenotype** | **UK Biobank** | | | **Mount Sinai Data Warehouse** | | |
| --- | --- | --- | --- | --- | --- | --- | --- |
|  |  | **Cases** | **Controls** | **Excluded** | **Cases** | **Controls** | **Excluded** |
| 701.6 | Acquired acanthosis nigricans | 204 | 426667 | 9536 | 4504 | 3236199 | 95163 |
| 704.8 | Other specified diseases of hair and hair follicles | 143 | 426377 | 9887 | 10979 | 3243427 | 81460 |
| 706.1 | Acne | 145 | 422186 | 14076 | 70890 | 3186649 | 78327 |
| 709.5 | Dermatomyositis | 169 | 415979 | 20259 | 1112 | 3131598 | 203156 |
| 709.6 | Other specified diffuse diseases of connective tissue | 137 | 415979 | 20291 | 1291 | 3131598 | 202977 |
| 712 | Infective connective tissue disorders | 196 | 340057 | 96154 | 674 | 3288094 | 47098 |
| 713.5 | Arthropathy associated with neurological disorders | 170 | 340057 | 96180 | 381 | 3288094 | 47391 |
| 722.3 | Schmorl's nodes | 114 | 402672 | 33621 | 23 | 3203280 | 132563 |
| 722.8 | Postlaminectomy syndrome | 122 | 402672 | 33613 | 2479 | 3203280 | 130107 |
| 724.8 | Other symptoms referable to back | 201 | 402672 | 33534 | 103 | 3203280 | 132483 |
| 726.4 | Calcaneal spur; Exostosis NOS | 141 | 379146 | 57120 | 10066 | 3137629 | 188171 |
| 727.2 | Bursitis disorders | 189 | 379146 | 57072 | 721 | 3137629 | 197516 |
| 737 | Curvature of spine | 147 | 410135 | 26125 | 822 | 3263385 | 71659 |
| 737.2 | Lordosis (acquired) | 141 | 410135 | 26131 | 168 | 3263385 | 72313 |
| 741.1 | Ankylosis of joint | 180 | 423934 | 12293 | 384 | 3254285 | 81197 |
| 742 | Derangement of joint, non-traumatic | 122 | 423934 | 12351 | 24 | 3254285 | 81557 |
| 743.4 | Stress fracture | 126 | 416177 | 20104 | 2910 | 3265276 | 67680 |
| 748 | Anomalies of respiratory system, congenital | 119 | 435831 | 457 | 2040 | 3320556 | 13270 |
| 750.1 | Upper gastrointestinal congenital anomalies | 163 | 432301 | 3943 |  |  |  |
| 750.15 | Congenital anomalies of stomach | 107 | 432301 | 3999 | 392 | 3307101 | 28373 |
| 751.2 | Congenital anomalies of urinary system | 143 | 432301 | 3963 | 2162 | 3307101 | 26603 |
| 752.1 | Neural tube defects | 127 | 435312 | 968 | 9045 | 3318696 | 8125 |
| 753.1 | Congenital cataract and lens anomalies | 151 | 435312 | 944 | 554 | 3318696 | 16616 |
| 755.6 | Other congenital anomalies of lower limb, including pelvic girdle | 111 | 434047 | 2249 | 1340 | 3307069 | 27457 |
| 755.61 | Congenital hip dysplasia and deformity | 201 | 434047 | 2159 | 1842 | 3307069 | 26955 |

**Supplementary Table S2 (cont).**

| **Phecode** | **Phenotype** | **UK Biobank** | | | **Mount Sinai Data Warehouse** | | |
| --- | --- | --- | --- | --- | --- | --- | --- |
|  |  | **Cases** | **Controls** | **Excluded** | **Cases** | **Controls** | **Excluded** |
| 756 | Other congenital musculoskeletal anomalies | 115 | 434047 | 2245 | 1649 | 3307069 | 27148 |
| 756.3 | Congenital anomalies of muscle, tendon, fascia, and connective tissue | 170 | 434047 | 2190 | 1624 | 3307069 | 27173 |
| 758 | Chromosomal anomalies and genetic disorders | 144 | 434030 | 2233 | 1383 | 3312785 | 21698 |
| 758.1 | Chromosomal anomalies | 218 | 434030 | 2159 | 4176 | 3312785 | 18905 |
| 759.1 | Anomalies of endocrine glands, congenital | 152 | 434030 | 2225 | 851 | 3312785 | 22230 |
| 790.1 | Elevated sedimentation rate | 146 | 418908 | 17353 | 2951 | 3294655 | 38260 |
| 794 | Abnormal results of other function studies (bladder, pancreas, placenta, spleen, etc) | 176 | 436231 | 0 | 1142 | 3334724 | 0 |
| 797 | Shock | 150 | 435943 | 314 | 6805 | 3326627 | 2434 |
| 801.1 | Fracture of foot | 170 | 396337 | 39900 | 631 | 3239202 | 96033 |
| 860 | Bone marrow or stem cell transplant | 173 | 436234 | 0 | 135 | 3335731 | 0 |
| 870.8 | Open wound of genital organs | 100 | 418395 | 17912 | 888 | 3247916 | 87062 |
| 871.4 | Open wound of toe(s) | 177 | 418395 | 17835 | 2035 | 3247916 | 85915 |
| 913 | Toxic effect of venom | 203 | 436204 | 0 | 1891 | 3333960 | 15 |
| 938 | Dermatitis due to solar radiation | 198 | 420232 | 15977 | 2204 | 3076717 | 256945 |
| 960.1 | Adverse effects of antibacterials (not penicillins) | 125 | 371693 | 64589 | 2359 | 3297064 | 36443 |
| 975 | Poisoning by agents primarily acting on the smooth and skeletal muscles and respiratory system | 136 | 371693 | 64578 | 0 | 3297064 | 38802 |
| 987 | Toxic effect of other gases, fumes, or vapors | 165 | 434532 | 1710 | 1344 | 3331461 | 3061 |
| 989 | Toxic effect of other substances, chiefly nonmedicinal as to source | 109 | 434532 | 1766 | 1242 | 3331461 | 3163 |

**Supplementary Table S3. Model performance by phecode for 155 rare phecodes in UK Biobank cohort.** For each performance metric, table shows results from five cross-validated models (CV 1-5) and the median across all five models (Med).

|  | **Number of Positive Predictions** | | | | | | **Predictive Odds Ratio** | | | | | | **Positive Predictive Value** | | | | | |
| --- | --- | --- | --- | --- | --- | --- | --- | --- | --- | --- | --- | --- | --- | --- | --- | --- | --- | --- |
| **Phecode** | **CV 1** | **CV 2** | **CV 3** | **CV 4** | **CV 5** | **Med** | **CV 1** | **CV 2** | **CV 3** | **CV 4** | **CV 5** | **Med** | **CV 1** | **CV 2** | **CV 3** | **CV 4** | **CV 5** | **Med** |
| 008.7 | 3453 | 164 | 677 | 1414 | 389 | 677 | 15 | 39 | 11 | 14 | 21 | 15 | 5.8E-04 | 0 | 0 | 7.10E-04 | 0 | 0 |
| 070.1 | 1364 | 1263 | 212 | 2369 | 1407 | 1364 | 15 | 9.9 | 89 | 13 | 17 | 15 | 0.0015 | 7.9E-04 | 0.0094 | 0.0013 | 0.0014 | 0.0014 |
| 071 | 2217 | 486 | 2030 | 3476 | 2880 | 2217 | 26 | 100 | 46 | 34 | 9.5 | 34 | 0.0018 | 0.0062 | 0.0034 | 0.0020 | 3.5E-04 | 0.0020 |
| 071.1 | 1222 | 777 | 687 | 2441 | 604 | 777 | 42 | 77 | 85 | 41 | 89 | 77 | 0.0098 | 0.017 | 0.020 | 0.0078 | 0.017 | 0.017 |
| 079.1 | 2607 | 954 | 479 | 1613 | 531 | 954 | 32 | 72 | 33 | 31 | 56 | 33 | 0.0019 | 0.0063 | 0.0021 | 0.0025 | 0.0038 | 0.0025 |
| 079.2 | 3468 | 1210 | 1503 | 550 | 269 | 1210 | 12 | 48 | 62 | 45 | 27 | 45 | 5.8E-04 | 0.0017 | 0.0027 | 0.0018 | 0 | 0.0017 |
| 110.11 | 877 | 43 | 592 | 661 | 37 | 592 | 20 | 44 | 33 | 11 | 170 | 33 | 0.0046 | 0 | 0.0068 | 0.0015 | 0.027 | 0.0046 |
| 110.13 | 568 | 213 | 1200 | 2424 | 765 | 765 | 140 | 29 | 62 | 56 | 27 | 56 | 0.0088 | 0 | 0.0042 | 0.0033 | 0.0013 | 0.0033 |
| 110.2 | 1720 | 790 | 185 | 2206 | 3067 | 1720 | 30 | 40 | 66 | 53 | 23 | 40 | 0.0035 | 0.0038 | 0.0054 | 0.0050 | 0.0023 | 0.0038 |
| 112.3 | 4799 | 1836 | 843 | 6760 | 875 | 1836 | 40 | 110 | 28 | 35 | 88 | 40 | 0.0029 | 0.0060 | 0.0024 | 0.0016 | 0.0057 | 0.0029 |
| 130 | 270 | 93 | 122 | 208 | 7 | 122 | 29 | 26 | 21 | 15 | 380 | 26 | 0.0037 | 0 | 0 | 0 | 0 | 0 |
| 145.1 | 3068 | 1561 | 1024 | 2880 | 1355 | 1561 | 29 | 31 | 6.3 | 22 | 51 | 29 | 9.8E-04 | 0.0013 | 0 | 0.0010 | 0.0022 | 0.0010 |
| 149 | 3628 | 3238 | 1331 | 3644 | 2563 | 3238 | 84 | 64 | 160 | 180 | 63 | 84 | 0.0041 | 0.0031 | 0.011 | 0.0074 | 0.0043 | 0.0043 |
| 149.3 | 1953 | 1186 | 1163 | 2972 | 2404 | 1953 | 170 | 160 | 220 | 140 | 210 | 170 | 0.0087 | 0.0084 | 0.011 | 0.0057 | 0.0075 | 0.0084 |
| 149.9 | 4971 | 1141 | 1155 | 1201 | 907 | 1155 | 55 | 63 | 140 | 200 | 150 | 140 | 0.0024 | 0.0044 | 0.0069 | 0.0075 | 0.0088 | 0.0069 |
| 164 | 3366 | 288 | 461 | 2169 | 1455 | 1455 | 110 | 92 | 140 | 85 | 93 | 93 | 0.0051 | 0.0069 | 0.013 | 0.0046 | 0.0055 | 0.0055 |
| 174.3 | 1416 | 135 | 695 | 724 | 324 | 695 | 20 | 58 | 19 | 43 | 7.2 | 20 | 0.0028 | 0.0074 | 0.0029 | 0.0069 | 0 | 0.0029 |
| 187.1 | 1421 | 702 | 773 | 1156 | 717 | 773 | 53 | 88 | 60 | 54 | 56 | 56 | 0.014 | 0.020 | 0.019 | 0.014 | 0.014 | 0.014 |
| 187.8 | 8217 | 266 | 1695 | 1925 | 1703 | 1703 | 4.2 | 12 | 7.0 | 8.0 | 7.6 | 7.6 | 0.0016 | 0.0038 | 0.0029 | 0.0031 | 0.0029 | 0.0029 |
| 189.12 | 1301 | 434 | 1126 | 1481 | 202 | 1126 | 70 | 92 | 50 | 77 | 15 | 70 | 0.0085 | 0.012 | 0.0080 | 0.0095 | 0 | 0.0085 |
| 191.1 | 5975 | 1441 | 1624 | 2910 | 826 | 1624 | 1.6 | 31 | 42 | 38 | 51 | 38 | 0 | 0.0014 | 0.0018 | 0.0024 | 0.0024 | 0.0018 |
| 194 | 1320 | 244 | 89 | 340 | 79 | 244 | 43 | 110 | 240 | 130 | 28 | 110 | 0.0068 | 0.016 | 0.034 | 0.018 | 0 | 0.016 |
| 199.4 | 366 | 174 | 139 | 211 | 152 | 174 | 260 | 430 | 430 | 310 | 210 | 310 | 0.044 | 0.069 | 0.072 | 0.052 | 0.039 | 0.052 |
| 204.11 | 3206 | 4312 | 2812 | 4320 | 897 | 3206 | 200 | 130 | 290 | 90 | 240 | 200 | 0.0078 | 0.0044 | 0.0096 | 0.0035 | 0.011 | 0.0078 |
| 204.3 | 1360 | 2245 | 237 | 2080 | 731 | 1360 | 150 | 190 | 230 | 42 | 220 | 190 | 0.0088 | 0.0062 | 0.017 | 0.0019 | 0.011 | 0.0088 |

**Supplementary Table S3 (cont.)**

|  | **Number of Positive Predictions** | | | | | | **Predictive Odds Ratio** | | | | | | **Positive Predictive Value** | | | | | |
| --- | --- | --- | --- | --- | --- | --- | --- | --- | --- | --- | --- | --- | --- | --- | --- | --- | --- | --- |
| **Phecode** | **CV 1** | **CV 2** | **CV 3** | **CV 4** | **CV 5** | **Med** | **CV 1** | **CV 2** | **CV 3** | **CV 4** | **CV 5** | **Med** | **CV 1** | **CV 2** | **CV 3** | **CV 4** | **CV 5** | **Med** |
| 224 | 513 | 158 | 130 | 819 | 151 | 158 | 10 | 110 | 33 | 7.4 | 33 | 33 | 0 | 0.0063 | 0 | 0 | 0 | 0 |
| 224.1 | 1672 | 587 | 737 | 1345 | 1830 | 1345 | 20 | 15 | 31 | 15 | 27 | 20 | 0.0054 | 0.0034 | 0.0081 | 0.0037 | 0.0066 | 0.0054 |
| 225.2 | 1532 | 1392 | 1467 | 2772 | 1627 | 1532 | 87 | 37 | 26 | 44 | 19 | 37 | 0.0059 | 0.0022 | 0.0014 | 0.0018 | 6.1E-04 | 0.0018 |
| 242.2 | 522 | 15 | 82 | 920 | 2 | 82 | 8.9 | 100 | 19 | 1.9 | 680 | 19 | 0.0019 | 0 | 0 | 0 | 0 | 0 |
| 242.3 | 1318 | 955 | 2011 | 1313 | 285 | 1313 | 34 | 52 | 38 | 47 | 88 | 47 | 0.0076 | 0.013 | 0.0075 | 0.0099 | 0.021 | 0.0099 |
| 246.7 | 4845 | 3582 | 1110 | 4895 | 322 | 3582 | 53 | 44 | 32 | 47 | 67 | 47 | 0.0043 | 0.0050 | 0.0045 | 0.0043 | 0.0093 | 0.0045 |
| 250.12 | 310 | 107 | 158 | 243 | 209 | 209 | 17000 | 33000 | 36000 | 35000 | 15000 | 33000 | 0.12 | 0.32 | 0.28 | 0.14 | 0.14 | 0.14 |
| 250.3 | 9046 | 781 | 693 | 3454 | 933 | 933 | 15 | 34 | 89 | 63 | 130 | 63 | 0.0012 | 0.0026 | 0.0087 | 0.0032 | 0.013 | 0.0032 |
| 250.5 | 3209 | 1435 | 2187 | 3015 | 1577 | 2187 | 100 | 100 | 73 | 120 | 86 | 100 | 0.012 | 0.017 | 0.011 | 0.014 | 0.015 | 0.014 |
| 253 | 1260 | 756 | 498 | 1402 | 168 | 756 | 140 | 190 | 200 | 100 | 370 | 190 | 0.021 | 0.032 | 0.040 | 0.017 | 0.071 | 0.032 |
| 253.11 | 2020 | 684 | 346 | 623 | 299 | 623 | 120 | 590 | 1100 | 460 | 800 | 590 | 0.010 | 0.038 | 0.072 | 0.034 | 0.067 | 0.038 |
| 253.3 | 5143 | 2028 | 1620 | 3427 | 1716 | 2028 | 33 | 45 | 73 | 46 | 61 | 46 | 0.0035 | 0.0049 | 0.0074 | 0.0047 | 0.0064 | 0.0049 |
| 255.11 | 844 | 777 | 358 | 4858 | 786 | 786 | 130 | 94 | 170 | 31 | 120 | 120 | 0.020 | 0.017 | 0.028 | 0.0043 | 0.018 | 0.018 |
| 255.12 | 4434 | 781 | 251 | 4159 | 1599 | 1599 | 43 | 150 | 200 | 37 | 81 | 81 | 0.0045 | 0.018 | 0.028 | 0.0038 | 0.0088 | 0.0088 |
| 258 | 2846 | 373 | 189 | 1061 | 681 | 681 | 98 | 390 | 1200 | 270 | 270 | 270 | 0.011 | 0.048 | 0.12 | 0.026 | 0.028 | 0.028 |
| 261.41 | 2628 | 118 | 329 | 1294 | 141 | 329 | 24 | 100 | 11 | 29 | 26 | 26 | 0.0019 | 0.0085 | 0 | 0.0031 | 0 | 0.0019 |
| 270.35 | 1182 | 730 | 631 | 681 | 816 | 730 | 4500 | 1700 | 2800 | 3000 | 4400 | 3000 | 0.055 | 0.071 | 0.098 | 0.069 | 0.065 | 0.069 |
| 272.12 | 2164 | 100 | 295 | 1383 | 233 | 295 | 6.8 | 56 | 7.1 | 4.2 | 8.7 | 7.1 | 0.0014 | 0.010 | 0 | 7.2E-04 | 0 | 7.2E-04 |
| 277.51 | 1055 | 245 | 473 | 532 | 139 | 473 | 35 | 110 | 36 | 57 | 99 | 57 | 0.0028 | 0.0082 | 0.0021 | 0.0038 | 0.0072 | 0.0038 |
| 278.3 | 608 | 170 | 53 | 1917 | 904 | 608 | 28 | 28 | 93 | 12 | 7.6 | 28 | 0.0016 | 0 | 0 | 5.2E-04 | 0 | 0 |
| 278.4 | 2607 | 113 | 547 | 2840 | 291 | 547 | 6.3 | 27 | 30 | 11 | 11 | 11 | 7.7E-04 | 0 | 0.0037 | 0.0014 | 0 | 7.7E-04 |
| 281.9 | 7775 | 2598 | 955 | 4880 | 131 | 2598 | 31 | 60 | 56 | 39 | 200 | 56 | 0.0023 | 0.0046 | 0.0063 | 0.0037 | 0.023 | 0.0046 |
| 286.11 | 1608 | 300 | 81 | 2195 | 76 | 300 | 14 | 26 | 86 | 5.9 | 32 | 26 | 0.0025 | 0.0033 | 0.012 | 9.1E-04 | 0 | 0.0025 |
| 286.13 | 1001 | 223 | 398 | 2800 | 185 | 398 | 37 | 28 | 180 | 27 | 340 | 37 | 0.0030 | 0 | 0.010 | 0.0014 | 0.016 | 0.0030 |
| 286.5 | 1798 | 360 | 897 | 3958 | 424 | 897 | 190 | 480 | 510 | 110 | 510 | 480 | 0.010 | 0.028 | 0.028 | 0.0033 | 0.028 | 0.028 |
| 288.3 | 2596 | 681 | 945 | 703 | 374 | 703 | 19 | 45 | 35 | 39 | 40 | 39 | 0.0042 | 0.0088 | 0.0074 | 0.0085 | 0.008 | 0.0080 |
| 289.9 | 5895 | 308 | 1204 | 8582 | 1195 | 1204 | 14 | 43 | 29 | 12 | 39 | 29 | 0.0025 | 0.0065 | 0.0050 | 0.0017 | 0.0067 | 0.0050 |

**Supplementary Table S3 (cont.)**

|  | **Number of Positive Predictions** | | | | | | **Predictive Odds Ratio** | | | | | | **Positive Predictive Value** | | | | | |
| --- | --- | --- | --- | --- | --- | --- | --- | --- | --- | --- | --- | --- | --- | --- | --- | --- | --- | --- |
| **Phecode** | **CV 1** | **CV 2** | **CV 3** | **CV 4** | **CV 5** | **Med** | **CV 1** | **CV 2** | **CV 3** | **CV 4** | **CV 5** | **Med** | **CV 1** | **CV 2** | **CV 3** | **CV 4** | **CV 5** | **Med** |
| 290.3 | 5817 | 1305 | 2817 | 3006 | 1635 | 2817 | 41 | 130 | 86 | 93 | 79 | 86 | 0.0045 | 0.019 | 0.0092 | 0.0086 | 0.0098 | 0.0092 |
| 297 | 4060 | 1497 | 742 | 1429 | 1634 | 1497 | 20 | 12 | 24 | 30 | 25 | 24 | 0.0022 | 0.0013 | 0.0027 | 0.0042 | 0.0024 | 0.0024 |
| 300.4 | 3473 | 348 | 539 | 2670 | 1100 | 1100 | 11 | 260 | 10 | 30 | 51 | 30 | 8.6E-04 | 0.014 | 0 | 0.0015 | 0.0027 | 0.0015 |
| 305.2 | 2524 | 126 | 967 | 928 | 568 | 928 | 32 | 210 | 40 | 42 | 56 | 42 | 0.0059 | 0.048 | 0.0072 | 0.0086 | 0.012 | 0.0086 |
| 315.3 | 889 | 2 | 454 | 799 | 499 | 499 | 45 | 860 | 43 | 68 | 60 | 60 | 0.0079 | 0 | 0.0066 | 0.010 | 0.010 | 0.0079 |
| 323.2 | 217 | 210 | 207 | 236 | 240 | 217 | 8100 | 5700 | 3700 | 22000 | 20000 | 8100 | 0.15 | 0.13 | 0.13 | 0.18 | 0.12 | 0.13 |
| 325 | 1073 | 221 | 305 | 1164 | 324 | 324 | 8.2 | 33 | 70 | 76 | 150 | 70 | 0 | 0 | 0.0033 | 0.0034 | 0.0062 | 0.0033 |
| 337.1 | 5546 | 1473 | 3550 | 2100 | 1990 | 2100 | 34 | 48 | 38 | 47 | 41 | 41 | 0.0032 | 0.0061 | 0.0039 | 0.0052 | 0.0055 | 0.0052 |
| 353.1 | 456 | 99 | 255 | 1687 | 351 | 351 | 13 | 59 | 22 | 9.7 | 16 | 16 | 0.0022 | 0.010 | 0.0039 | 0.0024 | 0.0028 | 0.0028 |
| 353.2 | 2030 | 1015 | 854 | 1630 | 921 | 1015 | 49 | 60 | 25 | 75 | 50 | 50 | 0.0034 | 0.0049 | 0.0023 | 0.0067 | 0.0043 | 0.0043 |
| 361.2 | 8194 | 4567 | 4803 | 2898 | 3224 | 4567 | 31 | 42 | 48 | 40 | 39 | 40 | 0.0050 | 0.0066 | 0.0075 | 0.0079 | 0.0081 | 0.0075 |
| 362.3 | 2307 | 659 | 5405 | 2350 | 1780 | 2307 | 76 | 37 | 30 | 41 | 52 | 41 | 0.0065 | 0.0046 | 0.0024 | 0.0043 | 0.0067 | 0.0046 |
| 364.41 | 3900 | 4510 | 5164 | 3558 | 3682 | 3900 | 19 | 27 | 30 | 13 | 24 | 24 | 0.0028 | 0.0042 | 0.0039 | 0.0020 | 0.0033 | 0.0033 |
| 366.1 | 553 | 43 | 78 | 1104 | 134 | 134 | 27 | 88 | 54 | 18 | 33 | 33 | 0.0018 | 0 | 0 | 0.0018 | 0 | 0 |
| 366.3 | 3039 | 361 | 670 | 3799 | 1874 | 1874 | 23 | 150 | 53 | 33 | 50 | 50 | 0.0036 | 0.025 | 0.0090 | 0.0045 | 0.0059 | 0.0059 |
| 367.4 | 3698 | 2520 | 7438 | 2859 | 1625 | 2859 | 15 | 21 | 7.5 | 15 | 19 | 15 | 0.0016 | 0.0020 | 6.7E-04 | 0.0014 | 0.0018 | 0.0016 |
| 377.1 | 1730 | 109 | 2873 | 1313 | 205 | 1313 | 58 | 160 | 60 | 68 | 110 | 68 | 0.0069 | 0.018 | 0.0063 | 0.0091 | 0.015 | 0.0091 |
| 378 | 1041 | 365 | 691 | 1188 | 164 | 691 | 30 | 9.0 | 45 | 28 | 58 | 30 | 0.0038 | 0 | 0.0058 | 0.0034 | 0.0061 | 0.0038 |
| 388 | 596 | 137 | 422 | 403 | 283 | 403 | 110 | 230 | 150 | 120 | 150 | 150 | 0.017 | 0.036 | 0.026 | 0.020 | 0.025 | 0.025 |
| 394.4 | 3019 | 344 | 1816 | 2106 | 528 | 1816 | 78 | 460 | 160 | 130 | 320 | 160 | 0.0053 | 0.017 | 0.0039 | 0.0047 | 0.011 | 0.0053 |
| 401.2 | 1434 | 699 | 880 | 1549 | 763 | 880 | 650 | 1100 | 560 | 430 | 530 | 560 | 0.017 | 0.043 | 0.026 | 0.017 | 0.029 | 0.026 |
| 420.22 | 5452 | 1081 | 2076 | 2096 | 604 | 2076 | 76 | 76 | 77 | 62 | 130 | 76 | 0.0079 | 0.015 | 0.011 | 0.010 | 0.025 | 0.011 |
| 425.2 | 518 | 68 | 441 | 447 | 187 | 441 | 1700 | 6400 | 2000 | 3000 | 4500 | 3000 | 0.044 | 0.26 | 0.057 | 0.060 | 0.15 | 0.06 |
| 425.8 | 1632 | 1476 | 1736 | 1116 | 619 | 1476 | 190 | 160 | 100 | 120 | 380 | 160 | 0.0061 | 0.0068 | 0.0046 | 0.0072 | 0.016 | 0.0068 |
| 426.8 | 9438 | 1480 | 3177 | 3768 | 505 | 3177 | 30 | 64 | 45 | 52 | 100 | 52 | 0.0028 | 0.0095 | 0.0054 | 0.0053 | 0.016 | 0.0054 |
| 441.2 | 8404 | 1339 | 3133 | 3064 | 325 | 3064 | 22 | 37 | 26 | 11 | 58 | 26 | 0.0020 | 0.0037 | 0.0032 | 9.8E-04 | 0.0062 | 0.0032 |

**Supplementary Table S3 (cont.)**

|  | **Number of Positive Predictions** | | | | | | **Predictive Odds Ratio** | | | | | | **Positive Predictive Value** | | | | | |
| --- | --- | --- | --- | --- | --- | --- | --- | --- | --- | --- | --- | --- | --- | --- | --- | --- | --- | --- |
| **Phecode** | **CV 1** | **CV 2** | **CV 3** | **CV 4** | **CV 5** | **Med** | **CV 1** | **CV 2** | **CV 3** | **CV 4** | **CV 5** | **Med** | **CV 1** | **CV 2** | **CV 3** | **CV 4** | **CV 5** | **Med** |
| 446 | 4713 | 588 | 1464 | 3409 | 859 | 1464 | 31 | 180 | 80 | 41 | 93 | 80 | 0.0038 | 0.022 | 0.0096 | 0.0053 | 0.012 | 0.0096 |
| 459.1 | 708 | 105 | 9 | 1143 | 18 | 105 | 68 | 55 | 190 | 34 | 410 | 68 | 0.014 | 0.0095 | 0 | 0.0070 | 0.056 | 0.0095 |
| 473.1 | 2346 | 550 | 679 | 500 | 329 | 550 | 16 | 21 | 18 | 39 | 38 | 21 | 0.0017 | 0.0018 | 0.0015 | 0.004 | 0.0030 | 0.0018 |
| 500.1 | 1573 | 3419 | 1045 | 3596 | 871 | 1573 | 49 | 38 | 21 | 35 | 28 | 35 | 0.010 | 0.0064 | 0.0048 | 0.0064 | 0.0057 | 0.0064 |
| 519 | 4209 | 905 | 982 | 2528 | 752 | 982 | 200 | 170 | 100 | 210 | 220 | 200 | 0.012 | 0.024 | 0.020 | 0.020 | 0.033 | 0.020 |
| 523.1 | 290 | 102 | 648 | 623 | 342 | 342 | 20 | 18 | 9.3 | 2.9 | 5.2 | 9.3 | 0.0034 | 0 | 0.0015 | 0 | 0 | 0 |
| 527.1 | 131 | 2 | 237 | 474 | 92 | 131 | 53 | 2400 | 35 | 41 | 72 | 53 | 0 | 0 | 0 | 0.0021 | 0 | 0 |
| 527.7 | 1794 | 308 | 341 | 938 | 471 | 471 | 100 | 97 | 75 | 130 | 140 | 100 | 0.012 | 0.016 | 0.015 | 0.018 | 0.021 | 0.016 |
| 528.12 | 1607 | 45 | 203 | 1249 | 220 | 220 | 3.1 | 77 | 66 | 19 | 17 | 19 | 0 | 0 | 0.0049 | 0.0016 | 0 | 0 |
| 528.41 | 1421 | 73 | 201 | 289 | 166 | 201 | 6.2 | 40 | 16 | 120 | 19 | 19 | 7.0E-04 | 0 | 0 | 0.017 | 0 | 0 |
| 573.2 | 665 | 223 | 231 | 881 | 229 | 231 | 680 | 790 | 1200 | 730 | 1100 | 790 | 0.038 | 0.072 | 0.091 | 0.041 | 0.087 | 0.072 |
| 580.11 | 2941 | 740 | 1267 | 1494 | 189 | 1267 | 290 | 530 | 350 | 240 | 460 | 350 | 0.0075 | 0.032 | 0.014 | 0.011 | 0.037 | 0.014 |
| 588 | 6163 | 1518 | 903 | 236 | 143 | 903 | 68 | 150 | 220 | 540 | 850 | 220 | 0.0052 | 0.016 | 0.024 | 0.064 | 0.10 | 0.024 |
| 597.2 | 291 | 224 | 847 | 1155 | 662 | 662 | 1600 | 1300 | 730 | 680 | 1100 | 1100 | 0.11 | 0.13 | 0.045 | 0.042 | 0.056 | 0.056 |
| 609.2 | 635 | 1019 | 388 | 1919 | 330 | 635 | 11 | 18 | 19 | 6.0 | 14 | 14 | 0.0079 | 0.012 | 0.013 | 0.0042 | 0.0091 | 0.0091 |
| 611.1 | 1163 | 1765 | 1975 | 3238 | 647 | 1765 | 25 | 31 | 17 | 9.5 | 13 | 17 | 0.0017 | 0.0011 | 5.1E-04 | 3.1E-04 | 0 | 5.1E-04 |
| 612.3 | 1639 | 417 | 1915 | 5426 | 232 | 1639 | 3.1 | 13 | 3.1 | 3.3 | 21 | 3.3 | 0 | 0 | 0 | 1.8E-04 | 0 | 0 |
| 613.5 | 1804 | 1888 | 1811 | 8409 | 1337 | 1811 | 21 | 9.4 | 13 | 12 | 16 | 13 | 0.0028 | 0.0011 | 0.0017 | 0.0014 | 0.0022 | 0.0017 |
| 620 | 2198 | 1985 | 952 | 3071 | 1480 | 1985 | 50 | 51 | 58 | 37 | 79 | 51 | 0.0064 | 0.0065 | 0.0095 | 0.0046 | 0.010 | 0.0065 |
| 626.11 | 2981 | 731 | 1538 | 2133 | 2213 | 2133 | 4.7 | 12 | 8.7 | 8.4 | 8.7 | 8.7 | 0.0017 | 0.0041 | 0.0026 | 0.0033 | 0.0027 | 0.0027 |
| 626.4 | 3403 | 847 | 2849 | 1510 | 493 | 1510 | 8.3 | 8.7 | 12 | 11 | 21 | 11 | 0.0032 | 0.0035 | 0.0049 | 0.0046 | 0.0081 | 0.0046 |
| 643.1 | 9457 | 7394 | 10241 | 8270 | 7893 | 8270 | 51 | 53 | 51 | 32 | 41 | 51 | 0.0051 | 0.0066 | 0.0043 | 0.0053 | 0.0056 | 0.0053 |
| 647 | 6533 | 5930 | 5326 | 5909 | 6496 | 5930 | 110 | 66 | 77 | 46 | 62 | 66 | 0.0047 | 0.0047 | 0.0051 | 0.0037 | 0.0045 | 0.0047 |
| 649 | 6528 | 6160 | 6984 | 7576 | 5802 | 6528 | 29 | 46 | 75 | 41 | 46 | 46 | 0.0046 | 0.0055 | 0.0070 | 0.0049 | 0.0062 | 0.0055 |
| 656.2 | 2327 | 502 | 300 | 1011 | 210 | 502 | 220 | 170 | 260 | 230 | 200 | 220 | 0.019 | 0.026 | 0.040 | 0.027 | 0.029 | 0.027 |
| 686.5 | 162 | 54 | 87 | 693 | 9 | 87 | 200 | 120 | 280 | 73 | 820 | 200 | 0.012 | 0 | 0.011 | 0.0029 | 0 | 0.0029 |

**Supplementary Table S3 (cont.)**

|  | **Number of Positive Predictions** | | | | | | **Predictive Odds Ratio** | | | | | | **Positive Predictive Value** | | | | | |
| --- | --- | --- | --- | --- | --- | --- | --- | --- | --- | --- | --- | --- | --- | --- | --- | --- | --- | --- |
| **Phecode** | **CV 1** | **CV 2** | **CV 3** | **CV 4** | **CV 5** | **Med** | **CV 1** | **CV 2** | **CV 3** | **CV 4** | **CV 5** | **Med** | **CV 1** | **CV 2** | **CV 3** | **CV 4** | **CV 5** | **Med** |
| 690.1 | 6275 | 288 | 13 | 602 | 299 | 299 | 2.4 | 22 | 150 | 17 | 37 | 22 | 4.8E-04 | 0.0035 | 0 | 0.0033 | 0.0067 | 0.0033 |
| 691 | 1043 | 91 | 94 | 271 | 93 | 94 | 2000 | 20000 | 26000 | 38000 | 34000 | 26000 | 0.026 | 0.41 | 0.43 | 0.14 | 0.35 | 0.35 |
| 694.1 | 912 | 365 | 1326 | 3918 | 351 | 912 | 22 | 30 | 10 | 7.7 | 18 | 18 | 0.0044 | 0.0055 | 0.0023 | 0.0018 | 0.0028 | 0.0028 |
| 695.21 | 1624 | 1213 | 1067 | 1477 | 1362 | 1362 | 42 | 35 | 60 | 96 | 58 | 58 | 0.0025 | 0.0025 | 0.0028 | 0.0041 | 0.0037 | 0.0028 |
| 695.81 | 2920 | 115 | 89 | 1991 | 1383 | 1383 | 4.3 | 36 | 38 | 13 | 3.2 | 13 | 3.4E-04 | 0 | 0 | 0.0010 | 0 | 0 |
| 696.3 | 1079 | 390 | 578 | 868 | 398 | 578 | 180 | 160 | 200 | 180 | 300 | 180 | 0.0074 | 0.0077 | 0.0087 | 0.0081 | 0.018 | 0.0081 |
| 701.1 | 2270 | 1144 | 178 | 1670 | 319 | 1144 | 2.9 | 4.6 | 35 | 3.7 | 17 | 4.6 | 0 | 0 | 0 | 0 | 0 | 0 |
| 701.3 | 494 | 493 | 182 | 406 | 167 | 406 | 84 | 210 | 460 | 420 | 930 | 420 | 0.0040 | 0.012 | 0.022 | 0.022 | 0.048 | 0.022 |
| 701.6 | 782 | 184 | 322 | 1327 | 356 | 356 | 37 | 69 | 63 | 20 | 25 | 37 | 0.0090 | 0.016 | 0.016 | 0.0053 | 0.0056 | 0.0090 |
| 704.8 | 829 | 672 | 16 | 587 | 159 | 587 | 10 | 4.5 | 190 | 5.4 | 54 | 10 | 0.0012 | 0 | 0 | 0 | 0.0063 | 0 |
| 706.1 | 544 | 148 | 130 | 2931 | 31 | 148 | 44 | 20 | 21 | 8.2 | 100 | 21 | 0.0055 | 0 | 0 | 0.0010 | 0 | 0 |
| 709.5 | 295 | 635 | 643 | 1054 | 198 | 635 | 150 | 52 | 160 | 110 | 260 | 150 | 0.024 | 0.0079 | 0.023 | 0.013 | 0.035 | 0.023 |
| 709.6 | 894 | 188 | 553 | 447 | 628 | 553 | 160 | 250 | 71 | 160 | 150 | 160 | 0.012 | 0.027 | 0.0072 | 0.013 | 0.011 | 0.012 |
| 712 | 453 | 277 | 427 | 570 | 1073 | 453 | 35000 | 4500 | 9100 | 24000 | 11000 | 11000 | 0.16 | 0.22 | 0.15 | 0.11 | 0.058 | 0.15 |
| 713.5 | 1121 | 719 | 459 | 345 | 264 | 459 | 810 | 770 | 1600 | 1900 | 1600 | 1600 | 0.029 | 0.043 | 0.081 | 0.096 | 0.11 | 0.081 |
| 722.3 | 3291 | 837 | 2084 | 4308 | 2804 | 2804 | 79 | 140 | 76 | 70 | 54 | 76 | 0.0036 | 0.0084 | 0.0038 | 0.003 | 0.0036 | 0.0036 |
| 722.8 | 957 | 469 | 1396 | 1196 | 1031 | 1031 | 480 | 450 | 230 | 230 | 430 | 430 | 0.018 | 0.028 | 0.0093 | 0.013 | 0.015 | 0.015 |
| 724.8 | 2017 | 516 | 167 | 1252 | 220 | 516 | 62 | 190 | 220 | 72 | 120 | 120 | 0.0099 | 0.033 | 0.048 | 0.014 | 0.027 | 0.027 |
| 726.4 | 929 | 2664 | 933 | 3045 | 2884 | 2664 | 39 | 15 | 12 | 15 | 12 | 15 | 0.0054 | 0.0015 | 0.0011 | 0.0026 | 0.0014 | 0.0015 |
| 727.2 | 272 | 503 | 828 | 334 | 517 | 503 | 55 | 36 | 27 | 18 | 37 | 36 | 0.011 | 0.0080 | 0.0060 | 0.0030 | 0.0077 | 0.0077 |
| 737 | 6240 | 2298 | 740 | 4564 | 1264 | 2298 | 32 | 20 | 54 | 23 | 24 | 24 | 0.0024 | 0.0017 | 0.0068 | 0.0022 | 0.0032 | 0.0024 |
| 737.2 | 3537 | 5248 | 2466 | 3558 | 2949 | 3537 | 38 | 36 | 24 | 12 | 25 | 25 | 0.0034 | 0.0029 | 0.0028 | 0.0011 | 0.0024 | 0.0028 |
| 741.1 | 1648 | 293 | 2113 | 1263 | 309 | 1263 | 14 | 50 | 18 | 12 | 29 | 18 | 0.0024 | 0.0068 | 0.0033 | 0.0016 | 0.0032 | 0.0032 |
| 742 | 2311 | 1112 | 2549 | 2736 | 978 | 2311 | 40 | 45 | 44 | 28 | 24 | 40 | 0.0030 | 0.0036 | 0.0031 | 0.0026 | 0.0020 | 0.0030 |
| 743.4 | 3352 | 2043 | 1726 | 1159 | 1054 | 1726 | 22 | 49 | 30 | 22 | 120 | 30 | 0.0024 | 0.0039 | 0.0023 | 0.0017 | 0.0095 | 0.0024 |
| 748 | 843 | 245 | 319 | 1111 | 236 | 319 | 7.2 | 120 | 25 | 51 | 100 | 51 | 0 | 0.0082 | 0 | 0.0036 | 0.0085 | 0.0036 |

**Supplementary Table S3 (cont.)**

|  | **Number of Positive Predictions** | | | | | | **Predictive Odds Ratio** | | | | | | **Positive Predictive Value** | | | | | |
| --- | --- | --- | --- | --- | --- | --- | --- | --- | --- | --- | --- | --- | --- | --- | --- | --- | --- | --- |
| **Phecode** | **CV 1** | **CV 2** | **CV 3** | **CV 4** | **CV 5** | **Med** | **CV 1** | **CV 2** | **CV 3** | **CV 4** | **CV 5** | **Med** | **CV 1** | **CV 2** | **CV 3** | **CV 4** | **CV 5** | **Med** |
| 750.1 | 5806 | 217 | 1258 | 3182 | 156 | 1258 | 4.9 | 19 | 7.5 | 16 | 24 | 16 | 5.2E-04 | 0 | 7.9E-04 | 0.0016 | 0 | 5.2E-04 |
| 750.15 | 4332 | 443 | 780 | 10913 | 735 | 780 | 8.0 | 19 | 7.5 | 41 | 34 | 19 | 4.6E-04 | 0 | 0 | 0.0010 | 0.0014 | 4.6E-04 |
| 751.2 | 4652 | 2411 | 2217 | 1691 | 1738 | 2217 | 12 | 13 | 13 | 10 | 18 | 13 | 0.0015 | 0.0017 | 0.0018 | 0.0012 | 0.0023 | 0.0017 |
| 752.1 | 1869 | 150 | 292 | 1461 | 23 | 292 | 18 | 84 | 17 | 3.2 | 190 | 18 | 0.0016 | 0.0067 | 0 | 0 | 0 | 0 |
| 753.1 | 2826 | 308 | 2292 | 1605 | 848 | 1605 | 15 | 46 | 28 | 19 | 11 | 19 | 0.0021 | 0.0065 | 0.0035 | 0.0025 | 0.0012 | 0.0025 |
| 755.6 | 991 | 96 | 463 | 1256 | 163 | 463 | 24 | 61 | 17 | 4.7 | 95 | 24 | 0.0010 | 0 | 0 | 0 | 0.0061 | 0 |
| 755.61 | 1813 | 1177 | 161 | 573 | 180 | 573 | 33 | 47 | 280 | 67 | 150 | 67 | 0.0066 | 0.010 | 0.056 | 0.016 | 0.033 | 0.016 |
| 756 | 1337 | 145 | 73 | 777 | 96 | 145 | 4.4 | 42 | 110 | 6.6 | 63 | 42 | 0 | 0 | 0 | 0 | 0 | 0 |
| 756.3 | 2217 | 218 | 205 | 1357 | 230 | 230 | 33 | 46 | 50 | 30 | 110 | 46 | 0.0041 | 0.0046 | 0.0049 | 0.0044 | 0.013 | 0.0046 |
| 758 | 1951 | 253 | 1086 | 2810 | 863 | 1086 | 45 | 190 | 61 | 28 | 99 | 61 | 0.0046 | 0.020 | 0.0064 | 0.0032 | 0.0093 | 0.0064 |
| 758.1 | 1084 | 37 | 94 | 281 | 16 | 94 | 24 | 53 | 21 | 7.3 | 110 | 24 | 0.0046 | 0 | 0 | 0 | 0 | 0 |
| 759.1 | 504 | 92 | 208 | 230 | 361 | 230 | 5.8 | 35 | 13 | 37 | 7.5 | 13 | 0 | 0 | 0 | 0.0043 | 0 | 0 |
| 790.1 | 603 | 1324 | 418 | 2089 | 1301 | 1301 | 19 | 14 | 39 | 27 | 39 | 27 | 0.0017 | 0.0015 | 0.0048 | 0.0029 | 0.0038 | 0.0029 |
| 794 | 4381 | 3794 | 5459 | 3141 | 1870 | 3794 | 74 | 91 | 69 | 80 | 86 | 80 | 0.0073 | 0.0090 | 0.0066 | 0.011 | 0.013 | 0.0090 |
| 797 | 6457 | 1260 | 1816 | 3952 | 2400 | 2400 | 24 | 55 | 46 | 46 | 43 | 46 | 0.0022 | 0.0063 | 0.0050 | 0.0043 | 0.0046 | 0.0046 |
| 801.1 | 3205 | 870 | 1295 | 795 | 2386 | 1295 | 5.3 | 7.7 | 5.7 | 15 | 2.7 | 5.7 | 9.4E-04 | 0.0011 | 7.7E-04 | 0.0025 | 4.2E-04 | 9.4E-04 |
| 860 | 1641 | 1464 | 657 | 1110 | 1344 | 1344 | 1700 | 1900 | 1200 | 2500 | 1500 | 1700 | 0.026 | 0.029 | 0.046 | 0.032 | 0.028 | 0.029 |
| 870.8 | 1248 | 283 | 172 | 845 | 116 | 283 | 5.7 | 34 | 45 | 8.8 | 82 | 34 | 0 | 0 | 0 | 0 | 0 | 0 |
| 871.4 | 2624 | 283 | 316 | 737 | 121 | 316 | 13 | 8.4 | 7.8 | 11 | 17 | 11 | 0.0023 | 0 | 0 | 0.0014 | 0 | 0 |
| 913 | 64 | 11 | 80 | 624 | 51 | 64 | 26 | 140 | 21 | 25 | 31 | 26 | 0 | 0 | 0 | 0.0064 | 0 | 0 |
| 938 | 33 | 11 | 72 | 275 | 4 | 33 | 170 | 180 | 28 | 7.1 | 460 | 170 | 0.030 | 0 | 0 | 0 | 0 | 0 |
| 960.1 | 5168 | 1462 | 966 | 4772 | 1605 | 1605 | 30 | 32 | 41 | 26 | 34 | 32 | 0.0017 | 0.0027 | 0.0041 | 0.0017 | 0.0031 | 0.0027 |
| 975 | 3875 | 2957 | 2800 | 4293 | 340 | 2957 | 17 | 44 | 24 | 26 | 110 | 26 | 0.0021 | 0.0037 | 0.0029 | 0.0026 | 0.015 | 0.0029 |
| 987 | 687 | 400 | 523 | 455 | 61 | 455 | 43 | 49 | 25 | 48 | 110 | 48 | 0.0073 | 0.0075 | 0.0038 | 0.0066 | 0.016 | 0.0073 |
| 989 | 527 | 1375 | 1570 | 515 | 17 | 527 | 90 | 53 | 48 | 59 | 320 | 59 | 0.0057 | 0.0029 | 0.0025 | 0.0039 | 0 | 0.0029 |

**Supplementary Table S4. Model performance by phecode for 151 rare phecodes in MSDW cohort.**

| **Phecode** | **Number of Positive Predictions** | **Predictive Odds Ratio** | **Positive Predictive Value** |
| --- | --- | --- | --- |
| 008.7 | 2125 | 35 | 0.0061 |
| 070.1 | 6713 | 9.7 | 0.0010 |
| 071 | 51801 | 97 | 0 |
| 071.1 | 16291 | 320 | 0 |
| 079.1 | 54185 | 7.1 | 0.0023 |
| 079.2 | 81022 | 14 | 0.010 |
| 110.11 | 15351 | 14 | 0.11 |
| 110.13 | 17013 | 9.5 | 0.038 |
| 110.2 | 29449 | 9.4 | 0.036 |
| 112.3 | 79198 | 17 | 0.020 |
| 130 | 1008 | 3.2 | 0.0055 |
| 145.1 | 6931 | 37 | 8.7E-04 |
| 149 | 13595 | 200 | 0.019 |
| 149.3 | 2332 | 1300 | 0.062 |
| 149.9 | 18683 | 170 | 0.019 |
| 164 | 5909 | 430 | 0.031 |
| 174.3 | 1366 | 60 | 0.044 |
| 187.1 | 2988 | 57 | 0.0066 |
| 187.8 | 2867 | 21 | 0.0030 |
| 189.12 | 16564 | 210 | 0.011 |
| 191.1 | 26696 | 55 | 0.0049 |
| 194 | 12939 | 83 | 0.0073 |
| 199.4 | 2514 | 170 | 0.040 |
| 204.11 | 39477 | 27 | 0.0037 |
| 204.3 | 4176 | 160 | 0.0076 |
| 224 | 439 | 120 | 0.018 |
| 224.1 | 949 | 54 | 0.046 |
| 225.2 | 40958 | 33 | 0.0027 |
| 242.2 | 14351 | 9.8 | 0.0017 |
| 242.3 | 11721 | 420 | 0 |
| 246.7 | 783436 | 4.0 | 0.0068 |
| 250.12 | 11468 | 160 | 0.054 |
| 250.3 | 61654 | 27 | 0.010 |
| 250.5 | 71614 | 3.8 | 0.0015 |
| 253 | 140370 | 26 | 0.011 |
| 253.11 | 9467 | 320 | 0.033 |
| 253.3 | 20658 | 98 | 0.016 |
| 255.11 | 29219 | 57 | 0.023 |
| 255.12 | 33902 | 56 | 0.010 |
| 258 | 11407 | 340 | 0.014 |

**Supplementary Table S4 (cont).**

| **Phecode** | **Number of Positive Predictions** | **Predictive Odds Ratio** | **Positive Predictive Value** |
| --- | --- | --- | --- |
| 261.41 | 12224 | 9.0 | 0.0033 |
| 272.12 | 8610 | 27 | 0.14 |
| 277.51 | 7490 | 6.8 | 0.0088 |
| 278.3 | 989 | 15 | 0.01 |
| 278.4 | 18366 | 14 | 0.11 |
| 286.11 | 229 | 12 | 0 |
| 286.13 | 27941 | 10 | 0.0018 |
| 286.5 | 32013 | 28 | 0.0017 |
| 288.3 | 23487 | 21 | 0.016 |
| 290.3 | 8284 | 69 | 0.012 |
| 297 | 37369 | 16 | 5.6E-04 |
| 300.4 | 11103 | 8.4 | 0.019 |
| 305.2 | 22625 | 9.8 | 0.021 |
| 315.3 | 1659 | 41 | 0.040 |
| 323.2 | 13372 | 11000 | 0.040 |
| 325 | 27424 | 50 | 0.0059 |
| 337.1 | 51266 | 25 | 0.0075 |
| 353.1 | 14409 | 32 | 0.033 |
| 353.2 | 46953 | 40 | 0.020 |
| 361.2 | 31520 | 120 | 0.0079 |
| 362.3 | 21627 | 240 | 0.0045 |
| 364.41 | 28362 | 64 | 0.022 |
| 366.1 | 3997 | 200 | 0.022 |
| 366.3 | 16436 | 140 | 0.0057 |
| 367.4 | 13475 | 20 | 0.06 |
| 377.1 | 12023 | 63 | 0.05 |
| 378 | 8767 | 73 | 0.075 |
| 388 | 13932 | 57 | 0.13 |
| 394.4 | 79742 | 86 | 0.016 |
| 401.2 | 73564 | 810 | 0.16 |
| 420.22 | 30468 | 39 | 0.0061 |
| 425.2 | 15804 | 120 | 0.062 |
| 425.8 | 20808 | 95 | 0.0021 |
| 426.8 | 192723 | 30 | 0.0094 |
| 441.2 | 14259 | 26 | 0.0066 |
| 446 | 63182 | 17 | 0.0031 |
| 459.1 | 17736 | 26 | 0.042 |
| 473.1 | 2718 | 33 | 0.065 |
| 500.1 | 120351 | 21 | 0.0019 |
| 519 | 14152 | 150 | 0.024 |

**Supplementary Table S4 (cont).**

| **Phecode** | **Number of Positive Predictions** | **Predictive Odds Ratio** | **Positive Predictive Value** |
| --- | --- | --- | --- |
| 523.1 | 5223 | 6.7 | 0.0025 |
| 527.1 | 739 | 16 | 0 |
| 527.7 | 12630 | 19 | 0.041 |
| 528.12 | 5825 | 12 | 0.013 |
| 528.41 | 279 | 23 | 0 |
| 573.2 | 6318 | 530 | 0.24 |
| 580.11 | 57559 | 220 | 0.0094 |
| 597.2 | 4039 | 220 | 6.0E-04 |
| 609.2 | 6456 | 31 | 0.050 |
| 611.1 | 10756 | 9.4 | 0.088 |
| 612.3 | 6692 | 4.3 | 0.0010 |
| 613.5 | 9188 | 13 | 0.073 |
| 620 | 20735 | 53 | 0.013 |
| 626.11 | 13912 | 7.2 | 0.19 |
| 626.4 | 18896 | 7.9 | 0.0089 |
| 643.1 | 74816 | 15 | 0.0092 |
| 647 | 55099 | 43 | 0.021 |
| 649 | 75788 | 67 | 0.099 |
| 656.2 | 4227 | 5.1 | 0.0063 |
| 686.5 | 431 | 350 | 0.057 |
| 690.1 | 5859 | 40 | 0.52 |
| 691 | 816 | 49 | 0.036 |
| 694.1 | 10840 | 7.3 | 0.010 |
| 695.21 | 14993 | 39 | 0.0017 |
| 695.81 | 19891 | 9.8 | 0.0017 |
| 696.3 | 14323 | 6.8 | 0.0087 |
| 701.1 | 14791 | 35 | 0.024 |
| 701.3 | 3493 | 49 | 0.038 |
| 701.6 | 20795 | 1.7 | 0.0023 |
| 704.8 | 10896 | 21 | 0.064 |
| 706.1 | 7158 | 10 | 0.18 |
| 709.5 | 17602 | 46 | 0.014 |
| 709.6 | 29989 | 80 | 0.023 |
| 712 | 16723 | 160 | 0.020 |
| 713.5 | 11310 | 110 | 0.010 |
| 722.3 | 95576 | 31 | 1.5E-04 |
| 722.8 | 112416 | 59 | 0.021 |
| 724.8 | 79283 | 27 | 6.5E-04 |
| 726.4 | 38736 | 5.6 | 0.017 |
| 727.2 | 6270 | 6.3 | 0.0012 |

**Supplementary Table S4 (cont).**

| **Phecode** | **Number of Positive Predictions** | **Predictive Odds Ratio** | **Positive Predictive Value** |
| --- | --- | --- | --- |
| 737 | 53640 | 9.0 | 0.0021 |
| 737.2 | 70937 | 6.3 | 2.9E-04 |
| 741.1 | 184 | 51 | 0 |
| 742 | 1373 | 69 | 0 |
| 743.4 | 26423 | 8.7 | 0.0074 |
| 748 | 11127 | 23 | 0.013 |
| 750.15 | 17533 | 8.0 | 8.9E-04 |
| 751.2 | 24526 | 11 | 0.0068 |
| 752.1 | 7378 | 20 | 0.051 |
| 753.1 | 7838 | 54 | 0.0081 |
| 755.6 | 725 | 14 | 0.0045 |
| 755.61 | 1063 | 27 | 0.014 |
| 756 | 674 | 7.5 | 0.0025 |
| 756.3 | 22599 | 17 | 0.0075 |
| 758 | 7334 | 44 | 0.017 |
| 758.1 | 3455 | 14 | 0.017 |
| 759.1 | 2818 | 22 | 0.0053 |
| 790.1 | 26351 | 18 | 0.014 |
| 794 | 204367 | 1.0 | 3.4E-04 |
| 797 | 29555 | 150 | 0.14 |
| 801.1 | 21158 | 4.1 | 7.5E-04 |
| 860 | 9092 | 1000 | 0.013 |
| 870.8 | 8299 | 2.3 | 5.4E-04 |
| 871.4 | 11089 | 13 | 0.0075 |
| 913 | 11024 | 3.5 | 0.0019 |
| 938 | 2488 | 26 | 0.018 |
| 960.1 | 96285 | 5.2 | 0.0034 |
| 975 | 84832 | 50 | 0 |
| 987 | 38751 | 3.0 | 0.0012 |
| 989 | 6892 | 13 | 0.0045 |

**Supplementary Table S5. Regression results for mortality, DALY, and related measures.** DALY = Disability Adjusted Life Years, YLD = Years Lost to Disability, YLL = Years of Life Lost. DALY, YLD, and YLL are tested by linear regression; mortality is tested by Cox proportional hazard regression. Coefficients reported are the effect of RarePT prediction as a binary variable on the listed outcome, controlling for age, sex, and self-reported ethnicity. “NaN” (not a number) indicates values that could not be calculated due to insufficient sample size in predicted cases; p=0 indicates a p-value below the numerical precision of Python’s statsmodels package (approximately 2 × 10^-308^).

| **Phecode** | **Coefficient (Std. Err.); p-value** | | | |
| --- | --- | --- | --- | --- |
|  | **DALY** | **YLD** | **YLL** | **Mortality** |
| 008.7 | 0.96 (0.076); p=9.7e-37 | 0.17 (0.0057); p=9.7e-37 | 0.80 (0.073); p=1.3e-27 | -0.14 (0.10); p=0.17 |
| 070.1 | 3.2 (0.070); p=0 | 0.18 (0.0053); p=0 | 3.0 (0.067); p=0 | 1.1 (0.050); p=1.4e-103 |
| 071 | 0.30 (0.059); p=2.5e-07 | 0.043 (0.0044); p=2.5e-07 | 0.26 (0.056); p=4.2e-06 | 0.46 (0.066); p=3.2e-12 |
| 071.1 | 0.26 (0.095); p=0.0067 | 0.043 (0.0072); p=0.0067 | 0.22 (0.092); p=0.018 | 0.71 (0.094); p=4.3e-14 |
| 079.1 | 1.8 (0.058); p=7.4e-201 | 0.17 (0.0044); p=7.4e-201 | 1.6 (0.056); p=6.5e-178 | 1.6 (0.036); p=0 |
| 079.2 | 1.8 (0.056); p=2.1e-236 | 0.16 (0.0042); p=2.1e-236 | 1.7 (0.053); p=8.3e-214 | 1.5 (0.036); p=0 |
| 110.11 | 1.0 (0.086); p=1.2e-32 | 0.17 (0.0065); p=1.2e-32 | 0.86 (0.083); p=3.5e-25 | 0.69 (0.071); p=1.7e-22 |
| 110.13 | 1.9 (0.056); p=1.4e-256 | 0.35 (0.0042); p=1.4e-256 | 1.6 (0.054); p=3.7e-186 | 1.4 (0.032); p=0 |
| 110.2 | 2.2 (0.046); p=0 | 0.37 (0.0034); p=0 | 1.9 (0.044); p=0 | 1.5 (0.028); p=0 |
| 112.3 | 3.3 (0.031); p=0 | 0.39 (0.0023); p=0 | 3.0 (0.030); p=0 | 1.6 (0.018); p=0 |
| 130 | 0.26 (0.29); p=0.38 | 0.024 (0.022); p=0.38 | 0.23 (0.28); p=0.40 | 0.70 (0.26); p=0.0065 |
| 145.1 | 0.41 (0.060); p=8.4e-12 | -0.021 (0.0045); p=8.4e-12 | 0.43 (0.057); p=7.3e-14 | -0.97 (0.10); p=3.2e-21 |
| 149 | 3.9 (0.034); p=0 | 0.18 (0.0026); p=0 | 3.7 (0.032); p=0 | 7.5e+10 (NaN); p=NaN |
| 149.3 | 3.2 (0.052); p=0 | 0.16 (0.0039); p=0 | 3.1 (0.050); p=0 | 1.8 (0.028); p=0 |
| 149.9 | 2.8 (0.048); p=0 | 0.12 (0.0036); p=0 | 2.7 (0.046); p=0 | 1.3 (0.031); p=0 |
| 164 | 3.9 (0.045); p=0 | 0.15 (0.0034); p=0 | 3.7 (0.043); p=0 | NaN (NaN); p=NaN |
| 174.3 | 0.046 (0.077); p=0.55 | -0.015 (0.0058); p=0.55 | 0.062 (0.074); p=0.40 | 0.017 (0.11); p=0.87 |
| 187.1 | 0.80 (0.064); p=2.0e-35 | 0.035 (0.0048); p=2.0e-35 | 0.76 (0.061); p=4.0e-35 | 0.75 (0.048); p=1.6e-54 |
| 187.8 | -0.30 (0.052); p=7.8e-09 | -0.045 (0.0039); p=7.8e-09 | -0.26 (0.050); p=3.1e-07 | -0.31 (0.064); p=1.6e-06 |
| 189.12 | 2.2 (0.052); p=0 | 0.15 (0.0039); p=0 | 2.1 (0.050); p=0 | 1.3 (0.030); p=0 |
| 191.1 | 3.2 (0.042); p=0 | 0.16 (0.0032); p=0 | 3.1 (0.041); p=0 | 1.9 (0.023); p=0 |
| 194 | 2.5 (0.098); p=6.8e-147 | 0.11 (0.0074); p=6.8e-147 | 2.4 (0.094); p=1.2e-145 | 1.9 (0.051); p=4.3e-291 |
| 199.4 | 2.9 (0.098); p=4.8e-188 | 0.14 (0.0074); p=4.8e-188 | 2.7 (0.094); p=4.2e-185 | 1.3 (0.068); p=1.6e-85 |
| 204.11 | 2.3 (0.032); p=0 | 0.12 (0.0024); p=0 | 2.1 (0.031); p=0 | 1.8 (0.019); p=0 |
| 204.3 | 2.4 (0.052); p=0 | 0.17 (0.0039); p=0 | 2.2 (0.050); p=0 | 1.8 (0.028); p=0 |

**Supplementary Table S5 (cont.)**

| **Phecode** | **Coefficient (Std. Err.); p-value** | | | |
| --- | --- | --- | --- | --- |
|  | **DALY** | **YLD** | **YLL** | **Mortality** |
| 224 | -0.15 (0.15); p=0.32 | 0.019 (0.011); p=0.32 | -0.17 (0.14); p=0.24 | -0.48 (0.21); p=0.024 |
| 224.1 | -0.40 (0.11); p=0.00017 | 0.050 (0.0079); p=0.00017 | -0.45 (0.10); p=1.0e-05 | -1.5 (0.23); p=1.0e-10 |
| 225.2 | 0.20 (0.061); p=0.00099 | 0.097 (0.0046); p=0.00099 | 0.10 (0.058); p=0.076 | 0.12 (0.074); p=0.11 |
| 242.2 | 1.5 (0.13); p=2.5e-33 | 0.27 (0.0097); p=2.5e-33 | 1.3 (0.12); p=5.2e-25 | 1.0 (0.095); p=1.4e-25 |
| 242.3 | -0.55 (0.070); p=2.4e-15 | -0.042 (0.0053); p=2.4e-15 | -0.51 (0.067); p=2.6e-14 | -1.1 (0.15); p=1.1e-14 |
| 246.7 | 1.7 (0.034); p=0 | 0.17 (0.0025); p=0 | 1.5 (0.032); p=0 | 1.2 (0.024); p=0 |
| 250.12 | 1.7 (0.057); p=1.9e-188 | 0.30 (0.0043); p=1.9e-188 | 1.4 (0.055); p=3.9e-137 | 1.6 (0.032); p=0 |
| 250.3 | 1.9 (0.03); p=0 | 0.26 (0.0022); p=0 | 1.7 (0.028); p=0 | 1.1 (0.021); p=0 |
| 250.5 | 0.34 (0.043); p=1.9e-15 | 0.050 (0.0033); p=1.9e-15 | 0.29 (0.041); p=1.6e-12 | 0.31 (0.071); p=1.0e-05 |
| 253 | 1.7 (0.043); p=0 | 0.24 (0.0032); p=0 | 1.5 (0.041); p=7.6e-276 | 0.96 (0.032); p=2.4e-199 |
| 253.11 | 0.87 (0.078); p=1.3e-28 | 0.15 (0.0059); p=1.3e-28 | 0.72 (0.075); p=1.4e-21 | 0.31 (0.081); p=0.00014 |
| 253.3 | 2.0 (0.043); p=0 | 0.29 (0.0032); p=0 | 1.8 (0.041); p=0 | 1.2 (0.028); p=0 |
| 255.11 | 2.5 (0.053); p=0 | 0.29 (0.0040); p=0 | 2.2 (0.051); p=0 | 1.2 (0.038); p=4.4e-220 |
| 255.12 | 0.84 (0.055); p=3.3e-53 | 0.13 (0.0041); p=3.3e-53 | 0.71 (0.053); p=2.7e-41 | 0.46 (0.05); p=3.2e-20 |
| 258 | 3.3 (0.066); p=0 | 0.23 (0.0050); p=0 | 3.1 (0.063); p=0 | 1.3 (0.043); p=4.7e-216 |
| 261.41 | 2.2 (0.067); p=3e-238 | 0.15 (0.0050); p=3e-238 | 2.1 (0.064); p=3.2e-224 | 1.4 (0.042); p=6e-259 |
| 270.35 | 2.9 (0.071); p=0 | 0.23 (0.0054); p=0 | 2.7 (0.069); p=0 | 1.5 (0.043); p=6.8e-261 |
| 272.12 | 1.6 (0.044); p=1.0e-291 | 0.26 (0.0033); p=1.0e-291 | 1.4 (0.042); p=1.3e-231 | 0.51 (0.036); p=7.9e-45 |
| 277.51 | 1.7 (0.07); p=2.2e-130 | 0.25 (0.0053); p=2.2e-130 | 1.4 (0.067); p=8.5e-103 | 1.4 (0.042); p=1.6e-240 |
| 278.3 | 0.23 (0.11); p=0.042 | 0.11 (0.0086); p=0.042 | 0.12 (0.11); p=0.27 | -0.69 (0.23); p=0.0028 |
| 278.4 | 2.6 (0.053); p=0 | 0.33 (0.0040); p=0 | 2.3 (0.051); p=0 | 1.3 (0.034); p=7.6e-292 |
| 281.9 | 2.7 (0.029); p=0 | 0.36 (0.0022); p=0 | 2.3 (0.028); p=0 | 1.5 (0.018); p=0 |
| 286.11 | 1.2 (0.10); p=2.1e-32 | 0.15 (0.0077); p=2.1e-32 | 1.1 (0.098); p=2e-27 | 0.58 (0.10); p=1.3e-08 |
| 286.13 | 1.1 (0.068); p=5e-56 | 0.16 (0.0052); p=5.0e-56 | 0.92 (0.066); p=2.9e-44 | 0.63 (0.065); p=2.6e-22 |
| 286.5 | 2.5 (0.042); p=0 | 0.28 (0.0032); p=0 | 2.2 (0.040); p=0 | 1.3 (0.026); p=0 |
| 288.3 | 2.1 (0.052); p=0 | 0.22 (0.0039); p=0 | 1.9 (0.050); p=1.2e-291 | 1.6 (0.029); p=0 |
| 289.9 | 3.1 (0.032); p=0 | 0.31 (0.0024); p=0 | 2.8 (0.031); p=0 | 1.6 (0.019); p=0 |
| 290.3 | 1.7 (0.034); p=0 | 0.38 (0.0025); p=0 | 1.4 (0.032); p=0 | 1.4 (0.023); p=0 |
| 297 | -0.20 (0.045); p=5.1e-06 | 0.048 (0.0034); p=5.1e-06 | -0.25 (0.043); p=4.4e-09 | 0.17 (0.057); p=0.0028 |

**Supplementary Table S5 (cont.)**

| **Phecode** | **Coefficient (Std. Err.); p-value** | | | |
| --- | --- | --- | --- | --- |
|  | **DALY** | **YLD** | **YLL** | **Mortality** |
| 300.4 | 0.76 (0.037); p=3.6e-95 | 0.28 (0.0027); p=3.6e-95 | 0.48 (0.035); p=1.0e-42 | 0.62 (0.038); p=3.0e-59 |
| 305.2 | 1.5 (0.044); p=1.0e-249 | 0.31 (0.0033); p=1.0e-249 | 1.2 (0.042); p=2.2e-169 | 1.3 (0.035); p=0 |
| 315.3 | 1.6 (0.09); p=1.9e-68 | 0.52 (0.0068); p=1.9e-68 | 1.1 (0.087); p=2.4e-34 | 1.6 (0.053); p=1.7e-202 |
| 323.2 | 0.42 (0.12); p=8.0e-04 | 0.13 (0.0094); p=8e-04 | 0.28 (0.12); p=0.018 | 0.67 (0.14); p=9.2e-07 |
| 325 | 2.6 (0.09); p=5.3e-183 | 0.20 (0.0068); p=5.3e-183 | 2.4 (0.087); p=9.1e-169 | 1.8 (0.054); p=6.1e-248 |
| 337.1 | 1.6 (0.032); p=0 | 0.30 (0.0024); p=0 | 1.3 (0.031); p=0 | 0.89 (0.023); p=0 |
| 353.1 | 0.090 (0.083); p=0.28 | 0.11 (0.0062); p=0.28 | -0.021 (0.080); p=0.79 | -0.31 (0.13); p=0.014 |
| 353.2 | -0.20 (0.046); p=8.9e-06 | 0.11 (0.0034); p=8.9e-06 | -0.31 (0.044); p=1.7e-12 | -0.80 (0.085); p=3.0e-21 |
| 361.2 | -0.64 (0.025); p=6.9e-145 | 0.018 (0.0019); p=6.9e-145 | -0.66 (0.024); p=5.5e-166 | -1.2 (0.049); p=2.8e-138 |
| 362.3 | -0.24 (0.032); p=2.0e-14 | 0.071 (0.0024); p=2.0e-14 | -0.31 (0.031); p=9.3e-25 | -0.26 (0.040); p=6.6e-11 |
| 364.41 | -0.74 (0.027); p=1.1e-172 | -0.0083 (0.0020); p=1.1e-172 | -0.74 (0.026); p=5.4e-183 | -2 (0.076); p=8.5e-146 |
| 366.1 | -0.31 (0.077); p=7.4e-05 | 0.074 (0.0058); p=7.4e-05 | -0.38 (0.074); p=3.1e-07 | -0.74 (0.13); p=3.6e-09 |
| 366.3 | -0.60 (0.053); p=2.6e-29 | 0.026 (0.0040); p=2.6e-29 | -0.62 (0.051); p=2.6e-34 | -1.0 (0.10); p=7.6e-25 |
| 367.4 | -0.78 (0.042); p=1.5e-76 | 0.024 (0.0032); p=1.5e-76 | -0.80 (0.040); p=7.6e-88 | -2.1 (0.13); p=5.5e-59 |
| 377.1 | 0.51 (0.060); p=2.4e-17 | 0.18 (0.0045); p=2.4e-17 | 0.32 (0.057); p=1.5e-08 | 0.35 (0.057); p=5.8e-10 |
| 378 | 0.55 (0.085); p=8.2e-11 | 0.14 (0.0064); p=8.2e-11 | 0.41 (0.082); p=4.7e-07 | 0.35 (0.082); p=2.0e-05 |
| 388 | -0.51 (0.095); p=7e-08 | -0.032 (0.0072); p=7.0e-08 | -0.48 (0.091); p=1.4e-07 | -1.0 (0.20); p=9.3e-07 |
| 394.4 | 1.6 (0.034); p=0 | 0.25 (0.0025); p=0 | 1.4 (0.032); p=0 | 0.78 (0.026); p=1.3e-201 |
| 401.2 | 2.3 (0.024); p=0 | 0.32 (0.0018); p=0 | 2.0 (0.023); p=0 | 1.4 (0.015); p=0 |
| 420.22 | 1.5 (0.04); p=3.1e-291 | 0.17 (0.0030); p=3.1e-291 | 1.3 (0.039); p=1.1e-247 | 1.1 (0.027); p=0 |
| 425.2 | 1.3 (0.071); p=2.8e-78 | 0.14 (0.0053); p=2.8e-78 | 1.2 (0.068); p=1.2e-68 | 1.1 (0.047); p=1.2e-115 |
| 425.8 | 1.2 (0.057); p=4.5e-98 | 0.2 (0.0043); p=4.5e-98 | 1.0 (0.055); p=6.2e-75 | 1.3 (0.035); p=1.5e-291 |
| 426.8 | 1.4 (0.019); p=0 | 0.21 (0.0014); p=0 | 1.2 (0.018); p=0 | 0.94 (0.015); p=0 |
| 441.2 | 3.9 (0.038); p=0 | 0.34 (0.0028); p=0 | 3.5 (0.036); p=0 | 1.2 (0.025); p=0 |
| 446 | 1.1 (0.035); p=1.4e-209 | 0.15 (0.0027); p=1.4e-209 | 0.94 (0.034); p=1.5e-168 | 0.85 (0.028); p=3.3e-210 |
| 459.1 | 3.6 (0.074); p=0 | 0.33 (0.0056); p=0 | 3.2 (0.071); p=0 | 1.6 (0.040); p=0 |
| 473.1 | 0.34 (0.078); p=1.1e-05 | 0.11 (0.0059); p=1.1e-05 | 0.24 (0.075); p=0.0016 | 0.20 (0.079); p=0.011 |
| 500.1 | 1.8 (0.034); p=0 | 0.23 (0.0025); p=0 | 1.6 (0.033); p=0 | 1.2 (0.022); p=0 |
| 519 | 2.1 (0.044); p=0 | 0.19 (0.0033); p=0 | 1.9 (0.042); p=0 | 1.1 (0.031); p=1.4e-257 |

**Supplementary Table S5 (cont.)**

| **Phecode** | **Coefficient (Std. Err.); p-value** | | | |
| --- | --- | --- | --- | --- |
|  | **DALY** | **YLD** | **YLL** | **Mortality** |
| 523.1 | 0.68 (0.14); p=8.8e-07 | 0.052 (0.010); p=8.8e-07 | 0.62 (0.13); p=2.3e-06 | 1.1 (0.11); p=2.0e-23 |
| 527.1 | 0.76 (0.23); p=0.00073 | 0.067 (0.017); p=0.00073 | 0.69 (0.22); p=0.0013 | 0.45 (0.24); p=0.054 |
| 527.7 | 1.4 (0.072); p=1.2e-85 | 0.17 (0.0055); p=1.2e-85 | 1.3 (0.070); p=3.3e-72 | 1.6 (0.043); p=0 |
| 528.12 | 2.5 (0.086); p=1.1e-190 | 0.25 (0.0065); p=1.1e-190 | 2.3 (0.082); p=7.3e-168 | 1.5 (0.054); p=6.2e-171 |
| 528.41 | 0.31 (0.16); p=0.050 | 0.041 (0.012); p=0.050 | 0.27 (0.15); p=0.076 | 0.30 (0.17); p=0.074 |
| 573.2 | 5.4 (0.063); p=0 | 0.31 (0.0048); p=0 | 5.1 (0.060); p=0 | 2.0 (0.032); p=0 |
| 580.11 | 2.1 (0.035); p=0 | 0.27 (0.0026); p=0 | 1.8 (0.034); p=0 | 1.5 (0.021); p=0 |
| 588 | 2.5 (0.032); p=0 | 0.32 (0.0024); p=0 | 2.2 (0.031); p=0 | 1.7 (0.018); p=0 |
| 597.2 | 2.1 (0.050); p=0 | 0.19 (0.0038); p=0 | 1.9 (0.048); p=0 | 1.3 (0.032); p=0 |
| 609.2 | 0.044 (0.066); p=0.51 | 0.052 (0.0005); p=0.51 | -0.0079 (0.063); p=0.90 | -0.50 (0.082); p=1.3e-09 |
| 611.1 | 0.48 (0.068); p=1.5e-12 | 0.043 (0.0051); p=1.5e-12 | 0.44 (0.066); p=1.9e-11 | 0.74 (0.070); p=7.4e-26 |
| 612.3 | 0.078 (0.064); p=0.23 | 0.075 (0.0049); p=0.23 | 0.0027 (0.062); p=0.97 | -0.92 (0.16); p=1.3e-08 |
| 613.5 | 1.0 (0.044); p=2.5e-113 | 0.23 (0.0033); p=2.5e-113 | 0.77 (0.042); p=4.2e-74 | 0.52 (0.048); p=5.1e-27 |
| 620 | -0.24 (0.052); p=2.0e-06 | -0.021 (0.0039); p=2.0e-06 | -0.22 (0.050); p=6.0e-06 | -0.73 (0.12); p=2.0e-09 |
| 626.11 | -0.24 (0.046); p=1.3e-07 | -0.025 (0.0034); p=1.3e-07 | -0.22 (0.044); p=8.7e-07 | -1.0 (0.14); p=7.0e-14 |
| 626.4 | -0.37 (0.031); p=6.8e-34 | -0.023 (0.0023); p=6.8e-34 | -0.35 (0.030); p=1.9e-32 | -1.8 (0.13); p=1.2e-42 |
| 643.1 | -0.046 (0.026); p=0.075 | -0.035 (0.0020); p=0.075 | -0.012 (0.025); p=0.64 | -1.1 (0.12); p=7.2e-20 |
| 647 | -0.054 (0.031); p=0.083 | -0.035 (0.0024); p=0.083 | -0.019 (0.030); p=0.53 | -1.4 (0.18); p=1.2e-15 |
| 649 | -0.019 (0.029); p=0.51 | -0.029 (0.0022); p=0.51 | 0.010 (0.027); p=0.70 | -1.3 (0.15); p=1.2e-17 |
| 656.2 | 2.0 (0.069); p=6.1e-184 | 0.29 (0.0052); p=6.1e-184 | 1.7 (0.067); p=5.8e-146 | NaN (NaN); p=NaN |
| 686.5 | 2.5 (0.13); p=5.9e-88 | 0.29 (0.0096); p=5.9e-88 | 2.3 (0.12); p=4.0e-75 | 1.6 (0.081); p=4.3e-86 |
| 690.1 | 1.5 (0.062); p=5.5e-134 | 0.25 (0.0047); p=5.5e-134 | 1.3 (0.059); p=1.7e-102 | 0.88 (0.048); p=2.2e-73 |
| 691 | 2.3 (0.11); p=2.2e-98 | 0.24 (0.0081); p=2.2e-98 | 2 (0.10); p=1.3e-85 | 1.3 (0.077); p=2.8e-59 |
| 694.1 | 1.6 (0.066); p=6.2e-133 | 0.26 (0.0049); p=6.2e-133 | 1.4 (0.063); p=1.1e-101 | 0.65 (0.061); p=1.7e-26 |
| 695.21 | 0.52 (0.066); p=3.1e-15 | 0.084 (0.0050); p=3.1e-15 | 0.44 (0.063); p=5.8e-12 | 0.49 (0.070); p=3.2e-12 |
| 695.81 | 1.3 (0.076); p=6.4e-63 | 0.17 (0.0057); p=6.4e-63 | 1.1 (0.073); p=6.0e-52 | 1.1 (0.068); p=5.1e-56 |
| 696.3 | 0.019 (0.091); p=0.83 | 0.028 (0.0069); p=0.83 | -0.0085 (0.088); p=0.92 | -0.58 (0.15); p=0.00011 |
| 701.1 | 0.66 (0.066); p=4.7e-23 | 0.1 (0.0050); p=4.7e-23 | 0.56 (0.064); p=2.5e-18 | 0.20 (0.069); p=0.0041 |
| 701.3 | 0.62 (0.11); p=2.0e-08 | 0.096 (0.0083); p=2.0e-08 | 0.52 (0.11); p=7.9e-07 | 0.53 (0.10); p=1.9e-07 |

**Supplementary Table S5 (cont.)**

| **Phecode** | **Coefficient (Std. Err.); p-value** | | | |
| --- | --- | --- | --- | --- |
|  | **DALY** | **YLD** | **YLL** | **Mortality** |
| 701.6 | 0.034 (0.099); p=0.73 | -0.019 (0.0074); p=0.73 | 0.052 (0.095); p=0.58 | -1.4 (0.22); p=4.7e-10 |
| 704.8 | -0.024 (0.11); p=0.83 | -0.0028 (0.0085); p=0.83 | -0.021 (0.11); p=0.84 | -1.4 (0.26); p=9e-08 |
| 706.1 | 0.021 (0.12); p=0.86 | 0.08 (0.0091); p=0.86 | -0.059 (0.12); p=0.61 | -0.43 (0.19); p=0.02 |
| 709.5 | 0.12 (0.077); p=0.13 | 0.062 (0.0058); p=0.13 | 0.054 (0.074); p=0.47 | 0.22 (0.088); p=0.011 |
| 709.6 | 0.33 (0.074); p=9.6e-06 | 0.081 (0.0056); p=9.6e-06 | 0.25 (0.071); p=0.00052 | 0.46 (0.079); p=4e-09 |
| 712 | 1.6 (0.06); p=4e-151 | 0.22 (0.0045); p=4e-151 | 1.4 (0.058); p=8.6e-122 | 1.3 (0.039); p=1.6e-251 |
| 713.5 | 1.6 (0.054); p=9e-206 | 0.33 (0.004); p=9e-206 | 1.3 (0.052); p=1.8e-142 | 1.5 (0.031); p=0 |
| 722.3 | 0.38 (0.026); p=2.2e-48 | 0.17 (0.0019); p=2.2e-48 | 0.21 (0.025); p=3.5e-17 | 0.028 (0.032); p=0.38 |
| 722.8 | -0.24 (0.031); p=1.5e-14 | 0.1 (0.0024); p=1.5e-14 | -0.35 (0.03); p=2.3e-30 | -0.85 (0.062); p=5.7e-42 |
| 724.8 | 0.12 (0.034); p=0.00035 | 0.13 (0.0025); p=0.00035 | -0.0089 (0.032); p=0.78 | -0.2 (0.046); p=1.2e-05 |
| 726.4 | -0.45 (0.039); p=6.7e-31 | 0.045 (0.0029); p=6.7e-31 | -0.5 (0.038); p=5.6e-40 | -1.2 (0.09); p=8.6e-41 |
| 727.2 | -0.61 (0.056); p=3.7e-27 | -0.017 (0.0043); p=3.7e-27 | -0.59 (0.054); p=9.2e-28 | -1.2 (0.13); p=2e-20 |
| 737 | 1.6 (0.032); p=0 | 0.34 (0.0024); p=0 | 1.3 (0.031); p=0 | 0.8 (0.027); p=1.8e-196 |
| 737.2 | 0.53 (0.03); p=2.4e-71 | 0.22 (0.0022); p=2.4e-71 | 0.32 (0.029); p=2e-28 | 0.18 (0.034); p=1.4e-07 |
| 741.1 | 0.46 (0.079); p=4.1e-09 | 0.15 (0.0059); p=4.1e-09 | 0.32 (0.076); p=2.9e-05 | -0.14 (0.1); p=0.18 |
| 742 | -0.66 (0.044); p=1.3e-49 | -0.028 (0.0033); p=1.3e-49 | -0.63 (0.043); p=2.3e-49 | -0.99 (0.084); p=3.4e-32 |
| 743.4 | 1.4 (0.043); p=2.8e-246 | 0.3 (0.0032); p=2.8e-246 | 1.1 (0.041); p=3.1e-166 | 1.2 (0.027); p=0 |
| 748 | 1.9 (0.093); p=2.5e-93 | 0.095 (0.007); p=2.5e-93 | 1.8 (0.089); p=2.1e-91 | 0.87 (0.07); p=2.8e-36 |
| 750.1 | 1 (0.052); p=4.3e-91 | 0.2 (0.0039); p=4.3e-91 | 0.85 (0.05); p=1e-65 | -0.086 (0.059); p=0.14 |
| 750.15 | 1.9 (0.039); p=0 | 0.27 (0.0029); p=0 | 1.6 (0.037); p=0 | 0.53 (0.033); p=3.7e-59 |
| 751.2 | 0.5 (0.046); p=7.7e-27 | 0.11 (0.0035); p=7.7e-27 | 0.38 (0.044); p=4e-18 | -0.009 (0.049); p=0.85 |
| 752.1 | 0.25 (0.099); p=0.011 | 0.23 (0.0074); p=0.011 | 0.019 (0.095); p=0.84 | -0.019 (0.13); p=0.89 |
| 753.1 | -0.65 (0.065); p=1.9e-23 | 0.069 (0.0049); p=1.9e-23 | -0.72 (0.063); p=1.5e-30 | -1.6 (0.16); p=1.9e-23 |
| 755.6 | -0.31 (0.13); p=0.016 | 0.028 (0.0096); p=0.016 | -0.34 (0.12); p=0.0062 | -0.87 (0.25); p=0.00052 |
| 755.61 | -0.53 (0.11); p=1.5e-06 | -0.0051 (0.0084); p=1.5e-06 | -0.53 (0.11); p=6.8e-07 | -1.1 (0.28); p=3.6e-05 |
| 756 | 0.12 (0.13); p=0.37 | 0.15 (0.01); p=0.37 | -0.027 (0.13); p=0.83 | -0.46 (0.22); p=0.04 |
| 756.3 | 0.39 (0.069); p=1.1e-08 | 0.23 (0.0052); p=1.1e-08 | 0.16 (0.066); p=0.015 | -0.58 (0.13); p=1.2e-05 |
| 758 | 0.72 (0.066); p=8.1e-28 | 0.1 (0.0049); p=8.1e-28 | 0.62 (0.063); p=1.4e-22 | 0.23 (0.065); p=0.00031 |
| 758.1 | 1.6 (0.11); p=2.6e-48 | 0.21 (0.0082); p=2.6e-48 | 1.4 (0.1); p=1.2e-39 | 1 (0.082); p=1e-35 |

**Supplementary Table S5 (cont.)**

| **Phecode** | **Coefficient (Std. Err.); p-value** | | | |
| --- | --- | --- | --- | --- |
|  | **DALY** | **YLD** | **YLL** | **Mortality** |
| 759.1 | 0.048 (0.11); p=0.66 | 0.078 (0.0081); p=0.66 | -0.03 (0.1); p=0.77 | -0.47 (0.17); p=0.0071 |
| 790.1 | 1.9 (0.06); p=1.1e-218 | 0.28 (0.0045); p=1.1e-218 | 1.6 (0.058); p=4.7e-174 | 1 (0.045); p=2.1e-116 |
| 794 | -0.039 (0.034); p=0.25 | -0.039 (0.0025); p=0.25 | 6.9e-05 (0.032); p=1 | -0.91 (0.15); p=5.7e-10 |
| 797 | 4.3 (0.031); p=0 | 0.27 (0.0023); p=0 | 4 (0.029); p=0 | NaN (NaN); p=NaN |
| 801.1 | -0.31 (0.043); p=7.1e-13 | 0.079 (0.0032); p=7.1e-13 | -0.39 (0.041); p=6.1e-21 | -0.17 (0.056); p=0.0023 |
| 860 | 2.9 (0.053); p=0 | 0.21 (0.004); p=0 | 2.7 (0.051); p=0 | NaN (NaN); p=NaN |
| 870.8 | 0.079 (0.13); p=0.54 | 0.06 (0.0096); p=0.54 | 0.019 (0.12); p=0.88 | 0.42 (0.15); p=0.0047 |
| 871.4 | 0.42 (0.077); p=5.5e-08 | 0.084 (0.0058); p=5.5e-08 | 0.34 (0.074); p=6e-06 | 0.63 (0.066); p=7.2e-22 |
| 913 | -0.33 (0.24); p=0.17 | -0.042 (0.018); p=0.17 | -0.29 (0.23); p=0.21 | -1.5 (0.71); p=0.034 |
| 938 | 1.3 (0.2); p=4.2e-11 | 0.11 (0.015); p=4.2e-11 | 1.2 (0.19); p=3.6e-10 | 1 (0.14); p=1e-12 |
| 960.1 | 1.2 (0.034); p=8.9e-292 | 0.28 (0.0025); p=8.9e-292 | 0.97 (0.033); p=3.9e-193 | 1.2 (0.025); p=0 |
| 975 | 1.5 (0.034); p=0 | 0.33 (0.0025); p=0 | 1.2 (0.032); p=2.2e-280 | 1.2 (0.023); p=0 |
| 987 | 1.1 (0.074); p=7.1e-55 | 0.39 (0.0055); p=7.1e-55 | 0.76 (0.071); p=1.1e-26 | 1.2 (0.06); p=3.5e-91 |
| 989 | 1.6 (0.069); p=1.7e-122 | 0.47 (0.0051); p=1.7e-122 | 1.1 (0.066); p=4.6e-67 | 1.6 (0.047); p=3.5e-240 |

**Supplementary Table S6. List of diagnostic tests relevant to rare phecodes.** Relationships are derived from the SNOMED-CT “interpreted as” annotation; see Methods for details.

| **Phe-code** | **Phecode description** | **UK Biobank field** | **Expected Direction** | **ICD-10-CM** | **ICD-10-CM description** | **SNOMED-CT** | **SNOMED-CT description** |
| --- | --- | --- | --- | --- | --- | --- | --- |
| 071.1 | HIV infection, symptomatic | Haemo-globin (30020) | below | B20 | Human immunodeficiency virus [HIV] disease | 420543008 | Anemia associated with AIDS |
|  |  |  |  |  |  | 421102007 | Aplastic anemia with AIDS (acquired immunodeficiency syndrome) |
|  |  |  |  |  |  | 421851008 | Acquired hemolytic anemia associated with acquired immunodeficiency syndrome (disorder) |
|  |  |  |  |  |  | 713349004 | Anaemia co-occurrent with human immunodeficiency virus infection |
|  |  |  |  |  |  | 713508003 | Aplastic anaemia co-occurrent with human immunodeficiency virus infection |
|  |  |  |  |  |  | 713533000 | Acquired hemolytic anemia co-occurrent with human immunodeficiency virus infection (disorder) |
|  |  | Neutro-phill count (30140) | below |  |  | 416729007 | Neutropenia with AIDS (acquired immunodeficiency syndrome) |
|  |  | Platelet count (30080) | below |  |  | 421102007 | Aplastic anemia with AIDS (acquired immunodeficiency syndrome) |
|  |  |  |  |  |  | 421766003 | Thrombocytopenia associated with AIDS (disorder) |
|  |  |  |  |  |  | 713508003 | Aplastic anaemia co-occurrent with human immunodeficiency virus infection |

**Supplementary Table S6 (cont).**

| **Phe-code** | **Phecode description** | **UK Biobank field** | **Expected Direction** | **ICD-10-CM** | **ICD-10-CM description** | **SNOMED-CT** | **SNOMED-CT description** |
| --- | --- | --- | --- | --- | --- | --- | --- |
| 071.1 (cont.) | HIV infection, symptomatic (cont.) | Red blood cell count (30010) | below | B20 | Human immunodeficiency virus [HIV] disease | 420543008 | Anemia associated with AIDS |
|  |  |  |  |  |  | 421102007 | Aplastic anemia with AIDS (acquired immunodeficiency syndrome) |
|  |  |  |  |  |  | 421851008 | Acquired hemolytic anemia associated with acquired immunodeficiency syndrome (disorder) |
|  |  |  |  |  |  | 713349004 | Anaemia co-occurrent with human immunodeficiency virus infection |
|  |  |  |  |  |  | 713508003 | Aplastic anaemia co-occurrent with human immunodeficiency virus infection |
|  |  |  |  |  |  | 713533000 | Acquired hemolytic anemia co-occurrent with human immunodeficiency virus infection (disorder) |
|  |  | White blood cell count (30000) |  |  |  | 421102007 | Aplastic anemia with AIDS (acquired immunodeficiency syndrome) |
|  |  |  |  |  |  | 713508003 | Aplastic anaemia co-occurrent with human immunodeficiency virus infection |
| 079.2 | Infectious mono-nucleosis | Haemo-globin (30020) | below | B27.9 | Infectious mononucleosis, unspecified | 127054006 | Cold agglutinin disease due to Epstein-Barr virus infection (disorder) |
|  |  | Red blood cell count (30010) | below |  |  |  |  |

**Supplementary Table S6 (cont).**

| **Phe-code** | **Phecode description** | **UK Biobank field** | **Expected Direction** | **ICD-10-CM** | **ICD-10-CM description** | **SNOMED-CT** | **SNOMED-CT description** |
| --- | --- | --- | --- | --- | --- | --- | --- |
| 250.12 | Type 1 diabetes with renal mani-festations | Albumin (30600) | below | E10.2 | Type 1 diabetes mellitus with kidney complications | 71721000119101 | Nephrotic syndrome due to type 1 diabetes mellitus (disorder) |
|  |  |  |  | E10.21 | Type 1 diabetes mellitus with diabetic nephropathy |  | Nephrotic syndrome due to type 1 diabetes mellitus (disorder) |
| 253 | Disorders of the pituitary gland and its hypo-thalamic control | Body mass index (21001) | above | E23.3 | Hypothalamic dysfunction, not elsewhere classified | 773663004 | Rapid-onset childhood obesity, hypothalamic dysfunction, hypoventilation, autonomic dysregulation syndrome |
|  |  |  |  | E23.6 | Other disorders of pituitary gland | 62999006 | Adiposogenital dystrophy (disorder) |
|  |  |  |  |  |  | 1187531009 | Obesity due to pituitary disease (disorder) |
|  |  |  |  | E23.7 | Disorder of pituitary gland, unspecified | 1187531009 | Obesity due to pituitary disease (disorder) |
| 277.51 | Lipoprotein disorders | Chol-esterol (30690) | above | E77.8 | Other disorders of glycoprotein metabolism | 1208738002 | TMEM199 congenital disorder of glycosylation |
| 278.3 | Localized adiposity | Body mass index (21001) | above | E65 | Localized adiposity | 57337005 | Steatopygia |
|  |  |  |  |  |  | 238135003 | Fat pad syndrome |
|  |  |  |  |  |  | 270486005 | Localized adiposity |

**Supplementary Table S6 (cont).**

| **Phe-code** | **Phecode description** | **UK Biobank field** | **Expected Direction** | **ICD-10-CM** | **ICD-10-CM description** | **SNOMED-CT** | **SNOMED-CT description** |
| --- | --- | --- | --- | --- | --- | --- | --- |
| 281.9 | Deficiency anemias | Haemo-globin (30020) | below | D53.9 | Nutritional anemia, unspecified | 48580008 | Anemia due to starvation |
|  |  |  |  |  |  | 52565000 | Non megaloblastic anaemia associated with nutritional deficiency |
|  |  |  |  |  |  | 66612000 | Nutritional anemia (disorder) |
|  |  |  |  |  |  | 83414005 | Macrocytic anemia |
|  |  |  |  |  |  | 86325007 | Non megaloblastic anemia due to alcoholism (disorder) |
|  |  |  |  |  |  | 234364007 | Combined deficiency anaemia |
|  |  |  |  |  |  | 267513007 | Deficiency anemias |
|  |  |  |  |  |  | 291262006 | Simple chronic anemia |
|  |  |  |  |  |  | 1142030006 | Nutritional anemia of pregnancy (disorder) |
|  |  |  |  |  |  | 1142062009 | Macrocytic anemia of pregnancy |
|  |  | Red blood cell count (30010) | below |  |  | 48580008 | Anemia due to starvation |
|  |  |  |  |  |  | 52565000 | Non megaloblastic anaemia associated with nutritional deficiency |
|  |  |  |  |  |  | 66612000 | Nutritional anemia (disorder) |
|  |  |  |  |  |  | 83414005 | Macrocytic anemia |
|  |  |  |  |  |  | 86325007 | Non megaloblastic anemia due to alcoholism (disorder) |
|  |  |  |  |  |  | 234364007 | Combined deficiency anaemia |
|  |  |  |  |  |  | 267513007 | Deficiency anemias |
|  |  |  |  |  |  | 291262006 | Simple chronic anemia |
|  |  |  |  |  |  | 1142030006 | Nutritional anemia of pregnancy (disorder) |
| 286.11 | Von Willebrand's disease | Platelet count (30080) | below | D69.8 | Other specified hemorrhagic conditions | 711407000 | Thrombocytopathy, asplenia and miosis (disorder) |
|  |  |  |  |  |  | 721119004 | Pseudothrombocytopenia (finding) |
|  |  |  |  |  |  | 783194008 | Bleeding diathesis due to thromboxane synthesis deficiency |

**Supplementary Table S6 (cont).**

| **Phe-code** | **Phecode description** | **UK Biobank field** | **Expected Direction** | **ICD-10-CM** | **ICD-10-CM description** | **SNOMED-CT** | **SNOMED-CT description** |
| --- | --- | --- | --- | --- | --- | --- | --- |
| 288.3 | Eosinophilia | Eosino-phill count (30150) | above | D72.1 | Eosinophilia | 30981000 | Secondary eosinophilia (disorder) |
|  |  |  |  |  |  | 64249002 | Allergic eosinophilia |
|  |  |  |  |  |  | 79336007 | Familial eosinophilia |
|  |  |  |  |  |  | 191358004 | Hereditary eosinophilia (disorder) |
|  |  |  |  |  |  | 191360002 | Drug-induced eosinophilia |
|  |  |  |  |  |  | 239910001 | Toxic oil syndrome (disorder) |
|  |  |  |  |  |  | 320150004 | Idiopathic eosinophilia (disorder) |
|  |  |  |  |  |  | 386789004 | Eosinophil count raised (finding) |
|  |  |  |  |  |  | 402404006 | Episodic angioedema with eosinophilia (disorder) |
|  |  |  |  |  |  | 419455006 | Disorder characterised by eosinophilia |
|  |  |  |  |  |  | 419769007 | Increased blood eosinophil number (finding) |
|  |  |  |  |  |  | 423486005 | Disseminated eosinophilic collagen disease |
|  |  |  |  |  |  | 722067005 | Severe combined immunodeficiency with hypereosinophilia (disorder) |
|  |  |  |  |  |  | 735441009 | Constitutional eosinophilia |
|  |  |  |  |  |  | 735442002 | Acquired eosinophilia |
|  |  |  |  |  |  | 860824009 | Eosinophilia due to infectious disease (disorder) |
|  |  |  |  |  |  | 267513007 | Deficiency anemias |
|  |  |  |  |  |  | 291262006 | Simple chronic anemia |
|  |  |  |  |  |  | 1142030006 | Nutritional anemia of pregnancy (disorder) |

**Supplementary Table S6 (cont).**

| **Phe-code** | **Phecode description** | **UK Biobank field** | **Expected Direction** | **ICD-10-CM** | **ICD-10-CM description** | **SNOMED-CT** | **SNOMED-CT description** |
| --- | --- | --- | --- | --- | --- | --- | --- |
| 289.9 | Abnormality of red blood cells | Haemat-ocrit (30030) | above | R71 | Abnormality of red blood cells | 165413005 | Hematocrit - PCV - high (finding) |
|  |  |  |  | R71.8 | Other abnormality of red blood cells |  |  |
|  |  |  | below | R71 | Abnormality of red blood cells | 165414004 | Hematocrit - packed cell volume - low (finding) |
|  |  |  |  | R71.0 | Precipitous drop in hematocrit |  |  |
|  |  |  | outside | R71 | Abnormality of red blood cells | 165416002 | Haematocrit - PCV abnormal |
|  |  | Haemo-globin (30020) | above | R71.8 | Other abnormality of red blood cells | 131141003 | Increased hemoglobin (finding) |
|  |  |  | below | D74.0 | Congenital methemoglobinemia | 47526003 | HNSHA due to NADH diaphorase deficiency (disorder) |
|  |  |  |  |  |  | 70517008 | HNSHA due to NADH-methemoglobin reductase deficiency (disorder) |
|  |  |  |  | R71 | Abnormality of red blood cells | 18662002 | Acquired Heinz body anaemia |
|  |  |  |  |  |  | 323666000 | Anemia due to intrinsic red cell abnormality (disorder) |
|  |  |  |  | R71.8 | Other abnormality of red blood cells | 5994005 | Hereditary elliptocytosis due to deficiency of protein 4.1 (disorder) |
|  |  |  |  |  |  | 8857001 | Hereditary elliptocytosis due to alpha spectrin defect (disorder) |
|  |  |  |  |  |  | 10564005 | Severe hereditary spherocytosis due to combined deficiency of spectrin AND ankyrin |
|  |  |  |  |  |  | 15121005 | Hereditary elliptocytosis due to glycophorin C deficiency (disorder) |

**Supplementary Table S6 (cont).**

| **Phe-code** | **Phecode description** | **UK Biobank field** | **Expected Direction** | **ICD-10-CM** | **ICD-10-CM description** | **SNOMED-CT** | **SNOMED-CT description** |
| --- | --- | --- | --- | --- | --- | --- | --- |
| 289.9 (cont.) | Abnormality of red blood cells (cont.) | Haemo-globin (30020) (cont.) | below (cont.) | R71.8 (cont.) | Other abnormality of red blood cells (cont.) | 15121005 | Hereditary elliptocytosis due to glycophorin C deficiency (disorder) |
|  |  |  |  |  |  | 24975009 | Mild hereditary spherocytosis due to combined deficiency of spectrin AND ankyrin (disorder) |
|  |  |  |  |  |  | 25266006 | Hereditary spherocytosis due to spectrin deficiency |
|  |  |  |  |  |  | 32648007 | Mild hereditary spherocytosis due to spectrin deficiency |
|  |  |  |  |  |  | 33905008 | Hereditary spherocytosis due to deficiency of protein 4.2 |
|  |  |  |  |  |  | 47516005 | Hereditary spherocytosis due to combined deficiency of spectrin AND ankyrin |
|  |  |  |  |  |  | 66262001 | Hereditary elliptocytosis due to beta spectrin-ankyrin interaction (disorder) |
|  |  |  |  |  |  | 69981004 | Hereditary spherocytosis due to beta spectrin defect |
|  |  |  |  |  |  | 73073009 | Hereditary elliptocytosis due to beta spectrin defect in self-association (disorder) |
|  |  |  |  |  |  | 75443009 | Hereditary elliptocytosis due to abnormal protein 4.1 (disorder) |
|  |  |  |  |  |  | 77413008 | Severe hereditary spherocytosis due to spectrin deficiency |
|  |  |  |  |  |  | 234410008 | Hereditary elliptocytosis with transient poikilocytosis |
|  |  |  | outside |  |  | 441793007 | Hemoglobin level outside reference range |

**Supplementary Table S6 (cont).**

| **Phe-code** | **Phecode description** | **UK Biobank field** | **Expected Direction** | **ICD-10-CM** | **ICD-10-CM description** | **SNOMED-CT** | **SNOMED-CT description** |
| --- | --- | --- | --- | --- | --- | --- | --- |
| 289.9 (cont.) | Abnormality of red blood cells (cont.) | Mean corpus-cular haemo-globin (30050) | above | R71 | Abnormality of red blood cells | 165440005 | MCH - raised |
|  |  |  |  | R71.8 | Other abnormality of red blood cells |  |  |
|  |  |  |  |  |  | 165448003 | Mean corpuscular hemoglobin concentration above reference range |
|  |  |  | below | R71 | Abnormality of red blood cells | 165439008 | MCH - low |
|  |  |  |  |  |  | 165447008 | Mean corpuscular haemoglobin concentration below reference range |
|  |  |  |  | R71.8 | Other abnormality of red blood cells | 165439008 | MCH - low |
|  |  |  |  |  |  | 165447008 | Mean corpuscular haemoglobin concentration below reference range |
|  |  |  | outside | R71 | Abnormality of red blood cells | 165442002 | Mean corpuscular hemoglobin abnormal (finding) |
|  |  |  |  | R71.8 | Other abnormality of red blood cells |  |  |
|  |  | Mean corpus-cular volume (30040) | above | R71 | Abnormality of red blood cells | 165454002 | MCV - raised (finding) |
|  |  |  |  | R71.8 | Other abnormality of red blood cells |  |  |
|  |  |  | below | R71 | Abnormality of red blood cells | 165455001 | Mean corpuscular volume below reference range (finding) |
|  |  |  |  | R71.8 | Other abnormality of red blood cells |  |  |

**Supplementary Table S6 (cont).**

| **Phe-code** | **Phecode description** | **UK Biobank field** | **Expected Direction** | **ICD-10-CM** | **ICD-10-CM description** | **SNOMED-CT** | **SNOMED-CT description** |
| --- | --- | --- | --- | --- | --- | --- | --- |
| 289.9 (cont.) | Abnormality of red blood cells (cont.) | Red blood cell count (30010) | above | D75.0 | Familial erythrocytosis | 17342003 | Familial erythrocytosis (disorder) |
|  |  |  |  |  |  | 127065001 | Familial erythrocytosis due to diphosphoglycerate mutase deficiency (disorder) |
|  |  |  |  |  |  | 127066000 | Familial polycythaemia vera |
|  |  |  |  |  |  | 770407006 | Chuvash erythrocytosis (disorder) |
|  |  |  |  |  |  | 1153349004 | Primary familial polycythemia due to erythropoietin receptor mutation |
|  |  |  |  |  |  | 1153350004 | Polycythaemia due to PHD2 mutation |
|  |  |  |  |  |  | 1153352007 | Polycythemia due to HIF2A mutation (disorder) |
|  |  |  |  | R71 | Abnormality of red blood cells | 165424007 | Red blood cell count above reference range |
|  |  |  |  | R71.8 | Other abnormality of red blood cells |  |  |
|  |  |  | below | D74.0 | Congenital methemoglobinemia | 47526003 | HNSHA due to NADH diaphorase deficiency (disorder) |
|  |  |  |  |  |  | 70517008 | HNSHA due to NADH-methemoglobin reductase deficiency (disorder) |
|  |  |  |  | R71 | Abnormality of red blood cells | 18662002 | Acquired Heinz body anaemia |
|  |  |  |  |  |  | 165423001 | Red blood cell count below reference range (finding) |
|  |  |  |  | R71.8 | Other abnormality of red blood cells | 5994005 | Hereditary elliptocytosis due to deficiency of protein 4.1 (disorder) |
|  |  |  |  |  |  | 8857001 | Hereditary elliptocytosis due to alpha spectrin defect (disorder) |
|  |  |  |  |  |  | 10564005 | Severe hereditary spherocytosis due to combined deficiency of spectrin AND ankyrin |

**Supplementary Table S6 (cont).**

| **Phe-code** | **Phecode description** | **UK Biobank field** | **Expected Direction** | **ICD-10-CM** | **ICD-10-CM description** | **SNOMED-CT** | **SNOMED-CT description** |
| --- | --- | --- | --- | --- | --- | --- | --- |
| 289.9 (cont.) | Abnormality of red blood cells (cont.) | Red blood cell count (30010) (cont.) | below (cont.) | R71.8 (cont.) | Other abnormality of red blood cells (cont.) | 15121005 | Hereditary elliptocytosis due to glycophorin C deficiency (disorder) |
|  |  |  |  |  |  | 24975009 | Mild hereditary spherocytosis due to combined deficiency of spectrin AND ankyrin (disorder) |
|  |  |  |  |  |  | 25266006 | Hereditary spherocytosis due to spectrin deficiency |
|  |  |  |  |  |  | 32648007 | Mild hereditary spherocytosis due to spectrin deficiency |
|  |  |  |  |  |  | 33905008 | Hereditary spherocytosis due to deficiency of protein 4.2 |
|  |  |  |  |  |  | 47516005 | Hereditary spherocytosis due to combined deficiency of spectrin AND ankyrin |
|  |  |  |  |  |  | 62574001 | Erythropenia |
|  |  |  |  |  |  | 66262001 | Hereditary elliptocytosis due to beta spectrin-ankyrin interaction (disorder) |
|  |  |  |  |  |  | 69981004 | Hereditary spherocytosis due to beta spectrin defect |
|  |  |  |  |  |  | 73073009 | Hereditary elliptocytosis due to beta spectrin defect in self-association (disorder) |
|  |  |  |  |  |  | 75443009 | Hereditary elliptocytosis due to abnormal protein 4.1 (disorder) |
|  |  |  |  |  |  | 77413008 | Severe hereditary spherocytosis due to spectrin deficiency |
|  |  |  |  |  |  | 165423001 | Red blood cell count below reference range (finding) |
|  |  |  |  |  |  | 234410008 | Hereditary elliptocytosis with transient poikilocytosis |

**Supplementary Table S6 (cont).**

| **Phe-code** | **Phecode description** | **UK Biobank field** | **Expected Direction** | **ICD-10-CM** | **ICD-10-CM description** | **SNOMED-CT** | **SNOMED-CT description** |
| --- | --- | --- | --- | --- | --- | --- | --- |
| 289.9 (cont.) | Abnormality of red blood cells (cont.) | Red blood cell count (30010) (cont.) | outside | R71 | Abnormality of red blood cells | 165427000 | RBC count abnormal (finding) |
|  |  |  |  | R71.8 | Other abnormality of red blood cells |  |  |
|  |  | Reticul-ocyte count (30250) | above | R71 | Abnormality of red blood cells | 46049004 | Reticulocytosis (finding) |
|  |  |  |  |  |  | 170824004 | Reticulocytosis after B12 (finding) |
|  |  |  | outside |  |  | 165685003 | Reticulocyte count outside reference range (finding) |
| 315.3 | Mental retardation | Body mass index (21001) | above | F78 | Other intellectual disabilities | 719160009 | Syndromic X-linked intellectual disability type 7 |
|  |  |  |  | F79 | Unspecified intellectual disabilities | 715628009 | MORM syndrome |
|  |  |  |  |  |  | 722037004 | Mental retardation, epileptic seizures, hypogonadism and hypogenitalism, microcephaly, obesity syndrome (disorder) |
|  |  |  |  |  |  | 724137002 | MOMO syndrome |
|  |  |  |  |  |  | 763350002 | Intellectual disability, obesity, brain malformation, facial dysmorphism syndrome |
|  |  |  |  |  |  | 770750002 | Intellectual disability, seizures, macrocephaly, obesity syndrome (disorder) |
|  |  |  |  |  |  | 774102003 | Intellectual disability, obesity, prognathism, eye and skin anomalies syndrome |
|  |  |  |  |  |  | 776204008 | Colobomatous microphthalmia, obesity, hypogenitalism, intellectual disability syndrome (disorder) |

**Supplementary Table S6 (cont).**

| **Phe-code** | **Phecode description** | **UK Biobank field** | **Expected Direction** | **ICD-10-CM** | **ICD-10-CM description** | **SNOMED-CT** | **SNOMED-CT description** |
| --- | --- | --- | --- | --- | --- | --- | --- |
| 315.3 (cont.) | Mental retardation (cont.) | Bone mineral density (78) | below | F79 | Unspecified intellectual disabilities | 732954002 | Osteopenia, intellectual disability, sparse hair syndrome |
|  |  | Haemo-globin (30020) | below | F79 | Unspecified intellectual disabilities | 715342005 | Alpha thalassemia X-linked intellectual disability syndrome |
|  |  |  |  |  |  | 720982007 | Alport syndrome, intellectual disability, midface hypoplasia, elliptocytosis syndrome (disorder) |
|  |  |  |  |  |  | 734349003 | Alpha-thalassemia intellectual disability syndrome linked to chromosome 16 (disorder) |
|  |  | Hearing (20019,20021) | below | F78 | Other intellectual disabilities | 722213009 | Severe X-linked intellectual disability Gustavson type |
|  |  | Neutro-phill count (30140) | below | F79 | Unspecified intellectual disabilities | 719156006 | X-linked intellectual disability with hypogammaglobulinemia and progressive neurological deterioration syndrome (disorder) |
|  |  | Red blood cell count (30010) | below | F79 | Unspecified intellectual disabilities | 715342005 | Alpha thalassemia X-linked intellectual disability syndrome |
|  |  |  |  |  |  | 720982007 | Alport syndrome, intellectual disability, midface hypoplasia, elliptocytosis syndrome (disorder) |
|  |  |  |  |  |  | 734349003 | Alpha-thalassemia intellectual disability syndrome linked to chromosome 16 (disorder) |

**Supplementary Table S6 (cont).**

| **Phe-code** | **Phecode description** | **UK Biobank field** | **Expected Direction** | **ICD-10-CM** | **ICD-10-CM description** | **SNOMED-CT** | **SNOMED-CT description** |
| --- | --- | --- | --- | --- | --- | --- | --- |
| 366.1 | Nonsenile Cataract | Height (50) | below | H26.009 | Unspecified infantile and juvenile cataract, unspecified eye | 1220597000 | Retinitis pigmentosa, juvenile cataract, short stature, intellectual disability syndrome (disorder) |
| 377.1 | Optic atrophy | Hearing (20019,20021) | below | H47.22 | Hereditary optic atrophy | 722213009 | Severe X-linked intellectual disability Gustavson type |
| 459.1 | Hemorrhage NOS | Platelet count (30080) | below | R58 | Hemorrhage, not elsewhere classified | 49886003 | Thrombocytopenia due to blood loss |
| 519 | Other diseases of respiratory system, not elsewhere classified | Red blood cell count (30010) | above | J98.9 | Respiratory disorder, unspecified | 191373003 | Polycythemia due to cyanotic respiratory disease |
| 588 | Disorders resulting from impaired renal function | Create-inine (30700) | outside | N25.9 | Disorder resulting from impaired renal tubular function, unspecified | 244941000119101 | Abnormal renal tubular function (finding) |
|  |  | Haemo-globin (30020) | below | N25.89 | Other disorders resulting from impaired renal tubular function | 726669007 | Central nervous system calcification, deafness, tubular acidosis, anemia syndrome |
|  |  | Red blood cell count (30010) | below |  |  |  |  |

**Supplementary Table S6 (cont).**

| **Phe-code** | **Phecode description** | **UK Biobank field** | **Expected Direction** | **ICD-10-CM** | **ICD-10-CM description** | **SNOMED-CT** | **SNOMED-CT description** |
| --- | --- | --- | --- | --- | --- | --- | --- |
| 588 (cont.) | Disorders resulting from impaired renal function (cont.) | Urate (30880) | above | M10.31 | Gout due to renal impairment, shoulder | 304281000119100 | Chronic tophaceous gout of shoulder due to renal impairment (disorder) |
|  |  |  |  |  |  | 304291000119102 | Chronic gout of shoulder without tophus due to renal impairment (disorder) |
|  |  |  |  | M10.32 | Gout due to renal impairment, elbow | 303901000119100 | Chronic tophaceous gout of elbow due to renal impairment (disorder) |
|  |  |  |  |  |  | 303911000119102 | Chronic gout of elbow without tophus due to renal impairment |
|  |  |  |  |  |  | 308791000119101 | Gout of elbow due to renal impairment |
|  |  |  |  | M10.33 | Gout due to renal impairment, wrist | 298961000119102 | Gout of wrist due to renal impairment |
|  |  |  |  |  |  | 304341000119104 | Chronic tophaceous gout of wrist due to renal impairment (disorder) |
|  |  |  |  |  |  | 304351000119102 | Chronic gout of wrist without tophus due to renal impairment (disorder) |
|  |  |  |  | M10.34 | Gout due to renal impairment, hand | 303921000119109 | Chronic tophaceous gout of hand due to renal impairment |
|  |  |  |  |  |  | 303931000119107 | Chronic gout of hand without tophus due to renal impairment |
|  |  |  |  |  |  | 308801000119100 | Gout of hand due to renal impairment |
|  |  |  |  | M10.35 | Gout due to renal impairment, hip | 303941000119103 | Chronic tophaceous gout of hip due to renal impairment |
|  |  |  |  |  |  | 303951000119101 | Chronic gout of hip without tophus due to renal impairment |
|  |  |  |  |  |  | 308811000119102 | Gout of hip due to renal impairment |
|  |  |  |  | M10.36 | Gout due to renal impairment, knee | 303961000119104 | Chronic tophaceous gout of knee due to renal impairment (disorder) |
|  |  |  |  |  |  | 303971000119105 | Chronic gout of knee without tophus due to renal impairment (disorder) |
|  |  |  |  |  |  | 308821000119109 | Gout of knee due to renal impairment (disorder) |

**Supplementary Table S6 (cont).**

| **Phe-code** | **Phecode description** | **UK Biobank field** | **Expected Direction** | **ICD-10-CM** | **ICD-10-CM description** | **SNOMED-CT** | **SNOMED-CT description** |
| --- | --- | --- | --- | --- | --- | --- | --- |
| 588 (cont.) | Disorders resulting from impaired renal function (cont.) | Urea (30670) | above | N25.89 | Other disorders resulting from impaired renal tubular function | 83850008 | Uraemic acidosis |
| 649 | Other conditions or status of the mother compli-cating pregnancy, childbirth, or the puerperium | Body mass index (21001) | above | O99.2 | Endocrine, nutritional and metabolic diseases complicating pregnancy, childbirth and the puerperium | 10750551000119100 | Obesity in mother complicating childbirth |
|  |  |  |  |  |  | 15750121000119108 | Severe obesity complicating pregnancy |
|  |  |  |  | O99.210 | Obesity complicating pregnancy, unspecified trimester | 171000119107 | Maternal obesity complicating pregnancy, childbirth and the puerperium, antepartum (disorder) |
|  |  |  |  |  |  | 15750121000119108 | Severe obesity complicating pregnancy |
|  |  |  |  | O99.214 | Obesity complicating childbirth | 10750551000119100 | Obesity in mother complicating childbirth |

**Supplementary Table S6 (cont).**

| **Phe-code** | **Phecode description** | **UK Biobank field** | **Expected Direction** | **ICD-10-CM** | **ICD-10-CM description** | **SNOMED-CT** | **SNOMED-CT description** |
| --- | --- | --- | --- | --- | --- | --- | --- |
| 686.5 | Pyoderma | Neutro-phill count (30140) | above | L08.0 | Pyoderma | 1254916008 | Hidradenitis suppurativa pyoderma gangrenosum complex (disorder) |
|  |  |  |  | L88 | Pyoderma gangrenosum | 74578003 | Pyoderma gangrenosum (disorder) |
|  |  |  |  |  |  | 238985006 | Bullous pyoderma (disorder) |
|  |  |  |  |  |  | 403702001 | Parastomal pyoderma gangrenosum |
|  |  |  |  |  |  | 724015007 | Pyogenic arthritis, pyoderma gangrenosum, acne syndrome (disorder) |
|  |  |  |  |  |  | 785724007 | Pyoderma gangrenosum, acne, suppurative hidradenitis syndrome (disorder) |
|  |  |  |  |  |  | 1256045009 | Superficial vegetating pyoderma gangrenosum (disorder) |
|  |  |  |  |  |  | 1256055008 | Ulcerative pyoderma gangrenosum |
|  |  |  |  |  |  | 1256078002 | Pyoderma gangrenosum due to inflammatory bowel disease (disorder) |
|  |  |  |  |  |  | 1256079005 | Pyoderma gangrenosum due to inflammatory polyarthropathy (disorder) |
|  |  |  |  |  |  | 1256081007 | Pustular pyoderma gangrenosum |
|  |  |  |  |  |  | 1258979003 | Pyoderma gangrenosum due to hematological disorder |

**Supplementary Table S6 (cont).**

| **Phe-code** | **Phecode description** | **UK Biobank field** | **Expected Direction** | **ICD-10-CM** | **ICD-10-CM description** | **SNOMED-CT** | **SNOMED-CT description** |
| --- | --- | --- | --- | --- | --- | --- | --- |
| 691 | Congenital anomalies of skin | Body mass index (21001) | above | Q82.9 | Congenital malformation of skin, unspecified | 774102003 | Intellectual disability, obesity, prognathism, eye and skin anomalies syndrome |
|  |  | Bone mineral density (78) | above | Q82.8 | Other specified congenital malformations of skin | 787408008 | Osteopathia striata, pigmentary dermopathy, white forelock syndrome (disorder) |
|  |  |  | below |  |  | 254116003 | Geroderma osteodysplastica (disorder) |
|  |  |  |  |  |  | 722113001 | Osteoporosis and oculocutaneous hypopigmentation syndrome |
|  |  |  |  |  |  | 778068007 | Autosomal recessive cutis laxa type 2B |
|  |  |  |  |  |  | 784381008 | Autosomal recessive cutis laxa type 2A (disorder) |
|  |  | Haemo-globin (30020) | below | Q82.8 | Other specified congenital malformations of skin | 60805002 | Haemolytic anaemia with emphysema AND cutis laxa |
|  |  |  |  |  |  | 723512008 | Revesz syndrome |
|  |  | Height (50) | below | Q82.9 | Congenital malformation of skin, unspecified | 1217229007 | Craniofacial dysplasia, short stature, ectodermal anomalies, intellectual disability syndrome (disorder) |
|  |  | Lympho-cyte count (30120) | below | Q82.8 | Other specified congenital malformations of skin | 1197428008 | Combined immunodeficiency, enteropathy spectrum (disorder) |
|  |  | Neutro-phill count (30140) | below | Q82.4 | Ectodermal dysplasia (anhidrotic) | 1003381002 | Onycho-tricho-dysplasia neutropenia syndrome (disorder) |

**Supplementary Table S6 (cont).**

| **Phe-code** | **Phecode description** | **UK Biobank field** | **Expected Direction** | **ICD-10-CM** | **ICD-10-CM description** | **SNOMED-CT** | **SNOMED-CT description** |
| --- | --- | --- | --- | --- | --- | --- | --- |
| 691 (cont.) | Congenital anomalies of skin (cont.) | Platelet count (30080) | below | Q82.8 | Other specified congenital malformations of skin | 723512008 | Revesz syndrome |
|  |  | Red blood cell count (30010) | below | Q82.8 | Other specified congenital malformations of skin | 60805002 | Haemolytic anaemia with emphysema AND cutis laxa |
|  |  |  |  |  |  | 723512008 | Revesz syndrome |
|  |  | White blood cell count (30000) | below |  |  |  |  |
| 704.8 | Other specified diseases of hair and hair follicles | Neutro-phill count (30140) | below | L67.8 | Other hair color and hair shaft abnormalities | 1003381002 | Onycho-tricho-dysplasia neutropenia syndrome (disorder) |
| 706.1 | Acne | Neutro-phill count (30140) | above | L70.0 | Acne vulgaris | 724015007 | Pyogenic arthritis, pyoderma gangrenosum, acne syndrome (disorder) |
|  |  |  | above | L70.9 | Acne, unspecified | 724015007 | Pyogenic arthritis, pyoderma gangrenosum, acne syndrome (disorder) |
|  |  |  |  |  |  | 785724007 | Pyoderma gangrenosum, acne, suppurative hidradenitis syndrome (disorder) |

**Supplementary Table S6 (cont).**

| **Phe-code** | **Phecode description** | **UK Biobank field** | **Dir-ection** | **ICD-10-CM** | **ICD-10-CM description** | **SNOMED-CT** | **SNOMED-CT description** |
| --- | --- | --- | --- | --- | --- | --- | --- |
| 709.6 | Other specified diffuse diseases of connective tissue | Eosino-phill count (30150) | above | M35.8 | Other specified systemic involvement of connective tissue | 95416007 | Eosinophilia myalgia syndrome |
|  |  |  |  |  |  | 239910001 | Toxic oil syndrome (disorder) |
|  |  |  |  |  |  | 403735006 | Eosinophilia-myalgia syndrome from tryptophan |
|  |  | Platelet count (30080) | below | M35.8 | Other specified systemic involvement of connective tissue | 1187615007 | TAFRO syndrome |
| 737 | Curvature of spine | Height (50) | below | Q67.5 | Congenital deformity of spine | 1197589000 | Steel syndrome |
| 755.61 | Congenital hip dysplasia and deformity | Height (50) | below | Q65.1 | Congenital dislocation of hip, bilateral | 1197589000 | Steel syndrome |
| 756 | Other congenital musculo-skeletal anomalies | Bone mineral density (78) | above | Q77.4 | Achondroplasia | 722114007 | Sclerosing dysplasia of bone, ichthyosis, premature ovarian failure syndrome (disorder) |
|  |  |  |  | Q77.8 | Other osteochondro-dysplasia with defects of growth of tubular bones and spine | 787408008 | Osteopathia striata, pigmentary dermopathy, white forelock syndrome (disorder) |
|  |  | Height (50) | above |  |  | 1169363004 | Overgrowth, metaphyseal undermodeling, spondylar dysplasia syndrome |
|  |  |  | below | Q68.1 | Congenital deformity of finger(s) and hand | 1187277001 | Short stature, brachydactyly, obesity, global developmental delay syndrome (disorder) |
|  |  |  |  | Q68.8 | Other specified congenital musculoskeletal deformities | 1197589000 | Steel syndrome |
|  |  |  |  | Q77.7 | Spondyloepiphyseal dysplasia | 1187303004 | Progressive spondyloepimetaphyseal dysplasia, short stature, short fourth metatarsals, intellectual disability syndrome (disorder) |

**Supplementary Table S6 (cont).**

| **Phe-code** | **Phecode description** | **UK Biobank field** | **Expected Direction** | **ICD-10-CM** | **ICD-10-CM description** | **SNOMED-CT** | **SNOMED-CT description** |
| --- | --- | --- | --- | --- | --- | --- | --- |
| 756.3 | Congenital anomalies of muscle, tendon, fascia, and connective tissue | Bone mineral density (78) | above | Q79.6 | Ehlers-Danlos syndrome | 720861000 | Ehlers-Danlos syndrome progeroid type (disorder) |
|  |  |  |  | Q79.8 | Other congenital malformations of musculoskeletal system | 9723006 | Hyperphosphatasaemia with bone disease |
|  |  |  |  |  |  | 254120004 | Dysplasia with increased bone density (disorder) |
|  |  |  |  |  |  | 254123002 | Dysosteosclerosis (disorder) |
|  |  |  |  |  |  | 254132000 | Endosteal hyperostoses with cerebellar hypoplasia (disorder) |
|  |  |  |  |  |  | 278833002 | Craniometadiaphyseal dysplasia |
|  |  |  | below | Q79.6 | Ehlers-Danlos syndrome | 1255121003 | Classical-like Ehlers-Danlos syndrome type 2 |
|  |  |  |  | Q79.8 | Other congenital malformations of musculoskeletal system | 254104009 | Dysplasia with decreased bone density |
|  |  |  |  |  |  | 254114000 | Singleton-Merten syndrome |
|  |  |  |  |  |  | 254116003 | Geroderma osteodysplastica (disorder) |
|  |  |  |  |  |  | 389199001 | Cole-Carpenter dysplasia |
|  |  |  |  |  |  | 722113001 | Osteoporosis and oculocutaneous hypopigmentation syndrome |
|  |  | Height (50) | below | Q79.6 | Ehlers-Danlos syndrome | 1251499005 | Beta-1,3-galactosyltransferase 6-related spondylodysplastic Ehlers-Danlos syndrome (disorder) |

**Supplementary Table S6 (cont).**

| **Phe-code** | **Phecode description** | **UK Biobank field** | **Expected Direction** | **ICD-10-CM** | **ICD-10-CM description** | **SNOMED-CT** | **SNOMED-CT description** |
| --- | --- | --- | --- | --- | --- | --- | --- |
| 758 | Chromo-somal anomalies and genetic disorders | Body mass index (21001) | above | Q87.1 | Congenital malformation syndromes predominantly associated with short stature | 770680004 | Prader-Willi-like syndrome (disorder) |
|  |  |  |  |  |  | 1229943004 | SIM1-related Prader-Willi-like syndrome |
|  |  |  |  |  |  | 1229946007 | MAGE family member L2-related Prader-Willi-like syndrome (disorder) |
|  |  |  |  |  |  | 1255335006 | X-linked intellectual disability, short stature, overweight syndrome |
|  |  | Height (50) | below |  |  | 1172605003 | Retinitis pigmentosa, hearing loss, premature aging, short stature, facial dysmorphism syndrome |
|  |  |  |  |  |  | 1187277001 | Short stature, brachydactyly, obesity, global developmental delay syndrome (disorder) |
|  |  |  |  |  |  | 1197592001 | Intrauterine growth restriction, short stature, early adult-onset diabetes syndrome (disorder) |
|  |  |  |  |  |  | 1217372003 | Severe myopia, generalized joint laxity, short stature syndrome |
|  |  |  |  |  |  | 1220596009 | Microcephalic primordial dwarfism, insulin resistance syndrome (disorder) |
|  |  |  |  |  |  | 1237618009 | Short stature, optic nerve atrophy, Pelger-HuÃ«t anomaly syndrome (disorder) |
|  |  |  |  |  |  | 1255335006 | X-linked intellectual disability, short stature, overweight syndrome |
| 758.1 | Chromo-somal anomalies | Body mass index (21001) | above | Q93.5 | Other deletions of part of a chromosome | 1003380001 | 6q16 microdeletion syndrome |
|  |  |  |  | Q93.88 | Other microdeletions |  |  |
|  |  |  |  | 99.8 | Other specified chromosome abnormalities | 700150001 | Congenital leptin deficiency |

**Supplementary Table S6 (cont).**

| **Phe-code** | **Phecode description** | **UK Biobank field** | **Expected Direction** | **ICD-10-CM** | **ICD-10-CM description** | **SNOMED-CT** | **SNOMED-CT description** |
| --- | --- | --- | --- | --- | --- | --- | --- |
| 758.1 (cont.) | Chromo-somal anomalies (cont.) | Bone mineral density (78) | above | Q93.5 | Other deletions of part of a chromosome | 719046005 | 12q14 microdeletion syndrome |
|  |  |  |  | Q93.88 | Other microdeletions | 719046005 | 12q14 microdeletion syndrome |
|  |  | Chol-esterol (30690) | above | Q99.8 | Other specified chromosome abnormalities | 403829002 | Familial hypercholesterolemia due to heterozygous LDL receptor mutation |
|  |  |  |  |  |  | 403830007 | Familial hypercholesterolaemia due to homozygous LDL receptor mutation |
|  |  | Haemo-globin (30020) | below | Q98.8 | Other specified sex chromosome abnormalities, male phenotype | 715342005 | Alpha thalassemia X-linked intellectual disability syndrome |
|  |  |  |  | Q99.9 | Chromosomal abnormality, unspecified | 734349003 | Alpha-thalassemia intellectual disability syndrome linked to chromosome 16 (disorder) |
|  |  | Height (50) | below | Q93.5 | Other deletions of part of a chromosome | 1228844002 | 1p35.2 microdeletion syndrome |
|  |  |  |  |  |  | 1228886008 | 9q33.3q34.11 microdeletion syndrome (disorder) |
|  |  |  |  |  |  | 1229882003 | 11q22.2q22.3 microdeletion syndrome (disorder) |
|  |  |  |  | Q93.88 | Other microdeletions | 1228844002 | 1p35.2 microdeletion syndrome |
|  |  |  |  |  |  | 1228886008 | 9q33.3q34.11 microdeletion syndrome (disorder) |
|  |  |  |  |  |  | 1229882003 | 11q22.2q22.3 microdeletion syndrome (disorder) |
|  |  |  |  | Q96.8 | Other variants of Turner's syndrome | 1237345002 | 46,XX ovarian dysgenesis, short stature syndrome |

**Supplementary Table S6 (cont).**

| **Phe-code** | **Phecode description** | **UK Biobank field** | **Expected Direction** | **ICD-10-CM** | **ICD-10-CM description** | **SNOMED-CT** | **SNOMED-CT description** |
| --- | --- | --- | --- | --- | --- | --- | --- |
| 758.1 (cont.) | Chromo-somal anomalies (cont.) | Neutro-phill count (30140) | below | Q99.8 | Other specified chromosome abnormalities | 234416002 | X-linked hypogammaglobulinaemia |
|  |  | Red blood cell count (30010) | below | Q98.8 | Other specified sex chromosome abnormalities, male phenotype | 715342005 | Alpha thalassemia X-linked intellectual disability syndrome |
|  |  |  |  | Q99.9 | Chromosomal abnormality, unspecified | 734349003 | Alpha-thalassemia intellectual disability syndrome linked to chromosome 16 (disorder) |
|  |  | White blood cell count (30000) | above | Q90.9 | Down syndrome, unspecified | 840505007 | Down syndrome co-occurrent with leukemoid reaction associated transient neonatal pustulosis |
| 794 | Abnormal results of other function studies (bladder, pancreas, placenta, spleen, etc) | Basal meta-bolic rate (23105) | above | R94.8 | Abnormal results of function studies of other organs and systems | 165110002 | Raised basal metabolic rate |
|  |  |  | outside | R94.8 | Abnormal results of function studies of other organs and systems | 442665002 | Basal metabolic rate outside reference range |
| 797 | Shock | Glucose (30740) | below | R57.8 | Other shock | 360546002 | Hypoglycemic shock (disorder) |

**Supplementary Table S6 (cont).**

| **Phe-code** | **Phecode description** | **UK Biobank field** | **Expected Direction** | **ICD-10-CM** | **ICD-10-CM description** | **SNOMED-CT** | **SNOMED-CT description** |
| --- | --- | --- | --- | --- | --- | --- | --- |
| 913 | Toxic effect of venom | Haemo-globin (30020) | below | T63.4 | Toxic effect of venom of other arthropods | 16645003 | Anemia due to insect venoms |
|  |  | Red blood cell count (30010) | below |  |  |  |  |
| 989 | Toxic effect of other substances, chiefly non-medicinal as to source | Haemo-globin (30020) | below | T65.9 | Toxic effect of unspecified substance | 424988008 | Anemia caused by substance (disorder) |
|  |  | Red blood cell count (30010) | below |  |  |  |  |

**Supplementary Table S7. Regression results for 75 diagnostic tests known to be relevant to rare phecodes.**

| **Phecode** | **Phenotype** | **Test** | **Direction** | **Coefficient (Std. Err.); p-value** | |
| --- | --- | --- | --- | --- | --- |
|  |  |  |  | **Logistic / binary** | **Linear / quantitative** |
| 071.1 | HIV infection, symptomatic | White blood cell count | below | 0.76 (0.17); p=1.1e-05 | 0.0083 (0.053); p=0.88 |
|  |  | Haemoglobin concentration | below | 0.64 (0.17); p=0.00019 | 0.23 (0.056); p=5.5e-05 |
|  |  | Platelet count | below | 1.3 (0.16); p=2.0e-17 | 0.057 (0.043); p=0.18 |
|  |  | Neutrophill count | below | 1.3 (0.28); p=2.5e-06 | 0.068 (0.042); p=0.11 |
|  |  | Red blood cell count | below | 1.0 (0.15); p=1.2e-11 | 0.21 (0.041); p=1.5e-07 |
| 079.2 | Infectious mononucleosis | Haemoglobin concentration | below | 0.91 (0.089); p=1.2e-24 | 0.36 (0.033); p=2.1e-27 |
|  |  | Red blood cell count | below | 1.3 (0.076); p=6.8e-62 | 0.32 (0.024); p=3.2e-40 |
| 250.12 | Type 1 diabetes with renal manifestations | Serum albumin | below | 3.1 (0.18); p=1.8e-64 | 0.68 (0.026); p=7.6e-156 |
| 253 | Disorders of the pituitary gland | Body mass index | above | 0.32 (0.034); p=3.8e-20 | 0.31 (0.030); p=4.1e-25 |
| 277.51 | Lipoprotein disorders | Total cholesterol | above | -0.88 (0.055); p=5.3e-57 | -1.1 (0.068); p=6.6e-63 |
| 278.3 | Localized adiposity | Body mass index | above | 0.45 (0.090); p=6.4e-07 | 0.48 (0.079); p=1.2e-09 |
| 281.9 | Deficiency anemias | Red blood cell count | below | 1.2 (0.043); p=7.9e-172 | 0.34 (0.013); p=1.1e-154 |
|  |  | Haemoglobin concentration | below | 1.4 (0.044); p=1.8e-207 | 0.60 (0.017); p=5.6e-260 |
| 286.11 | Von Willebrand's disease | Platelet count | below | 0.74 (0.23); p=0.0012 | -0.082 (0.046); p=0.074 |
| 288.3 | Eosinophilia | Eosinophill count | above | 0.77 (0.092); p=9.8e-17 | 0.29 (0.027); p=3.4e-26 |
| 289.9 | Abnormality of red blood cells | Haemoglobin concentration | outside | 0.084 (0.029); p=0.0040 | 0.24 (0.013); p=6.6e-72 |
|  |  | Mean corpuscular volume | outside | 0.94 (0.039); p=2.2e-131 | 0.3 (0.011); p=1.9e-171 |
|  |  | Red blood cell count | outside | 1.1 (0.045); p=5.8e-137 | 0.26 (0.0089); p=4.4e-189 |
|  |  | Reticulocyte count | outside | 0.37 (0.029); p=4.6e-37 | 0.18 (0.028); p=5.6e-11 |

**Supplementary Table S7 (cont).**

| **Phecode** | **Phenotype** | **Test** | **Direction** | **Coefficient (Std. Err.); p-value** | |
| --- | --- | --- | --- | --- | --- |
|  |  |  |  | **Logistic / binary** | **Linear / quantitative** |
| 289.9 | Abnormality of red blood cells | Mean corpuscular haemoglobin | outside | 0.22 (0.025); p=7.0e-18 | 0.39 (0.019); p=3.5e-90 |
|  |  | Haematocrit percentage | outside | 0.33 (0.027); p=2.4e-34 | 0.31 (0.013); p=1.5e-129 |
| 315.3 | Mental retardation | Haemoglobin concentration | below | 1.2 (0.13); p=4.1e-19 | 0.52 (0.054); p=1.5e-21 |
|  |  | Body mass index | above | 0.62 (0.070); p=1.0e-18 | 0.64 (0.064); p=1.4e-23 |
|  |  | Speech reception threshold (right ear) | above | 0.50 (0.12); p=5.5e-05 | 0.64 (0.11); p=9.9e-09 |
|  |  | Speech reception threshold (left ear) | above | 0.57 (0.12); p=3.4e-06 | 0.44 (0.11); p=8.7e-05 |
|  |  | Heel bone mineral density | below | 0.42 (0.10); p=2.2e-05 | 0.35 (0.11); p=0.00093 |
|  |  | Red blood cell count | below | 1.1 (0.13); p=1.5e-19 | 0.31 (0.040); p=1.7e-14 |
|  |  | Neutrophill count | below | -0.69 (0.71); p=0.33 | -0.39 (0.041); p=7.3e-22 |
| 366.1 | Nonsenile Cataract | Standing height | below | 0.26 (0.34); p=0.44 | -0.092 (0.032); p=0.0037 |
| 377.1 | Optic atrophy | Speech reception threshold (right ear) | above | 0.16 (0.084); p=0.061 | 0.18 (0.073); p=0.012 |
|  |  | Speech reception threshold (left ear) | above | 0.031 (0.085); p=0.72 | 0.14 (0.073); p=0.048 |
| 459.1 | Hemorrhage NOS | Platelet count | below | 1.0 (0.13); p=7.9e-16 | -0.019 (0.033); p=0.58 |
| 519 | Other diseases of respiratory system | Red blood cell count | above | 0.48 (0.26); p=0.060 | -0.017 (0.019); p=0.37 |
| 588 | Disorders resulting from impaired renal function | Serum creatinine | outside | 0.74 (0.029); p=2.9e-141 | 2.5 (0.025); p<2e-308 |
|  |  | Blood urea | above | 1.4 (0.033); p<2e-308 | 0.95 (0.016); p<2e-308 |
|  |  | Haemoglobin concentration | below | 1.4 (0.045); p=1.2e-209 | 0.61 (0.019); p=6e-221 |
|  |  | Serum urate | above | 0.57 (0.028); p=9.8e-89 | 0.6 (0.020); p=1.8e-200 |
|  |  | Red blood cell count | below | 1.3 (0.044); p=2.7e-197 | 0.36 (0.014); p=1.6e-149 |

**Supplementary Table S7 (cont).**

| **Phecode** | **Phenotype** | **Test** | **Direction** | **Coefficient (Std. Err.); p-value** | |
| --- | --- | --- | --- | --- | --- |
|  |  |  |  | **Logistic / binary** | **Linear / quantitative** |
| 649 | Other complications of pregnancy | Body mass index | above | -0.46 (0.028); p=3.1e-61 | -0.47 (0.020); p=1.1e-126 |
| 686.5 | Pyoderma | Neutrophill count | above | 1.2 (0.21); p=3.2e-09 | 0.28 (0.058); p=1.5e-06 |
| 691 | Congenital anomalies of skin | Red blood cell count | below | 0.68 (0.17); p=5.3e-05 | 0.12 (0.046); p=0.0097 |
|  |  | Neutrophill count | below | 0.42 (0.50); p=0.41 | -0.21 (0.048); p=1.6e-05 |
|  |  | Platelet count | below | 1 (0.21); p=9.3e-07 | 0.078 (0.048); p=0.11 |
|  |  | Standing height | below | 0.81 (0.39); p=0.036 | 0.14 (0.044); p=0.0011 |
|  |  | Lymphocyte count | below | 0.65 (0.19); p=0.00062 | -0.17 (0.092); p=0.061 |
|  |  | Body mass index | above | 0.48 (0.084); p=1.1e-08 | 0.57 (0.075); p=3.2e-14 |
|  |  | Haemoglobin concentration | below | 0.55 (0.18); p=0.0025 | 0.2 (0.063); p=0.0016 |
|  |  | Heel bone mineral density | outside | 0.38 (0.11); p=0.00041 | 0.3 (0.082); p=3.0e-04 |
|  |  | White blood cell count | below | 0.2 (0.26); p=0.45 | -0.26 (0.060); p=1.4e-05 |
| 704.8 | Other specified diseases of hair and follicles | Neutrophill count | below | 0.79 (0.45); p=0.079 | -0.0053 (0.050); p=0.92 |
| 706.1 | Acne | Neutrophil count | above | -0.015 (0.36); p=0.97 | 0.049 (0.053); p=0.36 |
| 709.6 | Other specified diffuse diseases of connective tissue | Eosinophil count | above | 0.33 (0.17); p=0.052 | 0.0071 (0.039); p=0.85 |
|  |  | Platelet count | below | 0.37 (0.21); p=0.077 | -0.0041 (0.033); p=0.90 |
| 737 | Curvature of spine | Standing height | below | 0.74 (0.11); p=6.4e-12 | 0.17 (0.013); p=2.6e-37 |
| 755.61 | Congenital hip dysplasia and deformity | Standing height | below | 0.45 (0.45); p=0.32 | -0.091 (0.046); p=0.047 |
| 756 | Other congenital musculoskeletal anomalies | Standing height | outside | 0.19 (0.23); p=0.41 | 0.043 (0.035); p=0.22 |
|  |  | Heel bone mineral density | above | 0.12 (0.21); p=0.58 | 0.091 (0.15); p=0.55 |

**Supplementary Table S7 (cont).**

| **Phecode** | **Phenotype** | **Test** | **Direction** | **Coefficient (Std. Err.); p-value** | |
| --- | --- | --- | --- | --- | --- |
|  |  |  |  | **Logistic / binary** | **Linear / quantitative** |
| 756.3 | Congenital anomalies of muscle and connective tissue | Standing height | below | 0.045 (0.31); p=0.88 | 0.12 (0.028); p=3.2e-05 |
|  |  | Heel bone mineral density | outside | 0.059 (0.071); p=0.40 | -0.024 (0.053); p=0.66 |
| 758 | Chromosomal anomalies and genetic disorders | Body mass index | above | -0.018 (0.056); p=0.75 | -0.031 (0.046); p=0.49 |
|  |  | Standing height | below | -0.36 (0.50); p=0.47 | -0.16 (0.027); p=4.4e-09 |
| 758.1 | Chromosomal anomalies | Haemoglobin concentration | below | 1.4 (0.15); p=2.6e-19 | 0.58 (0.064); p=3.9e-19 |
|  |  | Body mass index | above | 0.36 (0.087); p=4.1e-05 | 0.32 (0.076); p=1.9e-05 |
|  |  | Heel bone mineral density | above | -0.10 (0.17); p=0.55 | -0.33 (0.12); p=0.0076 |
|  |  | White blood cell count | above | 0.73 (0.19); p=0.00011 | 0.13 (0.061); p=0.026 |
|  |  | Total cholesterol | above | -0.52 (0.087); p=1.6e-09 | -0.64 (0.11); p=1.1e-09 |
|  |  | Red blood cell count | below | 1.5 (0.14); p=4.2e-26 | 0.38 (0.047); p=1.6e-15 |
|  |  | Neutrophil count | below | 0.95 (0.41); p=0.022 | -0.092 (0.048); p=0.058 |
|  |  | Standing height | below | 1.3 (0.39); p=0.00094 | 0.034 (0.045); p=0.45 |
| 794 | Abnormal results of other function studies | Basal metabolic rate | outside | -0.31 (0.028); p=9.6e-28 | -0.81 (0.036); p=4.5e-110 |
| 797 | Shock | Blood glucose | below | 0.028 (0.058); p=0.63 | -0.74 (0.038); p=8.2e-85 |
| 913 | Toxic effect of venom | Haemoglobin concentration | below | 0.05 (0.46); p=0.91 | -0.035 (0.14); p=0.80 |
|  |  | Red blood cell count | below | -0.13 (0.51); p=0.81 | -0.026 (0.10); p=0.80 |
| 989 | Toxic effect of other substances, chiefly nonmedicinal as to source | Haemoglobin concentration | below | 0.61 (0.12); p=3.3e-07 | 0.20 (0.040); p=9.7e-07 |
|  |  | Red blood cell count | below | 1.1 (0.097); p=6.9e-29 | 0.31 (0.030); p=2.5e-25 |

**Supplementary Table S8. Regression results for 100 permutations of phecode-test relationships.**

| **Permutation** | **Logistic / binary** | | | **Linear / quantitative** | | |
| --- | --- | --- | --- | --- | --- | --- |
|  | **# nominally significant** (*p* < 0.05) | **# Bonferroni significant**  (*p* < 0.00067) | **Median coefficient (log odds ratio)** | **# nominally significant**  (*p* < 0.05) | **# Bonferroni significant**  (*p* < 0.00067) | **Median coefficient (population std dev)** |
| *True data* | *54 (72%)* | *45 (60%)* | *0.57* | *43 (57%)* | *38 (51%)* | *0.17* |
| 1 | 28 (37%) | 18 (24%) | 0.14 | 22 (29%) | 14 (19%) | -0.022 |
| 2 | 30 (40%) | 22 (29%) | 0.14 | 24 (32%) | 15 (20%) | -9.9E-04 |
| 3 | 15 (20%) | 10 (13%) | 0.055 | 15 (20%) | 7 (9.3%) | -0.027 |
| 4 | 20 (27%) | 13 (17%) | 0.048 | 14 (19%) | 10 (13%) | -0.022 |
| 5 | 30 (40%) | 22 (29%) | 0.12 | 31 (41%) | 23 (31%) | 0.023 |
| 6 | 45 (60%) | 32 (43%) | 0.40 | 23 (31%) | 18 (24%) | -0.02 |
| 7 | 36 (48%) | 28 (37%) | 0.26 | 25 (33%) | 21 (28%) | 0.032 |
| 8 | 35 (47%) | 25 (33%) | 0.19 | 36 (48%) | 24 (32%) | 0.083 |
| 9 | 45 (60%) | 31 (41%) | 0.34 | 25 (33%) | 20 (27%) | -0.0047 |
| 10 | 27 (36%) | 24 (32%) | 0.056 | 22 (29%) | 15 (20%) | 0.0041 |
| 11 | 22 (29%) | 15 (20%) | 0.14 | 19 (25%) | 11 (15%) | -0.034 |
| 12 | 40 (53%) | 28 (37%) | 0.34 | 21 (28%) | 12 (16%) | -0.023 |
| 13 | 45 (60%) | 31 (41%) | 0.34 | 25 (33%) | 20 (27%) | -0.0047 |
| 14 | 45 (60%) | 37 (49%) | 0.53 | 27 (36%) | 16 (21%) | 0.039 |
| 15 | 28 (37%) | 18 (24%) | 0.14 | 22 (29%) | 14 (19%) | -0.022 |
| 16 | 36 (48%) | 28 (37%) | 0.26 | 25 (33%) | 21 (28%) | 0.032 |
| 17 | 16 (21%) | 11 (15%) | 0.051 | 15 (20%) | 10 (13%) | -0.027 |
| 18 | 39 (52%) | 33 (44%) | 0.56 | 31 (41%) | 19 (25%) | 0.020 |
| 19 | 36 (48%) | 28 (37%) | 0.26 | 25 (33%) | 21 (28%) | 0.032 |
| 20 | 22 (29%) | 15 (20%) | 0.14 | 19 (25%) | 11 (15%) | -0.034 |
| 21 | 39 (52%) | 29 (39%) | 0.22 | 39 (52%) | 24 (32%) | 0.089 |
| 22 | 31 (41%) | 23 (31%) | 0.17 | 17 (23%) | 10 (13%) | -0.039 |
| 23 | 24 (32%) | 21 (28%) | 0.13 | 14 (19%) | 11 (15%) | -0.023 |
| 24 | 35 (47%) | 25 (33%) | 0.19 | 36 (48%) | 24 (32%) | 0.083 |
| 25 | 42 (56%) | 37 (49%) | 0.48 | 20 (27%) | 12 (16%) | 0.0032 |
| 26 | 41 (55%) | 28 (37%) | 0.43 | 25 (33%) | 20 (27%) | 0.0093 |
| 27 | 28 (37%) | 25 (33%) | 0.044 | 22 (29%) | 19 (25%) | -0.033 |
| 28 | 28 (37%) | 18 (24%) | 0.14 | 22 (29%) | 14 (19%) | -0.022 |
| 29 | 24 (32%) | 17 (23%) | 0.051 | 28 (37%) | 23 (31%) | 0.0066 |
| 30 | 42 (56%) | 33 (44%) | 0.19 | 33 (44%) | 19 (25%) | 0.041 |
| 31 | 42 (56%) | 33 (44%) | 0.19 | 33 (44%) | 19 (25%) | 0.041 |
| 32 | 38 (51%) | 28 (37%) | 0.33 | 21 (28%) | 12 (16%) | -0.045 |
| 33 | 33 (44%) | 23 (31%) | 0.19 | 22 (29%) | 16 (21%) | 0.017 |
| 34 | 39 (52%) | 33 (44%) | 0.56 | 31 (41%) | 19 (25%) | 0.020 |
| 35 | 30 (40%) | 18 (24%) | 0.14 | 22 (29%) | 18 (24%) | -0.019 |
| 36 | 42 (56%) | 33 (44%) | 0.19 | 33 (44%) | 19 (25%) | 0.041 |

**Supplementary Table S8 (cont.)**

| **Permutation** | **Logistic / binary** | | | **Linear / quantitative** | | |
| --- | --- | --- | --- | --- | --- | --- |
|  | **# nominally significant** (*p* < 0.05) | **# Bonferroni significant**  (*p* < 0.00067) | **Median coefficient (log odds ratio)** | **# nominally significant**  (*p* < 0.05) | **# Bonferroni significant**  (*p* < 0.00067) | **Median coefficient (population std dev)** |
| *True data* | *54 (72%)* | *45 (60%)* | *0.57* | *43 (57%)* | *38 (51%)* | *0.17* |
| 37 | 24 (32%) | 17 (23%) | 0.051 | 28 (37%) | 23 (31%) | 0.0066 |
| 38 | 36 (48%) | 29 (39%) | 0.27 | 30 (40%) | 19 (25%) | 0.041 |
| 39 | 45 (60%) | 31 (41%) | 0.34 | 25 (33%) | 20 (27%) | -0.0047 |
| 40 | 34 (45%) | 27 (36%) | 0.15 | 31 (41%) | 23 (31%) | 0.047 |
| 41 | 40 (53%) | 27 (36%) | 0.31 | 31 (41%) | 23 (31%) | 0.053 |
| 42 | 28 (37%) | 18 (24%) | 0.14 | 22 (29%) | 14 (19%) | -0.022 |
| 43 | 18 (24%) | 14 (19%) | 0.023 | 18 (24%) | 14 (19%) | -0.034 |
| 44 | 36 (48%) | 29 (39%) | 0.27 | 30 (40%) | 19 (25%) | 0.041 |
| 45 | 41 (55%) | 34 (45%) | 0.60 | 32 (43%) | 20 (27%) | 0.039 |
| 46 | 23 (31%) | 17 (23%) | 0.13 | 16 (21%) | 11 (15%) | -0.027 |
| 47 | 45 (60%) | 31 (41%) | 0.34 | 25 (33%) | 20 (27%) | -0.0047 |
| 48 | 43 (57%) | 35 (47%) | 0.35 | 21 (28%) | 13 (17%) | -0.0034 |
| 49 | 34 (45%) | 28 (37%) | 0.21 | 26 (35%) | 22 (29%) | 0.039 |
| 50 | 21 (28%) | 17 (23%) | 0.11 | 14 (19%) | 11 (15%) | -0.03 |
| 51 | 39 (52%) | 33 (44%) | 0.56 | 31 (41%) | 19 (25%) | 0.02 |
| 52 | 13 (17%) | 8 (11%) | 0.019 | 11 (15%) | 7 (9.3%) | -0.018 |
| 53 | 36 (48%) | 28 (37%) | 0.26 | 25 (33%) | 21 (28%) | 0.032 |
| 54 | 36 (48%) | 28 (37%) | 0.26 | 25 (33%) | 21 (28%) | 0.032 |
| 55 | 45 (60%) | 37 (49%) | 0.53 | 27 (36%) | 16 (21%) | 0.039 |
| 56 | 20 (27%) | 16 (21%) | 0.049 | 16 (21%) | 12 (16%) | -0.014 |
| 57 | 22 (29%) | 15 (20%) | 0.06 | 12 (16%) | 8 (11%) | -0.022 |
| 58 | 42 (56%) | 33 (44%) | 0.19 | 33 (44%) | 19 (25%) | 0.041 |
| 59 | 31 (41%) | 22 (29%) | 0.15 | 30 (40%) | 22 (29%) | 0.032 |
| 60 | 40 (53%) | 30 (40%) | 0.35 | 29 (39%) | 16 (21%) | 0.023 |
| 61 | 24 (32%) | 17 (23%) | 0.051 | 28 (37%) | 23 (31%) | 0.0066 |
| 62 | 27 (36%) | 18 (24%) | 0.17 | 18 (24%) | 10 (13%) | -0.045 |
| 63 | 25 (33%) | 16 (21%) | 0.09 | 19 (25%) | 12 (16%) | -0.022 |
| 64 | 36 (48%) | 28 (37%) | 0.26 | 25 (33%) | 21 (28%) | 0.032 |
| 65 | 33 (44%) | 26 (35%) | 0.21 | 24 (32%) | 13 (17%) | 0.0041 |
| 66 | 36 (48%) | 28 (37%) | 0.26 | 25 (33%) | 21 (28%) | 0.032 |
| 67 | 36 (48%) | 28 (37%) | 0.26 | 25 (33%) | 21 (28%) | 0.032 |
| 68 | 30 (40%) | 20 (27%) | 0.17 | 20 (27%) | 10 (13%) | -0.022 |
| 69 | 35 (47%) | 26 (35%) | 0.30 | 28 (37%) | 15 (20%) | 0.039 |
| 70 | 38 (51%) | 31 (41%) | 0.20 | 31 (41%) | 23 (31%) | 0.047 |
| 71 | 36 (48%) | 28 (37%) | 0.26 | 25 (33%) | 21 (28%) | 0.032 |
| 72 | 45 (60%) | 31 (41%) | 0.34 | 25 (33%) | 20 (27%) | -0.0047 |

**Supplementary Table S8 (cont.)**

| **Permutation** | **Logistic / binary** | | | **Linear / quantitative** | | |
| --- | --- | --- | --- | --- | --- | --- |
|  | **# nominally significant** (*p* < 0.05) | **# Bonferroni significant**  (*p* < 0.00067) | **Median coefficient (log odds ratio)** | **# nominally significant**  (*p* < 0.05) | **# Bonferroni significant**  (*p* < 0.00067) | **Median coefficient (population std dev)** |
| *True data* | *54 (72%)* | *45 (60%)* | *0.57* | *43 (57%)* | *38 (51%)* | *0.17* |
| 73 | 28 (37%) | 18 (24%) | 0.14 | 22 (29%) | 14 (19%) | -0.022 |
| 74 | 31 (41%) | 22 (29%) | 0.15 | 30 (40%) | 22 (29%) | 0.032 |
| 75 | 24 (32%) | 19 (25%) | 0.12 | 20 (27%) | 15 (20%) | -0.023 |
| 76 | 24 (32%) | 17 (23%) | 0.051 | 28 (37%) | 23 (31%) | 0.0066 |
| 77 | 16 (21%) | 11 (15%) | 0.051 | 15 (20%) | 10 (13%) | -0.027 |
| 78 | 26 (35%) | 17 (23%) | 0.060 | 19 (25%) | 13 (17%) | -0.0034 |
| 79 | 30 (40%) | 18 (24%) | 0.14 | 23 (31%) | 19 (25%) | -0.0047 |
| 80 | 27 (36%) | 20 (27%) | 0.098 | 21 (28%) | 9 (12%) | -0.0034 |
| 81 | 31 (41%) | 24 (32%) | 0.19 | 14 (19%) | 7 (9.3%) | -0.018 |
| 82 | 24 (32%) | 17 (23%) | 0.051 | 28 (37%) | 23 (31%) | 0.0066 |
| 83 | 33 (44%) | 26 (35%) | 0.21 | 24 (32%) | 13 (17%) | 0.0041 |
| 84 | 32 (43%) | 26 (35%) | 0.21 | 24 (32%) | 19 (25%) | -0.0049 |
| 85 | 30 (40%) | 21 (28%) | 0.15 | 20 (27%) | 10 (13%) | -0.028 |
| 86 | 45 (60%) | 31 (41%) | 0.34 | 25 (33%) | 20 (27%) | -0.0047 |
| 87 | 10 (13%) | 5 (6.7%) | 0.019 | 9 (12%) | 3 (4%) | -0.022 |
| 88 | 28 (37%) | 18 (24%) | 0.14 | 22 (29%) | 14 (19%) | -0.022 |
| 89 | 45 (60%) | 37 (49%) | 0.60 | 33 (44%) | 18 (24%) | 0.041 |
| 90 | 28 (37%) | 23 (31%) | 0.044 | 25 (33%) | 21 (28%) | -0.0034 |
| 91 | 28 (37%) | 21 (28%) | 0.12 | 23 (31%) | 18 (24%) | -0.0043 |
| 92 | 38 (51%) | 26 (35%) | 0.31 | 24 (32%) | 19 (25%) | -9.40E-04 |
| 93 | 20 (27%) | 18 (24%) | 0.035 | 16 (21%) | 9 (12%) | -0.0034 |
| 94 | 24 (32%) | 19 (25%) | 0.12 | 20 (27%) | 15 (20%) | -0.023 |
| 95 | 45 (60%) | 31 (41%) | 0.34 | 25 (33%) | 20 (27%) | -0.0047 |
| 96 | 39 (52%) | 33 (44%) | 0.56 | 31 (41%) | 19 (25%) | 0.020 |
| 97 | 44 (59%) | 34 (45%) | 0.22 | 34 (45%) | 20 (27%) | 0.058 |
| 98 | 28 (37%) | 18 (24%) | 0.17 | 17 (23%) | 10 (13%) | -0.042 |
| 99 | 34 (45%) | 26 (35%) | 0.15 | 35 (47%) | 24 (32%) | 0.069 |
| 100 | 27 (36%) | 23 (31%) | 0.063 | 18 (24%) | 11 (15%) | -0.014 |

**Supplementary Table S9. Regression results for mortality, DALY, and related measures in controls only.** DALY = Disability Adjusted Life Years, YLD = Years Lost to Disability, YLL = Years of Life Lost. DALY, YLD, and YLL are tested by linear regression; mortality is tested by Cox proportional hazard regression. Coefficients reported are the effect of RarePT prediction as a binary variable on the listed outcome, controlling for age, sex, and self-reported ethnicity. “NaN” (not a number) indicates values that could not be calculated due to insufficient sample size in predicted cases; p=0 indicates a p-value below the numerical precision of Python’s statsmodels package (approximately 2 × 10^-308^).

| **Phecode** | **Coefficient (Std. Err.); p-value** | | | |
| --- | --- | --- | --- | --- |
|  | **DALY** | **YLD** | **YLL** | **Mortality** |
| 008.7 | 0.96 (0.077); p=1e-35 | 0.17 (0.0058); p=1e-35 | 0.80 (0.074); p=6.2e-27 | -0.11 (0.11); p=0.35 |
| 070.1 | 3.1 (0.074); p=0 | 0.17 (0.0056); p=0 | 2.9 (0.071); p=0 | 1.0 (0.055); p=9.8e-77 |
| 071 | 0.28 (0.062); p=5.9e-06 | 0.044 (0.0046); p=5.9e-06 | 0.24 (0.059); p=7.1e-05 | 0.37 (0.075); p=7.2e-07 |
| 071.1 | 0.24 (0.10); p=0.019 | 0.047 (0.0077); p=0.019 | 0.19 (0.099); p=0.05 | 0.58 (0.11); p=1.3e-07 |
| 079.1 | 1.8 (0.066); p=1.1e-155 | 0.16 (0.0049); p=1.1e-155 | 1.6 (0.063); p=3.5e-139 | 1.6 (0.042); p=0 |
| 079.2 | 1.7 (0.062); p=1.2e-174 | 0.15 (0.0046); p=1.2e-174 | 1.6 (0.059); p=2.5e-158 | 1.4 (0.041); p=1.6e-269 |
| 110.11 | 0.91 (0.088); p=2.4e-25 | 0.15 (0.0066); p=2.4e-25 | 0.76 (0.084); p=2e-19 | 0.71 (0.077); p=3.2e-20 |
| 110.13 | 1.8 (0.058); p=5e-203 | 0.34 (0.0044); p=5e-203 | 1.4 (0.056); p=3.9e-145 | 1.5 (0.035); p=0 |
| 110.2 | 2.1 (0.048); p=0 | 0.35 (0.0036); p=0 | 1.7 (0.046); p=0 | 1.5 (0.031); p=0 |
| 112.3 | 3.3 (0.033); p=0 | 0.38 (0.0024); p=0 | 2.9 (0.031); p=0 | 1.7 (0.020); p=0 |
| 130 | 0.30 (0.30); p=0.32 | 0.025 (0.022); p=0.32 | 0.27 (0.29); p=0.35 | 0.74 (0.26); p=0.0042 |
| 145.1 | 0.37 (0.061); p=9.6e-10 | -0.024 (0.0046); p=9.6e-10 | 0.40 (0.058); p=1.2e-11 | -1.1 (0.11); p=4.4e-23 |
| 149 | 4.1 (0.036); p=0 | 0.18 (0.0027); p=0 | 3.9 (0.034); p=0 | 7.5e+10 (4800000); p=0 |
| 149.3 | 3.5 (0.058); p=0 | 0.16 (0.0044); p=0 | 3.4 (0.056); p=0 | 1.8 (0.031); p=0 |
| 149.9 | 3.0 (0.051); p=0 | 0.12 (0.0039); p=0 | 2.9 (0.049); p=0 | 1.3 (0.033); p=0 |
| 164 | 1.9 (0.050); p=0 | 0.13 (0.0041); p=0 | 1.8 (0.048); p=0 | NaN (NaN); p=NaN |
| 174.3 | -0.056 (0.14); p=0.69 | -0.034 (0.011); p=0.69 | -0.022 (0.14); p=0.87 | -0.098 (0.21); p=0.64 |
| 187.1 | -2.1 (0.047); p=0 | -0.13 (0.0034); p=0 | -2.0 (0.046); p=0 | 0.64 (0.064); p=1.8e-23 |
| 187.8 | -2.1 (0.047); p=0 | -0.13 (0.0034); p=0 | -2.0 (0.046); p=0 | -0.28 (0.081); p=0.00058 |
| 189.12 | 2.2 (0.065); p=5.8e-252 | 0.14 (0.0049); p=5.8e-252 | 2.1 (0.063); p=2.3e-239 | 1.4 (0.038); p=3.9e-293 |
| 191.1 | 3.0 (0.044); p=0 | 0.16 (0.0034); p=0 | 2.9 (0.042); p=0 | 1.8 (0.025); p=0 |
| 194 | 2.8 (0.11); p=2.9e-151 | 0.12 (0.0081); p=2.9e-151 | 2.7 (0.10); p=5.1e-150 | 2.0 (0.053); p=0 |
| 199.4 | 0.83 (0.1); p=7e-16 | 0.056 (0.011); p=7e-16 | 0.78 (0.098); p=1.5e-15 | 0.93 (0.20); p=2.4e-06 |
| 204.11 | 2.2 (0.04); p=0 | 0.082 (0.003); p=0 | 2.1 (0.038); p=0 | 1.7 (0.025); p=0 |
| 204.3 | 2.3 (0.071); p=5.7e-230 | 0.14 (0.0054); p=5.7e-230 | 2.2 (0.068); p=9e-219 | 1.6 (0.041); p=0 |

**Supplementary Table S9 (cont.)**

| **Phecode** | **Coefficient (Std. Err.); p-value** | | | |
| --- | --- | --- | --- | --- |
|  | **DALY** | **YLD** | **YLL** | **Mortality** |
| 224 | -0.14 (0.15); p=0.34 | 0.020 (0.011); p=0.34 | -0.16 (0.14); p=0.26 | -0.53 (0.22); p=0.017 |
| 224.1 | -0.37 (0.11); p=0.00041 | 0.051 (0.008); p=0.00041 | -0.42 (0.10); p=2.8e-05 | -1.5 (0.24); p=2.9e-10 |
| 225.2 | 0.18 (0.061); p=0.0029 | 0.099 (0.0046); p=0.0029 | 0.083 (0.059); p=0.16 | 0.1 (0.077); p=0.18 |
| 242.2 | 1.4 (0.14); p=5.9e-23 | 0.26 (0.011); p=5.9e-23 | 1.2 (0.14); p=6.5e-17 | 1.1 (0.11); p=7e-23 |
| 242.3 | -0.58 (0.074); p=3.3e-15 | -0.044 (0.0055); p=3.3e-15 | -0.54 (0.071); p=3.3e-14 | -1.1 (0.16); p=1e-12 |
| 246.7 | 1.7 (0.036); p=0 | 0.17 (0.0027); p=0 | 1.5 (0.034); p=0 | 1.2 (0.026); p=0 |
| 250.12 | 1.2 (0.12); p=2e-23 | 0.16 (0.0089); p=2e-23 | 1.0 (0.12); p=2.4e-19 | 1.6 (0.075); p=1.4e-105 |
| 250.3 | 2.2 (0.04); p=0 | 0.22 (0.003); p=0 | 2.0 (0.039); p=0 | 1.1 (0.032); p=1.7e-243 |
| 250.5 | 0.20 (0.044); p=6.6e-06 | 0.021 (0.0033); p=6.6e-06 | 0.18 (0.042); p=2.7e-05 | 0.13 (0.10); p=0.19 |
| 253 | 1.8 (0.046); p=0 | 0.25 (0.0034); p=0 | 1.6 (0.044); p=1.5e-273 | 1.0 (0.034); p=3.7e-200 |
| 253.11 | 1.1 (0.094); p=1.0e-30 | 0.18 (0.0070); p=1.0e-30 | 0.90 (0.090); p=1.0e-23 | 0.30 (0.10); p=0.0028 |
| 253.3 | 2.1 (0.047); p=0 | 0.29 (0.0035); p=0 | 1.8 (0.046); p=0 | 1.3 (0.032); p=0 |
| 255.11 | 2.6 (0.059); p=0 | 0.29 (0.0044); p=0 | 2.4 (0.056); p=0 | 1.3 (0.042); p=1.5e-198 |
| 255.12 | 0.76 (0.058); p=9.8e-40 | 0.13 (0.0043); p=9.8e-40 | 0.64 (0.056); p=2.0e-30 | 0.43 (0.056); p=6.9e-15 |
| 258 | 3.7 (0.078); p=0 | 0.25 (0.0058); p=0 | 3.5 (0.075); p=0 | 1.5 (0.049); p=1.8e-195 |
| 261.41 | 2.2 (0.070); p=3.6e-212 | 0.14 (0.0052); p=3.6e-212 | 2.0 (0.067); p=2.3e-200 | 1.5 (0.046); p=4.0e-230 |
| 270.35 | 2.9 (0.080); p=8.1e-291 | 0.23 (0.0060); p=8.1e-291 | 2.7 (0.077); p=7.4e-267 | 1.5 (0.047); p=1.6e-234 |
| 272.12 | 2.0 (0.069); p=1.4e-177 | 0.26 (0.0049); p=1.4e-177 | 1.7 (0.066); p=1.3e-144 | 0.73 (0.057); p=2.4e-38 |
| 277.51 | 1.7 (0.071); p=4.3e-122 | 0.25 (0.0053); p=4.3e-122 | 1.4 (0.068); p=6.3e-96 | 1.4 (0.044); p=8.1e-229 |
| 278.3 | 0.22 (0.12); p=0.070 | 0.099 (0.0088); p=0.07 | 0.12 (0.12); p=0.30 | -0.60 (0.24); p=0.013 |
| 278.4 | 2.7 (0.06); p=0 | 0.31 (0.0043); p=0 | 2.4 (0.058); p=0 | 1.3 (0.038); p=2.3e-271 |
| 281.9 | 2.4 (0.036); p=0 | 0.33 (0.0027); p=0 | 2.0 (0.035); p=0 | 1.5 (0.025); p=0 |
| 286.11 | 1.1 (0.11); p=1.3e-24 | 0.15 (0.0082); p=1.3e-24 | 0.96 (0.10); p=2.8e-20 | 0.40 (0.12); p=0.0012 |
| 286.13 | 0.99 (0.071); p=9.8e-45 | 0.16 (0.0053); p=9.8e-45 | 0.83 (0.068); p=1.1e-34 | 0.57 (0.071); p=10e-15 |
| 286.5 | 2.4 (0.044); p=0 | 0.27 (0.0033); p=0 | 2.1 (0.043); p=0 | 1.3 (0.028); p=0 |
| 288.3 | 2.0 (0.056); p=7.8e-269 | 0.22 (0.0044); p=7.8e-269 | 1.7 (0.054); p=4.3e-230 | 1.6 (0.035); p=0 |
| 289.9 | 2.9 (0.034); p=0 | 0.31 (0.0026); p=0 | 2.6 (0.032); p=0 | 1.6 (0.022); p=0 |
| 290.3 | 1.5 (0.039); p=0 | 0.33 (0.0028); p=0 | 1.1 (0.037); p=5.7e-206 | 1.5 (0.031); p=0 |
| 297 | -0.26 (0.045); p=1.2e-08 | 0.0067 (0.0030); p=1.2e-08 | -0.26 (0.043); p=1.3e-09 | 0.047 (0.081); p=0.56 |

**Supplementary Table S9 (cont.)**

| **Phecode** | **Coefficient (Std. Err.); p-value** | | | |
| --- | --- | --- | --- | --- |
|  | **DALY** | **YLD** | **YLL** | **Mortality** |
| 300.4 | 0.70 (0.048); p=8.0e-49 | 0.18 (0.0032); p=8.0e-49 | 0.52 (0.046); p=6.3e-30 | 0.70 (0.062); p=1.3e-29 |
| 305.2 | 1.4 (0.057); p=1.3e-122 | 0.19 (0.0038); p=1.3e-122 | 1.2 (0.055); p=3.7e-98 | 1.4 (0.056); p=3.4e-133 |
| 315.3 | 1.5 (0.096); p=1.3e-54 | 0.49 (0.0071); p=1.3e-54 | 1.0 (0.092); p=1.6e-27 | 1.6 (0.056); p=2.8e-179 |
| 323.2 | 0.42 (0.14); p=0.0035 | 0.15 (0.011); p=0.0035 | 0.27 (0.14); p=0.051 | 0.72 (0.16); p=5.9e-06 |
| 325 | 2.6 (0.093); p=1.5e-173 | 0.20 (0.0070); p=1.5e-173 | 2.4 (0.089); p=1.4e-160 | 1.8 (0.055); p=2.4e-242 |
| 337.1 | 1.5 (0.038); p=0 | 0.24 (0.0027); p=0 | 1.2 (0.037); p=6.1e-246 | 0.83 (0.031); p=4.1e-157 |
| 353.1 | 0.31 (0.12); p=0.011 | 0.12 (0.0090); p=0.011 | 0.20 (0.12); p=0.095 | -0.088 (0.17); p=0.60 |
| 353.2 | -0.16 (0.072); p=0.025 | 0.11 (0.0053); p=0.025 | -0.27 (0.070); p=0.00012 | -0.73 (0.13); p=2.7e-08 |
| 361.2 | -0.69 (0.034); p=2.2e-90 | -0.012 (0.0025); p=2.2e-90 | -0.67 (0.033); p=3.1e-94 | -1.2 (0.069); p=7.4e-72 |
| 362.3 | -0.36 (0.05); p=4.5e-13 | 0.017 (0.0036); p=4.5e-13 | -0.38 (0.048); p=2.9e-15 | -0.57 (0.073); p=5.3e-15 |
| 364.41 | -0.74 (0.036); p=1.2e-95 | -0.027 (0.0026); p=1.2e-95 | -0.71 (0.034); p=4.5e-96 | -1.9 (0.10); p=5.4e-77 |
| 366.1 | 0.77 (0.19); p=4.1e-05 | 0.051 (0.014); p=4.1e-05 | 0.72 (0.18); p=6.9e-05 | 0.51 (0.18); p=0.0052 |
| 366.3 | -0.41 (0.085); p=1.9e-06 | -0.011 (0.0062); p=1.9e-06 | -0.40 (0.082); p=1.4e-06 | -0.61 (0.14); p=1.7e-05 |
| 367.4 | -0.79 (0.048); p=1.8e-60 | 0.024 (0.0036); p=1.8e-60 | -0.81 (0.046); p=2.3e-69 | -1.9 (0.13); p=2.1e-46 |
| 377.1 | 0.73 (0.075); p=1.1e-22 | 0.2 (0.0055); p=1.1e-22 | 0.54 (0.072); p=8.1e-14 | 0.54 (0.066); p=1.7e-16 |
| 378 | 0.67 (0.10); p=1.2e-10 | 0.14 (0.0077); p=1.2e-10 | 0.52 (0.10); p=1.5e-07 | 0.50 (0.093); p=5.1e-08 |
| 388 | -0.56 (0.11); p=1.2e-06 | -0.046 (0.0086); p=1.2e-06 | -0.51 (0.11); p=3.5e-06 | -1.1 (0.27); p=2.3e-05 |
| 394.4 | 1.3 (0.055); p=6.3e-127 | 0.24 (0.0040); p=6.3e-127 | 1.1 (0.053); p=7.0e-93 | 0.95 (0.043); p=2.8e-107 |
| 401.2 | 2.4 (0.051); p=0 | 0.28 (0.0036); p=0 | 2.1 (0.050); p=0 | 5.6e+10 (890000); p=0 |
| 420.22 | 1.2 (0.045); p=8.3e-149 | 0.16 (0.0034); p=8.3e-149 | 1.0 (0.044); p=3.6e-121 | 1.1 (0.031); p=2.7e-270 |
| 425.2 | 0.81 (0.083); p=1.6e-22 | 0.11 (0.0063); p=1.6e-22 | 0.71 (0.080); p=1.1e-18 | 1.0 (0.057); p=2.5e-72 |
| 425.8 | 0.78 (0.065); p=2.8e-33 | 0.18 (0.0049); p=2.8e-33 | 0.60 (0.062); p=6.4e-22 | 1.3 (0.040); p=1.6e-233 |
| 426.8 | 1.3 (0.031); p=0 | 0.19 (0.0023); p=0 | 1.1 (0.03); p=3.2e-292 | 1.1 (0.027); p=0 |
| 441.2 | 3.3 (0.040); p=0 | 0.31 (0.0032); p=0 | 3.0 (0.038); p=0 | 1.2 (0.031); p=0 |
| 446 | 0.86 (0.035); p=9.3e-132 | 0.13 (0.0028); p=9.3e-132 | 0.72 (0.034); p=1.3e-101 | 0.81 (0.033); p=3.8e-132 |
| 459.1 | 3.1 (0.11); p=1.7e-188 | 0.26 (0.0078); p=1.7e-188 | 2.8 (0.1.0); p=5.1e-170 | 1.8 (0.067); p=2.0e-155 |
| 473.1 | 0.32 (0.091); p=0.00046 | 0.11 (0.0068); p=0.00046 | 0.21 (0.087); p=0.016 | 0.19 (0.096); p=0.050 |
| 500.1 | 1.5 (0.045); p=1.7e-241 | 0.20 (0.0035); p=1.7e-241 | 1.3 (0.043); p=2.4e-197 | 1.1 (0.042); p=3.8e-149 |
| 519 | 2.1 (0.050); p=0 | 0.18 (0.0037); p=0 | 2.0 (0.048); p=0 | 1.1 (0.039); p=1.6e-176 |

**Supplementary Table S9 (cont.)**

| **Phecode** | **Coefficient (Std. Err.); p-value** | | | |
| --- | --- | --- | --- | --- |
|  | **DALY** | **YLD** | **YLL** | **Mortality** |
| 523.1 | 0.68 (0.15); p=5.1e-06 | 0.049 (0.011); p=5.1e-06 | 0.63 (0.14); p=1.1e-05 | 1.2 (0.12); p=5.7e-24 |
| 527.1 | 0.91 (0.24); p=0.00016 | 0.076 (0.018); p=0.00016 | 0.83 (0.23); p=0.00032 | 0.51 (0.25); p=0.043 |
| 527.7 | 1.5 (0.074); p=8.2e-86 | 0.17 (0.0056); p=8.2e-86 | 1.3 (0.071); p=1.3e-72 | 1.6 (0.044); p=1.0e-291 |
| 528.12 | 2.6 (0.09); p=1.3e-180 | 0.25 (0.0068); p=1.3e-180 | 2.3 (0.087); p=1.4e-159 | 1.5 (0.057); p=1.6e-152 |
| 528.41 | 0.29 (0.17); p=0.085 | 0.040 (0.013); p=0.085 | 0.25 (0.16); p=0.12 | 0.39 (0.17); p=0.027 |
| 573.2 | 4.6 (0.098); p=0 | 0.30 (0.0079); p=0 | 4.3 (0.094); p=0 | 1.9 (0.062); p=1e-200 |
| 580.11 | 1.7 (0.053); p=3.2e-215 | 0.20 (0.0040); p=3.2e-215 | 1.5 (0.051); p=1.7e-180 | 1.5 (0.041); p=1.1e-278 |
| 588 | 2.4 (0.046); p=0 | 0.29 (0.0034); p=0 | 2.2 (0.045); p=0 | 1.8 (0.031); p=0 |
| 597.2 | 2.1 (0.090); p=2.4e-115 | 0.20 (0.0067); p=2.4e-115 | 1.9 (0.087); p=4.0e-102 | 1.1 (0.066); p=1.3e-58 |
| 609.2 | -2.1 (0.049); p=0 | -0.12 (0.0034); p=0 | -1.9 (0.048); p=0 | -0.44 (0.12); p=0.00029 |
| 611.1 | 0.56 (0.076); p=1.4e-13 | 0.046 (0.0057); p=1.4e-13 | 0.52 (0.073); p=1.6e-12 | 0.8 (0.076); p=7.9e-26 |
| 612.3 | 0.063 (0.068); p=0.35 | 0.075 (0.0051); p=0.35 | -0.013 (0.065); p=0.85 | -0.89 (0.17); p=1.3e-07 |
| 613.5 | 1.0 (0.045); p=1.8e-116 | 0.23 (0.0034); p=1.8e-116 | 0.81 (0.044); p=7.1e-77 | 0.53 (0.049); p=6.9e-27 |
| 620 | -0.29 (0.054); p=7.9e-08 | -0.027 (0.0043); p=7.9e-08 | -0.27 (0.052); p=3.7e-07 | NaN (NaN); p=NaN |
| 626.11 | -0.33 (0.066); p=7.7e-07 | -0.032 (0.0051); p=7.7e-07 | -0.29 (0.063); p=3.6e-06 | NaN (NaN); p=NaN |
| 626.4 | -0.48 (0.047); p=1.8e-24 | -0.034 (0.0036); p=1.8e-24 | -0.45 (0.045); p=5.6e-23 | NaN (NaN); p=NaN |
| 643.1 | -0.20 (0.025); p=3.4e-15 | -0.047 (0.0020); p=3.4e-15 | -0.15 (0.024); p=3.2e-10 | NaN (NaN); p=NaN |
| 647 | -0.20 (0.031); p=7.9e-11 | -0.047 (0.0024); p=7.9e-11 | -0.15 (0.029); p=2.1e-07 | NaN (NaN); p=NaN |
| 649 | -0.17 (0.028); p=2.2e-09 | -0.041 (0.0022); p=2.2e-09 | -0.13 (0.027); p=2.4e-06 | NaN (NaN); p=NaN |
| 656.2 | 2.1 (0.071); p=4.9e-185 | 0.29 (0.0053); p=4.9e-185 | 1.8 (0.068); p=7.9e-148 | 8.0e+10 (9800000); p=0 |
| 686.5 | 2.5 (0.16); p=5.0e-57 | 0.26 (0.012); p=5.0e-57 | 2.3 (0.15); p=1.0e-49 | 1.6 (0.11); p=3.0e-49 |
| 690.1 | 1.5 (0.065); p=8.2e-117 | 0.24 (0.0048); p=8.2e-117 | 1.3 (0.063); p=4.1e-90 | 0.86 (0.052); p=2.5e-60 |
| 691 | 2.8 (0.13); p=2.1e-99 | 0.28 (0.0098); p=2.1e-99 | 2.5 (0.13); p=2.5e-87 | 1.5 (0.088); p=2.4e-62 |
| 694.1 | 1.6 (0.070); p=2.0e-113 | 0.25 (0.0052); p=2.0e-113 | 1.3 (0.067); p=9.3e-87 | 0.65 (0.066); p=3.9e-23 |
| 695.21 | 0.50 (0.068); p=1.1e-13 | 0.081 (0.0050); p=1.1e-13 | 0.42 (0.065); p=9.3e-11 | 0.50 (0.073); p=7.7e-12 |
| 695.81 | 1.2 (0.081); p=9.7e-52 | 0.15 (0.006); p=9.7e-52 | 1.1 (0.077); p=5.2e-43 | 1.0 (0.074); p=3.9e-44 |
| 696.3 | -0.0076 (0.094); p=0.94 | 0.023 (0.0070); p=0.94 | -0.030 (0.090); p=0.74 | -0.59 (0.16); p=0.00019 |
| 701.1 | 0.68 (0.070); p=4.7e-22 | 0.10 (0.0052); p=4.7e-22 | 0.57 (0.067); p=1.2e-17 | 0.23 (0.071); p=0.0015 |
| 701.3 | 0.56 (0.12); p=1.6e-06 | 0.088 (0.0088); p=1.6e-06 | 0.47 (0.11); p=2.5e-05 | 0.54 (0.11); p=5.5e-07 |

**Supplementary Table S9 (cont.)**

| **Phecode** | **Coefficient (Std. Err.); p-value** | | | |
| --- | --- | --- | --- | --- |
|  | **DALY** | **YLD** | **YLL** | **Mortality** |
| 701.6 | 0.037 (0.11); p=0.73 | -0.018 (0.0081); p=0.73 | 0.055 (0.10); p=0.60 | -1.3 (0.24); p=3.9e-08 |
| 704.8 | -0.014 (0.13); p=0.91 | -3.4e-05 (0.0094); p=0.91 | -0.014 (0.12); p=0.91 | -1.3 (0.27); p=2.6e-06 |
| 706.1 | 0.052 (0.13); p=0.69 | 0.077 (0.0097); p=0.69 | -0.025 (0.12); p=0.84 | -0.38 (0.19); p=0.047 |
| 709.5 | 0.13 (0.088); p=0.13 | 0.055 (0.0065); p=0.13 | 0.078 (0.085); p=0.36 | 0.13 (0.11); p=0.20 |
| 709.6 | 0.27 (0.086); p=0.0018 | 0.067 (0.0064); p=0.0018 | 0.20 (0.083); p=0.015 | 0.35 (0.098); p=0.00039 |
| 712 | 1.5 (0.080); p=6.1e-74 | 0.18 (0.0054); p=6.1e-74 | 1.3 (0.077); p=3.8e-61 | 1.4 (0.054); p=1.7e-152 |
| 713.5 | 1.5 (0.077); p=1.4e-88 | 0.28 (0.0052); p=1.4e-88 | 1.3 (0.074); p=3.2e-64 | 1.6 (0.045); p=2.0e-284 |
| 722.3 | 0.75 (0.047); p=1.5e-58 | 0.18 (0.0034); p=1.5e-58 | 0.57 (0.045); p=1.1e-36 | 0.56 (0.046); p=1.0e-33 |
| 722.8 | -0.29 (0.059); p=1.4e-06 | 0.087 (0.0043); p=1.4e-06 | -0.37 (0.057); p=5.8e-11 | -0.76 (0.12); p=5.5e-10 |
| 724.8 | 0.47 (0.066); p=4.9e-13 | 0.13 (0.0047); p=4.9e-13 | 0.35 (0.063); p=4.5e-08 | 0.36 (0.072); p=6.6e-07 |
| 726.4 | -0.40 (0.050); p=2.0e-15 | 0.044 (0.0036); p=2.0e-15 | -0.44 (0.048); p=4.9e-20 | -1.2 (0.12); p=5.1e-24 |
| 727.2 | -0.61 (0.069); p=1.2e-18 | -0.033 (0.0050); p=1.2e-18 | -0.58 (0.066); p=4.3e-18 | -1.0 (0.15); p=9.1e-12 |
| 737 | 1.7 (0.037); p=0 | 0.34 (0.0027); p=0 | 1.4 (0.036); p=0 | 0.88 (0.03); p=3.5e-195 |
| 737.2 | 0.58 (0.035); p=3.3e-60 | 0.21 (0.0026); p=3.3e-60 | 0.36 (0.034); p=6.5e-27 | 0.27 (0.039); p=3.8e-12 |
| 741.1 | 0.50 (0.087); p=7.7e-09 | 0.15 (0.0065); p=7.7e-09 | 0.36 (0.084); p=1.9e-05 | -0.09 (0.11); p=0.43 |
| 742 | -0.65 (0.045); p=1.5e-46 | -0.028 (0.0034); p=1.5e-46 | -0.62 (0.044); p=4e-46 | -0.97 (0.085); p=7.9e-30 |
| 743.4 | 1.4 (0.049); p=1.1e-177 | 0.29 (0.0036); p=1.1e-177 | 1.1 (0.047); p=2.7e-120 | 1.2 (0.032); p=1.3e-291 |
| 748 | 1.9 (0.093); p=4.5e-94 | 0.095 (0.007); p=4.5e-94 | 1.8 (0.090); p=3.5e-92 | 0.89 (0.070); p=3.6e-37 |
| 750.1 | 1.0 (0.052); p=2.5e-88 | 0.20 (0.0039); p=2.5e-88 | 0.84 (0.050); p=1.5e-63 | -0.079 (0.059); p=0.18 |
| 750.15 | 1.9 (0.039); p=0 | 0.27 (0.0029); p=0 | 1.6 (0.037); p=0 | 0.54 (0.033); p=1.8e-60 |
| 751.2 | 0.48 (0.047); p=6.8e-25 | 0.11 (0.0035); p=6.8e-25 | 0.37 (0.045); p=1.2e-16 | -0.033 (0.051); p=0.51 |
| 752.1 | 0.27 (0.10); p=0.0074 | 0.24 (0.0076); p=0.0074 | 0.034 (0.096); p=0.72 | -0.037 (0.14); p=0.79 |
| 753.1 | -0.65 (0.065); p=2.1e-23 | 0.068 (0.0049); p=2.1e-23 | -0.72 (0.063); p=1.8e-30 | -1.6 (0.16); p=3.2e-23 |
| 755.6 | -0.33 (0.13); p=0.012 | 0.025 (0.0099); p=0.012 | -0.35 (0.13); p=0.0051 | -0.87 (0.26); p=0.00071 |
| 755.61 | -0.54 (0.11); p=2.4e-06 | -0.0047 (0.0086); p=2.4e-06 | -0.54 (0.11); p=1.1e-06 | -1.1 (0.28); p=7.8e-05 |
| 756 | 0.13 (0.14); p=0.35 | 0.15 (0.010); p=0.35 | -0.022 (0.13); p=0.87 | -0.43 (0.22); p=0.054 |
| 756.3 | 0.38 (0.070); p=6.4e-08 | 0.23 (0.0053); p=6.4e-08 | 0.15 (0.067); p=0.026 | -0.55 (0.13); p=3.5e-05 |
| 758 | 0.72 (0.067); p=6.1e-27 | 0.10 (0.0050); p=6.1e-27 | 0.61 (0.064); p=7.4e-22 | 0.22 (0.066); p=0.0011 |
| 758.1 | 1.6 (0.11); p=1.2e-46 | 0.21 (0.0083); p=1.2e-46 | 1.4 (0.11); p=3.4e-38 | 1.0 (0.083); p=9.1e-35 |

**Supplementary Table S9 (cont.)**

| **Phecode** | **Coefficient (Std. Err.); p-value** | | | |
| --- | --- | --- | --- | --- |
|  | **DALY** | **YLD** | **YLL** | **Mortality** |
| 759.1 | 0.054 (0.11); p=0.61 | 0.079 (0.0081); p=0.61 | -0.025 (0.10); p=0.81 | -0.46 (0.17); p=0.008 |
| 790.1 | 1.9 (0.063); p=8.9e-201 | 0.28 (0.0047); p=8.9e-201 | 1.6 (0.061); p=1.1e-159 | 1.1 (0.049); p=7.7e-101 |
| 794 | -0.039 (0.034); p=0.26 | -0.039 (0.0026); p=0.26 | 0.00027 (0.033); p=0.99 | -0.9 (0.15); p=8.9e-10 |
| 797 | 4.3 (0.031); p=0 | 0.27 (0.0023); p=0 | 4.1 (0.03); p=0 | NaN (NaN); p=NaN |
| 801.1 | -0.30 (0.053); p=9.7e-09 | 0.087 (0.0039); p=9.7e-09 | -0.39 (0.051); p=1.6e-14 | -0.20 (0.073); p=0.0061 |
| 860 | 2.9 (0.055); p=0 | 0.20 (0.0041); p=0 | 2.6 (0.053); p=0 | NaN (NaN); p=NaN |
| 870.8 | 0.087 (0.13); p=0.52 | 0.062 (0.010); p=0.52 | 0.025 (0.13); p=0.85 | 0.48 (0.16); p=0.0026 |
| 871.4 | 0.57 (0.086); p=5.4e-11 | 0.09 (0.0064); p=5.4e-11 | 0.48 (0.083); p=9.4e-09 | 0.66 (0.074); p=2.6e-19 |
| 913 | -0.32 (0.24); p=0.19 | -0.04 (0.018); p=0.19 | -0.28 (0.23); p=0.23 | -1.5 (0.71); p=0.036 |
| 938 | 1.5 (0.23); p=3.6e-11 | 0.12 (0.017); p=3.6e-11 | 1.4 (0.22); p=2.4e-10 | 1.3 (0.15); p=1.4e-16 |
| 960.1 | 1 (0.042); p=4.0e-137 | 0.23 (0.0030); p=4e-137 | 0.81 (0.040); p=1.1e-89 | 1.2 (0.033); p=5.7e-292 |
| 975 | 1.4 (0.042); p=1.3e-264 | 0.27 (0.0030); p=1.3e-264 | 1.2 (0.040); p=6.9e-189 | 1.3 (0.031); p=0 |
| 987 | 1.2 (0.086); p=3.2e-46 | 0.37 (0.0064); p=3.2e-46 | 0.87 (0.083); p=1.8e-25 | 1.2 (0.067); p=2.4e-74 |
| 989 | 1.7 (0.078); p=1.5e-99 | 0.45 (0.0058); p=1.5e-99 | 1.2 (0.075); p=7.3e-58 | 1.6 (0.051); p=6.5e-213 |

**Supplementary Table S10. Regression results for 75 diagnostic tests in controls.**

| **Phecode** | **Phenotype** | **Test** | **Direction** | **Coefficient (Std. Err.); p-value** | |
| --- | --- | --- | --- | --- | --- |
|  |  |  |  | **Logistic / binary** | **Linear / quantitative** |
| 071.1 | HIV infection, symptomatic | White blood cell count | below | 0.41 (0.22); p=0.068 | -0.089 (0.057); p=0.12 |
|  |  | Haemoglobin concentration | below | 0.45 (0.20); p=0.025 | 0.12 (0.056); p=0.033 |
|  |  | Platelet count | below | 1.1 (0.19); p=7.7e-09 | -0.10 (0.046); p=0.026 |
|  |  | Neutrophill count | below | 0.83 (0.38); p=0.031 | -0.032 (0.046); p=0.49 |
|  |  | Red blood cell count | below | 0.75 (0.18); p=2.9e-05 | 0.045 (0.042); p=0.28 |
| 079.2 | Infectious mononucleosis | Haemoglobin concentration | below | 0.97 (0.099); p=1.0e-22 | 0.17 (0.034); p=5.1e-07 |
|  |  | Red blood cell count | below | 1.3 (0.085); p=6.6e-49 | 0.11 (0.025); p=2.6e-05 |
| 250.12 | Type 1 diabetes with renal manifestations | Serum albumin | below | 3.8 (0.32); p=3.6e-33 | 0.50 (0.059); p=1.3e-17 |
| 253 | Disorders of the pituitary gland | Body mass index | above | 0.28 (0.038); p=3.9e-13 | 0.027 (0.022); p=0.21 |
| 277.51 | Lipoprotein disorders | Total cholesterol | above | -0.88 (0.057); p=5.2e-54 | -0.44 (0.042); p=2e-25 |
| 278.3 | Localized adiposity | Body mass index | above | 0.28 (0.11); p=0.0094 | 0.21 (0.054); p=0.00015 |
| 281.9 | Deficiency anemias | Red blood cell count | below | 0.081 (0.016); p=4.3e-07 | 0.081 (0.016); p=4.3e-07 |
|  |  | Haemoglobin concentration | below | 0.12 (0.021); p=2.2e-08 | 0.12 (0.021); p=2.2e-08 |
| 286.11 | Von Willebrand's disease | Platelet count | below | 0.44 (0.31); p=0.15 | -0.14 (0.048); p=0.0028 |
| 288.3 | Eosinophilia | Eosinophill count | above | 0.73 (0.11); p=3.2e-11 | 0.16 (0.024); p=1.5e-11 |
| 289.9 | Abnormality of red blood cells | Haemoglobin concentration | outside | 0.067 (0.033); p=0.046 | 0.13 (0.0098); p=1.0e-42 |
|  |  | Mean corpuscular volume | outside | 0.90 (0.045); p=2.2e-90 | 0.087 (0.0087); p=1.0e-23 |
|  |  | Red blood cell count | Outside | 1.1 (0.052); p=4.7e-93 | 0.097 (0.0087); p=8.9e-29 |
|  |  | Reticulocyte count | Outside | 0.38 (0.033); p=2.7e-29 | 0.027 (0.0088); p=0.0018 |

**Supplementary Table S10 (cont).**

| **Phecode** | **Phenotype** | **Test** | **Direction** | **Coefficient (Std. Err.); p-value** | |
| --- | --- | --- | --- | --- | --- |
|  |  |  |  | **Logistic / binary** | **Linear / quantitative** |
| 289.9 | Abnormality of red blood cells | Mean corpuscular haemoglobin | outside | 0.20 (0.029); p=2.2e-12 | 0.067 (0.010); p=1.3e-10 |
|  |  | Haematocrit percentage | outside | 0.3 (0.031); p=5.6e-22 | 0.078 (0.0095); p=2.4e-16 |
| 315.3 | Mental retardation | Haemoglobin concentration | below | 1.2 (0.14); p=8.2e-19 | 0.29 (0.055); p=1.3e-07 |
|  |  | Body mass index | above | 0.63 (0.075); p=4.7e-17 | -0.078 (0.048); p=0.10 |
|  |  | Speech reception threshold (right ear) | above | 0.46 (0.13); p=0.00044 | 0.18 (0.074); p=0.013 |
|  |  | Speech reception threshold (left ear) | above | 0.57 (0.13); p=1.0e-05 | -0.031 (0.075); p=0.68 |
|  |  | Heel bone mineral density | below | 0.42 (0.11); p=8.1e-05 | -0.083 (0.12); p=0.50 |
|  |  | Red blood cell count | below | 1.2 (0.13); p=1.0e-18 | 0.14 (0.040); p=0.00079 |
|  |  | Neutrophill count | below | -1.3 (1.0); p=0.21 | -0.41 (0.043); p=5.2e-21 |
| 366.1 | Nonsenile Cataract | Standing height | below | 0.12 (1.0); p=0.90 | -0.064 (0.077); p=0.41 |
| 377.1 | Optic atrophy | Speech reception threshold (right ear) | above | -0.010 (0.11); p=0.92 | 0.13 (0.054); p=0.014 |
|  |  | Speech reception threshold (left ear) | above | 0.16 (0.10); p=0.12 | 0.0091 (0.055); p=0.87 |
| 459.1 | Hemorrhage NOS | Platelet count | below | 1.3 (0.20); p=2.0e-10 | -0.20 (0.054); p=0.00018 |
| 519 | Other diseases of respiratory system | Red blood cell count | above | 0.66 (0.28); p=0.019 | -0.009 (0.022); p=0.68 |
| 588 | Disorders resulting from impaired renal function | Serum creatinine | outside | -0.035 (0.055); p=0.53 | 0.13 (0.013); p=4.9e-23 |
|  |  | Blood urea | above | 0.4 (0.084); p=1.8e-06 | -0.2 (0.021); p=1.3e-21 |
|  |  | Haemoglobin concentration | below | 0.85 (0.091); p=8.0e-21 | 0.12 (0.028); p=2.7e-05 |
|  |  | Serum urate | above | 0.084 (0.050); p=0.091 | -0.097 (0.024); p=6.5e-05 |
|  |  | Red blood cell count | below | 0.92 (0.083); p=7.8e-29 | 0.059 (0.021); p=0.0048 |

**Supplementary Table S10 (cont).**

| **Phecode** | **Phenotype** | **Test** | **Direction** | **Coefficient (Std. Err.); p-value** | |
| --- | --- | --- | --- | --- | --- |
|  |  |  |  | **Logistic / binary** | **Linear / quantitative** |
| 649 | Other complications of pregnancy | Body mass index | above | *Failed to converge* | -0.12 (0.012); p=3e-22 |
| 686.5 | Pyoderma | Neutrophill count | above | 1.4 (0.26); p=1.3e-07 | 0.086 (0.065); p=0.19 |
| 691 | Congenital anomalies of skin | Red blood cell count | below | 0.73 (0.21); p=0.00043 | 0.0087 (0.054); p=0.87 |
|  |  | Neutrophill count | below | 0.084 (0.71); p=0.91 | -0.32 (0.059); p=6.5e-08 |
|  |  | Platelet count | below | 1.2 (0.24); p=1.2e-06 | -0.093 (0.059); p=0.12 |
|  |  | Standing height | below | 0.63 (0.51); p=0.22 | 0.19 (0.054); p=0.00056 |
|  |  | Lymphocyte count | below | 0.77 (0.23); p=0.00079 | -0.38 (0.12); p=0.0015 |
|  |  | Body mass index | above | 0.62 (0.10); p=1.2e-09 | -0.012 (0.065); p=0.85 |
|  |  | Haemoglobin concentration | below | 0.62 (0.22); p=0.0041 | 0.13 (0.073); p=0.080 |
|  |  | Heel bone mineral density | outside | 0.43 (0.14); p=0.0016 | -0.00029 (0.052); p=1.0 |
|  |  | White blood cell count | below | 0.21 (0.32); p=0.52 | -0.42 (0.074); p=1.5e-08 |
| 704.8 | Other specified diseases of hair and follicles | Neutrophill count | below | 0.79 (0.50); p=0.12 | -0.023 (0.055); p=0.68 |
| 706.1 | Acne | Neutrophil count | above | 0.14 (0.36); p=0.69 | 0.044 (0.050); p=0.38 |
| 709.6 | Other specified diffuse diseases of connective tissue | Eosinophil count | above | 0.23 (0.21); p=0.26 | -0.043 (0.034); p=0.21 |
|  |  | Platelet count | below | 0.27 (0.26); p=0.30 | -0.0053 (0.038); p=0.89 |
| 737 | Curvature of spine | Standing height | below | 0.71 (0.13); p=6.9e-08 | 0.14 (0.015); p=4.1e-20 |
| 755.61 | Congenital hip dysplasia and deformity | Standing height | below | 0.31 (0.51); p=0.55 | -0.12 (0.047); p=0.012 |
| 756 | Other congenital musculoskeletal anomalies | Standing height | outside | 0.17 (0.23); p=0.45 | 0.015 (0.031); p=0.64 |
|  |  | Heel bone mineral density | above | 0.15 (0.21); p=0.47 | 0.13 (0.12); p=0.25 |

**Supplementary Table S10 (cont).**

| **Phecode** | **Phenotype** | **Test** | **Direction** | **Coefficient (Std. Err.); p-value** | |
| --- | --- | --- | --- | --- | --- |
|  |  |  |  | **Logistic / binary** | **Linear / quantitative** |
| 756.3 | Congenital anomalies of muscle and connective tissue | Standing height | below | 0.097 (0.31); p=0.75 | 0.11 (0.028); p=0.00012 |
|  |  | Heel bone mineral density | outside | 0.056 (0.072); p=0.44 | -0.022 (0.025); p=0.38 |
| 758 | Chromosomal anomalies and genetic disorders | Body mass index | above | -0.0023 (0.057); p=0.97 | -0.055 (0.029); p=0.064 |
|  |  | Standing height | below | -0.29 (0.50); p=0.56 | -0.13 (0.027); p=8.1e-07 |
| 758.1 | Chromosomal anomalies | Haemoglobin concentration | below | 1.4 (0.16); p=4.0e-18 | 0.29 (0.061); p=1.6e-06 |
|  |  | Body mass index | above | 0.34 (0.088); p=0.00013 | -0.027 (0.051); p=0.60 |
|  |  | Heel bone mineral density | above | -0.085 (0.17); p=0.62 | -0.34 (0.093); p=0.00025 |
|  |  | White blood cell count | above | 0.75 (0.19); p=6.0e-05 | -0.029 (0.045); p=0.52 |
|  |  | Total cholesterol | above | -0.51 (0.088); p=5.7e-09 | -0.24 (0.072); p=0.00088 |
|  |  | Red blood cell count | below | 1.5 (0.14); p=2.4e-25 | 0.12 (0.046); p=0.0093 |
|  |  | Neutrophil count | below | 0.99 (0.41); p=0.017 | -0.11 (0.049); p=0.021 |
|  |  | Standing height | below | 1.2 (0.42); p=0.0040 | 0.0046 (0.045); p=0.92 |
| 794 | Abnormal results of other function studies | Basal metabolic rate | outside | -0.31 (0.029); p=2.9e-27 | -0.023 (0.0082); p=0.0051 |
| 797 | Shock | Blood glucose | below | 0.046 (0.058); p=0.43 | -0.79 (0.039); p=7.3e-91 |
| 913 | Toxic effect of venom | Haemoglobin concentration | below | 0.071 (0.47); p=0.88 | -0.011 (0.13); p=0.93 |
|  |  | Red blood cell count | below | -0.10 (0.51); p=0.84 | 0.011 (0.095); p=0.91 |
| 989 | Toxic effect of other substances, chiefly nonmedicinal as to source | Haemoglobin concentration | below | 0.63 (0.14); p=5.1e-06 | 0.13 (0.043); p=0.0022 |
|  |  | Red blood cell count | below | 0.98 (0.12); p=8.0e-17 | 0.16 (0.032); p=6.9e-07 |

**Supplementary Table S11. Regression results for 100 permutations of phecode-test relationships in controls.**

| **Permutation** | **Logistic / binary** | | | **Linear / quantitative** | | |
| --- | --- | --- | --- | --- | --- | --- |
|  | **# nominally significant** (*p* < 0.05) | **# Bonferroni significant**  (*p* < 0.00067) | **Median coefficient (log odds ratio)** | **# nominally significant**  (*p* < 0.05) | **# Bonferroni significant**  (*p* < 0.00067) | **Median coefficient (population std dev)** |
| *True data* | *47 (63%)* | *36 (48%)* | *0.45* | *29 (39%)* | *21 (28%)* | *0.0087* |
| 1 | 26 (35%) | 17 (23%) | 0.28 | 8 (11%) | 5 (6.7%) | -0.030 |
| 2 | 31 (41%) | 18 (24%) | 0.21 | 18 (24%) | 12 (16%) | -0.0075 |
| 3 | 17 (23%) | 11 (15%) | 0.12 | 5 (6.7%) | 4 (5.3%) | -0.033 |
| 4 | 18 (24%) | 9 (12%) | 0.066 | 8 (11%) | 4 (5.3%) | -0.041 |
| 5 | 26 (35%) | 17 (23%) | 0.15 | 22 (29%) | 16 (21%) | -0.026 |
| 6 | 48 (64%) | 33 (44%) | 0.63 | 12 (16%) | 5 (6.7%) | -0.068 |
| 7 | 32 (43%) | 21 (28%) | 0.31 | 18 (24%) | 12 (16%) | -0.030 |
| 8 | 32 (43%) | 22 (29%) | 0.14 | 21 (28%) | 11 (15%) | -0.029 |
| 9 | 48 (64%) | 32 (43%) | 0.72 | 12 (16%) | 5 (6.7%) | -0.068 |
| 10 | 27 (36%) | 19 (25%) | 0.20 | 15 (20%) | 10 (13%) | -0.0081 |
| 11 | 25 (33%) | 15 (20%) | 0.26 | 9 (12%) | 6 (8%) | -0.018 |
| 12 | 35 (47%) | 26 (35%) | 0.33 | 16 (21%) | 11 (15%) | -0.037 |
| 13 | 48 (64%) | 32 (43%) | 0.72 | 12 (16%) | 5 (6.7%) | -0.068 |
| 14 | 43 (57%) | 29 (39%) | 0.55 | 17 (23%) | 10 (13%) | -0.0026 |
| 15 | 26 (35%) | 17 (23%) | 0.28 | 8 (11%) | 5 (6.7%) | -0.030 |
| 16 | 32 (43%) | 21 (28%) | 0.31 | 18 (24%) | 12 (16%) | -0.030 |
| 17 | 17 (23%) | 11 (15%) | 0.089 | 9 (12%) | 7 (9.3%) | -0.044 |
| 18 | 38 (51%) | 27 (36%) | 0.58 | 24 (32%) | 14 (19%) | 0.007 |
| 19 | 32 (43%) | 21 (28%) | 0.31 | 18 (24%) | 12 (16%) | -0.030 |
| 20 | 25 (33%) | 15 (20%) | 0.26 | 9 (12%) | 6 (8%) | -0.018 |
| 21 | 36 (48%) | 26 (35%) | 0.22 | 22 (29%) | 11 (15%) | 0.0034 |
| 22 | 31 (41%) | 23 (31%) | 0.49 | 10 (13%) | 8 (11%) | -0.060 |
| 23 | 26 (35%) | 21 (28%) | 0.50 | 10 (13%) | 6 (8%) | -0.036 |
| 24 | 32 (43%) | 22 (29%) | 0.14 | 21 (28%) | 11 (15%) | -0.029 |
| 25 | 42 (56%) | 30 (40%) | 0.56 | 13 (17%) | 7 (9.3%) | -0.045 |
| 26 | 38 (51%) | 24 (32%) | 0.48 | 20 (27%) | 12 (16%) | 0.0097 |
| 27 | 28 (37%) | 18 (24%) | 0.05 | 21 (28%) | 16 (21%) | -0.030 |
| 28 | 26 (35%) | 17 (23%) | 0.28 | 8 (11%) | 5 (6.7%) | -0.030 |
| 29 | 19 (25%) | 12 (16%) | 0.091 | 23 (31%) | 18 (24%) | -0.031 |
| 30 | 40 (53%) | 28 (37%) | 0.27 | 13 (17%) | 6 (8%) | -0.040 |
| 31 | 40 (53%) | 28 (37%) | 0.27 | 13 (17%) | 6 (8%) | -0.040 |
| 32 | 40 (53%) | 28 (37%) | 0.49 | 11 (15%) | 6 (8%) | -0.018 |
| 33 | 31 (41%) | 20 (27%) | 0.35 | 16 (21%) | 13 (17%) | -0.030 |
| 34 | 38 (51%) | 27 (36%) | 0.58 | 24 (32%) | 14 (19%) | 0.0070 |
| 35 | 27 (36%) | 15 (20%) | 0.18 | 16 (21%) | 11 (15%) | -0.011 |

**Supplementary Table S11 (cont.)**

| **Permutation** | **Logistic / binary** | | | **Linear / quantitative** | | |
| --- | --- | --- | --- | --- | --- | --- |
|  | **# nominally significant** (*p* < 0.05) | **# Bonferroni significant**  (*p* < 0.00067) | **Median coefficient (log odds ratio)** | **# nominally significant**  (*p* < 0.05) | **# Bonferroni significant**  (*p* < 0.00067) | **Median coefficient (population std dev)** |
| *True data* | *47 (63%)* | *36 (48%)* | *0.45* | *29 (39%)* | *21 (28%)* | *0.0087* |
| 36 | 40 (53%) | 28 (37%) | 0.27 | 13 (17%) | 6 (8%) | -0.040 |
| 37 | 19 (25%) | 12 (16%) | 0.091 | 23 (31%) | 18 (24%) | -0.031 |
| 38 | 36 (48%) | 22 (29%) | 0.46 | 16 (21%) | 7 (9.3%) | 0.0074 |
| 39 | 48 (64%) | 32 (43%) | 0.72 | 12 (16%) | 5 (6.7%) | -0.068 |
| 40 | 31 (41%) | 22 (29%) | 0.13 | 23 (31%) | 13 (17%) | 0.0034 |
| 41 | 36 (48%) | 22 (29%) | 0.34 | 20 (27%) | 13 (17%) | 0.012 |
| 42 | 26 (35%) | 17 (23%) | 0.28 | 8 (11%) | 5 (6.7%) | -0.030 |
| 43 | 20 (27%) | 13 (17%) | 0.079 | 12 (16%) | 6 (8%) | -0.044 |
| 44 | 36 (48%) | 22 (29%) | 0.46 | 16 (21%) | 7 (9.3%) | 0.0074 |
| 45 | 40 (53%) | 28 (37%) | 0.60 | 23 (31%) | 13 (17%) | 0.0057 |
| 46 | 25 (33%) | 18 (24%) | 0.28 | 8 (11%) | 5 (6.7%) | -0.045 |
| 47 | 48 (64%) | 32 (43%) | 0.72 | 12 (16%) | 5 (6.7%) | -0.068 |
| 48 | 45 (60%) | 33 (44%) | 0.65 | 13 (17%) | 4 (5.3%) | -0.037 |
| 49 | 32 (43%) | 21 (28%) | 0.31 | 19 (25%) | 13 (17%) | -0.030 |
| 50 | 23 (31%) | 18 (24%) | 0.12 | 8 (11%) | 5 (6.7%) | -0.045 |
| 51 | 38 (51%) | 27 (36%) | 0.58 | 24 (32%) | 14 (19%) | 0.007 |
| 52 | 13 (17%) | 8 (11%) | 0.051 | 7 (9.3%) | 5 (6.7%) | -0.014 |
| 53 | 32 (43%) | 21 (28%) | 0.31 | 18 (24%) | 12 (16%) | -0.030 |
| 54 | 32 (43%) | 21 (28%) | 0.31 | 18 (24%) | 12 (16%) | -0.030 |
| 55 | 43 (57%) | 29 (39%) | 0.55 | 17 (23%) | 10 (13%) | -0.0026 |
| 56 | 20 (27%) | 12 (16%) | 0.072 | 12 (16%) | 9 (12%) | -0.013 |
| 57 | 23 (31%) | 15 (20%) | 0.12 | 6 (8%) | 3 (4%) | -0.037 |
| 58 | 40 (53%) | 28 (37%) | 0.27 | 13 (17%) | 6 (8%) | -0.040 |
| 59 | 26 (35%) | 16 (21%) | 0.15 | 22 (29%) | 16 (21%) | -0.029 |
| 60 | 40 (53%) | 29 (39%) | 0.48 | 14 (19%) | 8 (11%) | -0.0032 |
| 61 | 19 (25%) | 12 (16%) | 0.091 | 23 (31%) | 18 (24%) | -0.031 |
| 62 | 31 (41%) | 19 (25%) | 0.33 | 9 (12%) | 4 (5.3%) | -0.015 |
| 63 | 21 (28%) | 14 (19%) | 0.17 | 15 (20%) | 10 (13%) | -0.033 |
| 64 | 32 (43%) | 21 (28%) | 0.31 | 18 (24%) | 12 (16%) | -0.030 |
| 65 | 33 (44%) | 21 (28%) | 0.43 | 14 (19%) | 7 (9.3%) | -0.012 |
| 66 | 32 (43%) | 21 (28%) | 0.31 | 18 (24%) | 12 (16%) | -0.030 |
| 67 | 32 (43%) | 21 (28%) | 0.31 | 18 (24%) | 12 (16%) | -0.030 |
| 68 | 32 (43%) | 20 (27%) | 0.33 | 11 (15%) | 4 (5.3%) | -0.015 |
| 69 | 35 (47%) | 22 (29%) | 0.48 | 15 (20%) | 9 (12%) | -8.6E-05 |
| 70 | 35 (47%) | 25 (33%) | 0.21 | 23 (31%) | 12 (16%) | 0.0034 |
| 71 | 32 (43%) | 21 (28%) | 0.31 | 18 (24%) | 12 (16%) | -0.030 |

**Supplementary Table S11 (cont.)**

| **Permutation** | **Logistic / binary** | | | **Linear / quantitative** | | |
| --- | --- | --- | --- | --- | --- | --- |
|  | **# nominally significant** (*p* < 0.05) | **# Bonferroni significant**  (*p* < 0.00067) | **Median coefficient (log odds ratio)** | **# nominally significant**  (*p* < 0.05) | **# Bonferroni significant**  (*p* < 0.00067) | **Median coefficient (population std dev)** |
| *True data* | *47 (63%)* | *36 (48%)* | *0.45* | *29 (39%)* | *21 (28%)* | *0.0087* |
| 72 | 48 (64%) | 32 (43%) | 0.72 | 12 (16%) | 5 (6.7%) | -0.068 |
| 73 | 26 (35%) | 17 (23%) | 0.28 | 8 (11%) | 5 (6.7%) | -0.030 |
| 74 | 26 (35%) | 16 (21%) | 0.15 | 22 (29%) | 16 (21%) | -0.029 |
| 75 | 25 (33%) | 16 (21%) | 0.20 | 15 (20%) | 12 (16%) | -0.037 |
| 76 | 19 (25%) | 12 (16%) | 0.091 | 23 (31%) | 18 (24%) | -0.031 |
| 77 | 17 (23%) | 11 (15%) | 0.089 | 9 (12%) | 7 (9.3%) | -0.044 |
| 78 | 24 (32%) | 13 (17%) | 0.079 | 11 (15%) | 6 (8%) | -0.037 |
| 79 | 28 (37%) | 16 (21%) | 0.18 | 17 (23%) | 12 (16%) | -0.011 |
| 80 | 26 (35%) | 16 (21%) | 0.22 | 7 (9.3%) | 3 (4%) | -0.030 |
| 81 | 30 (40%) | 20 (27%) | 0.32 | 8 (11%) | 5 (6.7%) | -0.026 |
| 82 | 19 (25%) | 12 (16%) | 0.091 | 23 (31%) | 18 (24%) | -0.031 |
| 83 | 33 (44%) | 21 (28%) | 0.43 | 14 (19%) | 7 (9.3%) | -0.012 |
| 84 | 30 (40%) | 19 (25%) | 0.29 | 17 (23%) | 11 (15%) | -0.043 |
| 85 | 30 (40%) | 18 (24%) | 0.20 | 10 (13%) | 3 (4%) | -0.042 |
| 86 | 48 (64%) | 32 (43%) | 0.72 | 12 (16%) | 5 (6.7%) | -0.068 |
| 87 | 11 (15%) | 6 (8%) | 0.051 | 1 (1.3%) | 1 (1.3%) | -0.030 |
| 88 | 26 (35%) | 17 (23%) | 0.28 | 8 (11%) | 5 (6.7%) | -0.030 |
| 89 | 44 (59%) | 31 (41%) | 0.60 | 22 (29%) | 11 (15%) | 0.023 |
| 90 | 26 (35%) | 17 (23%) | 0.033 | 21 (28%) | 14 (19%) | -0.037 |
| 91 | 27 (36%) | 16 (21%) | 0.13 | 15 (20%) | 7 (9.3%) | -0.090 |
| 92 | 34 (45%) | 23 (31%) | 0.32 | 17 (23%) | 12 (16%) | -0.011 |
| 93 | 20 (27%) | 15 (20%) | 0.092 | 10 (13%) | 6 (8%) | -0.036 |
| 94 | 25 (33%) | 16 (21%) | 0.20 | 15 (20%) | 12 (16%) | -0.037 |
| 95 | 48 (64%) | 32 (43%) | 0.72 | 12 (16%) | 5 (6.7%) | -0.068 |
| 96 | 38 (51%) | 27 (36%) | 0.58 | 24 (32%) | 14 (19%) | 0.007 |
| 97 | 42 (56%) | 29 (39%) | 0.28 | 13 (17%) | 6 (8%) | -0.042 |
| 98 | 28 (37%) | 17 (23%) | 0.17 | 13 (17%) | 6 (8%) | -0.037 |
| 99 | 33 (44%) | 22 (29%) | 0.16 | 20 (27%) | 10 (13%) | -0.0061 |
| 100 | 27 (36%) | 21 (28%) | 0.22 | 10 (13%) | 6 (8%) | -0.013 |

**Table S12. Estimated numbers of undiagnosed cases for 32 rare phecodes.** TPR = True Positive Rate; FPR = False Positive Rate; 95% CI = 95% bootstrap confidence interval.

| **Phecode** | **Known Cases** | **Known Controls** | **Unknown** | **TPR** | **FPR** | **Undiagnosed Cases** (95% CI) | **Underdiagnosis Rate** (95% CI) |
| --- | --- | --- | --- | --- | --- | --- | --- |
| 071.1 | 210 | 393688 | 30856 | 0.16 | 0.0015 | 85 (0-210) | 0.29 (0-0.50) |
| 079.2 | 101 | 308505 | 116039 | 0.089 | 0.0040 | 2100 (1100-5500) | 0.95 (0.91-0.98) |
| 250.12 | 167 | 219450 | 171559 | 0.96 | 0.00093 | 72 (36-130) | 0.30 (0.17-0.43) |
| 253 | 214 | 200345 | 226256 | 0.29 | 0.0069 | 1300 (1000-2000) | 0.86 (0.82-0.91) |
| 277.51 | 121 | 73548 | 357442 | 0.066 | 0.0051 | 0 (0-0) | 0 (0-0) |
| 278.3 | 111 | 200449 | 200148 | 0.018 | 0.0011 | 2000 (0-8000) | 0.95 (0-0.99) |
| 281.9 | 148 | 286567 | 103121 | 0.26 | 0.01 | 1500 (1000-2100) | 0.91 (0.88-0.94) |
| 286.11 | 180 | 267560 | 161899 | 0.017 | 0.0013 | 2700 (0-12000) | 0.94 (0-0.98) |
| 288.3 | 211 | 285933 | 132802 | 0.13 | 0.0042 | 2000 (1400-3200) | 0.91 (0.85-0.94) |
| 289.9 | 188 | 385851 | 32884 | 0.15 | 0.013 | 1900 (1200-3000) | 0.91 (0.86-0.94) |
| 315.3 | 196 | 415882 | 18441 | 0.097 | 0.0017 | 510 (300-910) | 0.72 (0.59-0.83) |
| 366.1 | 118 | 233222 | 142628 | 0.025 | 4.8e-4 | 740 (0-3400) | 0.86 (0-0.97) |
| 377.1 | 163 | 111789 | 302902 | 0.14 | 0.0025 | 1900 (1100-3100) | 0.92 (0.87-0.95) |
| 459.1 | 200 | 238256 | 143208 | 0.040 | 0.0011 | 1600 (620-4300) | 0.89 (0.75-0.96) |
| 519 | 195 | 257213 | 156115 | 0.34 | 0.0061 | 350 (130-570) | 0.64 (0.40-0.76) |
| 588 | 194 | 368719 | 23280 | 0.40 | 0.0065 | 300 (180-380) | 0.60 (0.49-0.68) |
| 649 | 153 | 111327 | 127928 | 0.65 | 0.043 | 0 (0-0) | 0 (0-0) |
| 686.5 | 103 | 263302 | 150499 | 0.049 | 5.5e-4 | 990 (270-3700) | 0.91 (0.73-0.97) |
| 691 | 150 | 416622 | 5239 | 0.83 | 9.6e-4 | 6.0 (0-16) | 0.039 (0-0.092) |
| 704.8 | 143 | 270384 | 155993 | 0.0070 | 0.0011 | 720 (0-20000) | 0.83 (0-0.99) |
| 706.1 | 145 | 267782 | 154404 | 0.00 | 0.0010 | 0 (0-29000) | 0 (0-1) |
| 709.6 | 137 | 355833 | 60146 | 0.19 | 0.0023 | 0 (0-99) | 0 (0-0.47) |
| 737 | 147 | 254307 | 155828 | 0.20 | 0.012 | 990 (510-1700) | 0.87 (0.76-0.92) |
| 755.61 | 201 | 269348 | 164699 | 0.12 | 0.0013 | 0 (0-250) | 0 (0-0.57) |
| 756 | 115 | 301041 | 133006 | 0.00 | 8.8e-4 | 0 (0-11000) | 0 (0-0.99) |
| 756.3 | 170 | 301041 | 133006 | 0.082 | 0.0035 | 0 (0-250) | 0 (0-0.61) |
| 758 | 144 | 345306 | 88724 | 0.15 | 0.0037 | 320 (79-710) | 0.69 (0.34-0.84) |
| 758.1 | 218 | 426070 | 7960 | 0.014 | 0.0013 | 1100 (0-5900) | 0.83 (0-0.97) |
| 794 | 176 | 100444 | 335787 | 0.45 | 0.029 | 0 (0-0) | 0 (0-0) |
| 797 | 150 | 211755 | 224188 | 0.23 | 0.015 | 4700 (3500-7400) | 0.97 (0.96-0.98) |
| 913 | 203 | 316493 | 119711 | 0.0099 | 2.6e-4 | 1000 (0-4500) | 0.84 (0-0.95) |
| 989 | 109 | 315329 | 119203 | 0.11 | 0.0025 | 960 (470-2300) | 0.90 (0.8-0.96) |

**Supplementary Table S13. Description of model hyperparameters and final tuned values.** CV = Cross Validation.

| **Name** | **Description** | **Tuned Value** | | | | | |
| --- | --- | --- | --- | --- | --- | --- | --- |
|  |  | **CV 1** | **CV 2** | **CV 3** | **CV 4** | **CV 5** | **Full** |
| Input_deletion | Rate of random token deletion in training input | 0.001 | 0.001 | 0.001 | 0.001 | 0.001 | 0.001 |
| decoder_layers | Number of transformer decoder layers | 1 | 1 | 1 | 1 | 1 | 1 |
| dense_dim | Size of transformer feed-forward layers | 256 | 256 | 256 | 256 | 256 | 256 |
| attention_heads | Number of attention heads in transformer attention layers | 3 | 3 | 3 | 3 | 3 | 3 |
| internal_dropout | Rate of dropout regularization inside transformer layer | 0.2 | 0.2 | 0.2 | 0.2 | 0.2 | 0.2 |
| lr | Learning rate for Adam optimizer | 0.0001 | 0.0001 | 0.0001 | 0.0001 | 0.0001 | 0.0001 |
| epochs | Epoch with best validation loss | 4 | 3 | 3 | 4 | 3 | 3 |
